# The Application of Artificial Intelligence in Healthcare Practice: An Umbrella Review

**DOI:** 10.1101/2025.10.05.25337356

**Authors:** Adam Andersen, Ruiping Huang, Edward Jiusi Liu

## Abstract

Artificial intelligence (AI) is rapidly transforming healthcare practice, with growing evidence supporting its use in diagnosis, prognosis, treatment planning, and operational decision-making. The proliferation of systematic reviews in recent years underscores the need for an updated synthesis of the literature to inform research, policy, and practice. We searched PubMed, Web of Science, Scopus, IEEE Xplore, and CINAHL for systematic reviews and meta-analyses published between 2019 and November 2024. Eligible reviews focused on AI applications in healthcare practice, were peer-reviewed, and written in English. A total of 181 reviews met the inclusion criteria. Publication volume increased steadily, peaking in 2024. AI research was concentrated in high-density domains, such as radiology, oncology, and critical care. Across reviews, diagnostic imaging, electronic health record (EHR) data, and biomarkers/laboratory results accounted for 70% of training data sources, though newer data types, such as wearable device and sensor data, emerged from 2022 onward. Diagnosis, prognosis, and treatment comprised over 80% of AI applications, with novel uses emerging in recent years. Ethical concerns were reported in 64.6% of reviews, with privacy, model accuracy, data and algorithmic bias, and explainability as recurrent themes. The proportion of reviews reporting ethical concerns increased from 2021 to 2024. AI applications in healthcare are expanding in scope, diversifying in data sources, and evolving toward novel clinical and operational uses. The human-centered AI or Human-AI-Human paradigm, integrating computational precision with clinical expertise, holds significant promise but will require parallel advances in governance, regulatory frameworks, and ethical oversight to ensure safe adoption.

**Article highlights:**

- This umbrella review summarizes 181 systematic reviews on AI applications across healthcare practice fields.
- 70% of the reviews reported AI models trained on diagnostic imaging, EHR, and biomarker data.
- There has been recent growth in AI using wearable, sensor, and other health data.
- Diagnosis, prognosis, and treatment comprised over 80% of the applications in all the reviews.
- Ethical concerns, including privacy, accuracy, bias, and explainability, were raised in over half of the reviews.

## 1. Introduction

Artificial intelligence (AI) is rapidly developing and playing an increasingly important role in healthcare practice (Goktas & Grzybowski, 2025). AI technologies applied in healthcare offer opportunities to facilitate diagnosis, treatment, and prognosis, among other applications. The number of studies and reviews focusing on AI in healthcare practice has dramatically increased in the past decade (Ali et al., 2023). For example, according to PubMed, 2879 reviews focusing on AI in healthcare have been published from 2020 to September 2025. The growing number of AI reviews demonstrates its potential to improve healthcare outcomes on a large scale. This study synthesizes recent systematic reviews on AI applications in healthcare to map the current landscape, track the recent trends, and help guide future research.

AI research increasingly demonstrates that combining human expertise with AI capabilities can produce superior outcomes compared to either working in isolation (Zöller et al., 2025; Anderson et al., 2023). This combination is now referred to as human-AI collaboration, human-centered AI, or Human-AI-Human (H-AI-H) (OSPI, 2024). In clinical settings, H-AI-H decision-making has the potential to improve diagnostic and prognostic accuracy, generate effective treatment options, and enhance care planning by integrating computational precision with clinical judgment and contextual awareness (Oberije et al., 2024; Liu et al., 2025; Lång et al., 2023). For example, AI models may rapidly analyze imaging or laboratory data, while clinicians validate results, consider patient-specific factors, and make the final decision. This collaborative paradigm not only leverages the strengths of both humans and machines but also addresses some of the concerns related to overreliance on autonomous systems.

With the rapid advancement of AI, a large number of review studies have summarized the AI development and its growing integration into clinical workflows. Loh et al. (2022) conducted a systematic review of AI applications in healthcare from 2011 to 2022. The authors identified several categories of AI applications, including imaging and test results interpretation, diagnosis, treatment, patient data management, healthcare administration, and predictive medicine. The AI applications span many medical specialties and diseases. For example, Rashid et al. (2022) conducted a systematic review of AI applications in acute respiratory distress syndrome (ARDS). The findings included several applications with high levels of accuracy, including diagnosis, prognosis, and treatment. Xu & Xu (2024) conducted a systematic review of machine learning (ML) applications in preventative healthcare. Specifically, the use of ML to prevent or manage chronic disease, which represented several of the top ten leading causes of death worldwide in 2021 (WHO, 2021).

To synthesize those reviews, an umbrella review is needed. An umbrella review, also known as a review of reviews, is particularly suited to synthesizing a large and diverse body of evidence (Grant & Booth, 2009). Kolasa et al. (2024) conducted a literature review of systematic reviews of machine learning (ML) in healthcare and greatly synthesized the knowledge and improved our understanding of this important topic. The authors identified 220 systematic reviews published between 2010 and March 2023. They analyzed the reviews and showed the distribution of publication year, diagnoses, data types used to train models, and types of machine learning models used. They found that the most common diagnoses studied were neoplasms and diseases of the nervous system. Imaging and clinical notes were the two leading types of data sources used to train AI models. Overall, their findings highlighted both the rapid growth of ML-related healthcare research and gaps in explainability, validation, and integration into clinical workflows, contributing to our understanding of AI in healthcare.

Building on the previous reviews, we can further advance the understanding of AI in healthcare by synthesizing the reviews in the following ways:

(a) An update is needed to capture the recent advancement of AI applications in healthcare. Kolasa et al. (2024) reviewed the reviews up to March 2023, and 88% of the included reviews were published between 2020 and 2021. AI technology and research have advanced dramatically since then. Notably, the release of ChatGPT in late 2022 and the rapid diffusion of large language models (LLMs) in subsequent years have marked a major turning point in AI research and applications (Liu et al., 2023). With the advancement of AI applications in healthcare, reviews on AI in healthcare have skyrocketed in recent years. According to PubMed, 648 reviews focused on AI applications in healthcare practice were published between April 2023 and November 2024.
(b) A longitudinal analysis is needed to capture how themes have changed over time, such as data types and applications. A longitudinal analysis would help us understand the progress of AI in healthcare and predict its future development. For example, Cicek and Bagci (2025) identified new data types and applications. New data types, such as remote monitoring data, can allow for earlier detection and diagnosis. New applications, such as automating clinical documentation via AI-driven ambient listening tools, have been implemented into clinical practice.
(c) Ethical concerns need to be captured. Issues such as privacy, accuracy, bias, and explainability are significant challenges in healthcare (Bouderhem, 2024). To our knowledge, the analysis of ethical concerns about AI in healthcare and their evolution over time has not been extensively researched.

To fill in the gaps, we conduct an umbrella review, addressing the following research questions:

1. What are the characteristics of the review studies on AI applications in healthcare practice?
2. Healthcare data types

a. What types of healthcare data have been used to train AI models?
b. How have the healthcare data types changed over time?
3. AI healthcare applications

a. What AI tools have been applied in healthcare practice?
b. How have AI tools in healthcare practice changed over time?
4. Ethical concerns

a. What are the ethical concerns about AI applications in healthcare practice?
b. How have ethical concerns changed over time?

By answering these questions, we aim to provide a comprehensive overview of AI in healthcare practice, identify key trends, and highlight the gaps and limitations. This approach offers a holistic perspective on the current state of AI in healthcare practice, informs scholars entering the AI field, and guides future endeavors in this rapidly evolving area.

## 2. Methods

To answer the research questions, we conducted an umbrella review and managed the review process using Covidence, a systematic review data management software (https://www.covidence.org/). During the review process, we referenced the widely adopted Preferred Reporting Items for Systematic Reviews and Meta-Analyses (PRISMA) checklist (Page et al., 2021) (Figure 1). This section presents the search strategy, inclusion and exclusion criteria, screening process, data extraction and quality assessment, and data analysis methods.

**Figure 1:**
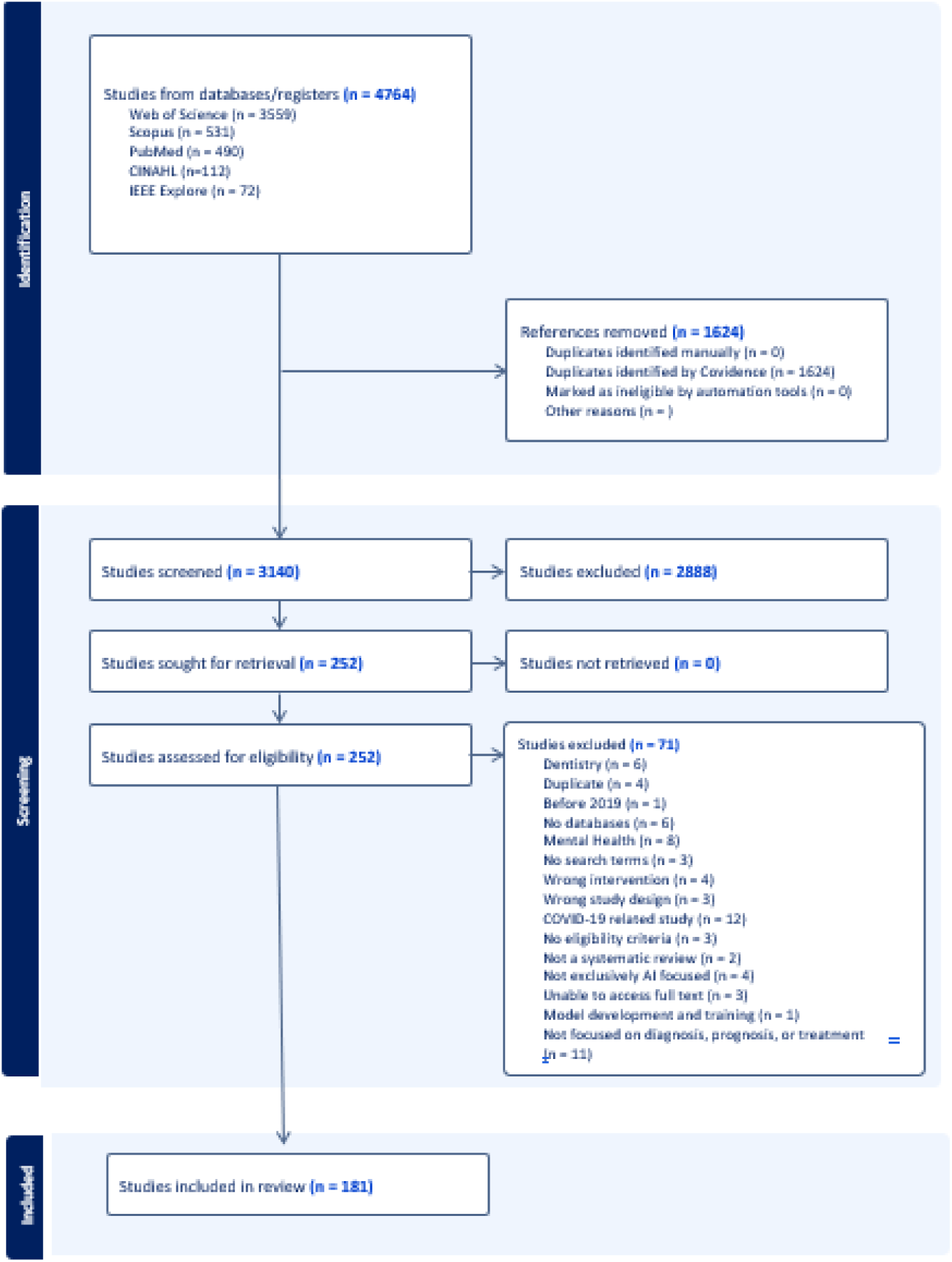
Search and screening processes exported from Covidence

### 2.1. Search Strategy and Study Selection

#### Keyword Search

In November 2024, the first author searched five databases: PubMed, Web of Science, Scopus, IEEE Xplore, and CINAHL. These databases were selected for their relevance to healthcare and AI, ensuring comprehensive coverage in our umbrella review. We used keywords related to AI and healthcare practice, (“artificial intelligence” OR “AI” OR “chatgpt” OR “large language models” OR “machine learning” OR “deep learning” OR “neural networks” OR “predictive analytics”) AND (“Delivery of Health Care” OR “healthcare” OR “health care”) AND (“Medicine” OR “Dentistry” OR “Nursing” OR “Pharmacy” OR “Allied Health” OR “Public Health” OR “Health Services administration” OR “Social work” OR “therapy” OR “diagnosis” OR “prognosis” OR “surgery” OR “prevention” OR “risk”) AND (‘Systematic review”). The initial search yielded 3140 articles after duplicates were removed.

### 2.2. Inclusion and Exclusion Criteria

We generated inclusion and exclusion criteria from the iteration of team brainstorming, discussion, and pilot testing (Table 1). Two researchers screened each paper independently in Covidence and included reviews that meet the following criteria: (a) articles published between 2019 and November 2024, (b) systematic reviews, meta-analyses, or scoping reviews; (c) focusing on AI applications in healthcare practice; (d) AI technology as the central focus; (e) peer-reviewed journal articles and conference papers; and (f) English articles.

**Table 1.** Inclusion and exclusion criteria.

| Criteria | Inclusion | Exclusion |
| --- | --- | --- |
| Publication Year | 2019 to November 2024 | Articles published before 2019 |
| Study Design | Systematic review / Meta-analysis / Scoping review (reviews that show at least the three elements: searching database, search terms, and inclusion and exclusion criteria) | Non-systematic reviews (reviews that don't show the three elements: searching database, search terms, and inclusion and exclusion criteria) |
| Review Focus | Reviews focus on AI applications in healthcare practice (AI tools that can be used by a healthcare professional to provide care for a patient in a healthcare setting (hospital or clinic). | 1. AI tools not applied to an aspect of healthcare practice, such as education and research.<br>2. Reviews focused on COVID-19, Dentistry, or Psychiatry and Mental Health<br>3. Not focused on AI or only partially focused on AI:<br>(1) Reviews that do not report on any aspect of AI (applications, impact, interpretation, or technologies).<br>(2) Reviews that focus on topics like augmented reality (AR), virtual reality (VR), blockchain, metaverse, or other technologies that may incorporate AI partially or not at all, rather than having AI as the central focus. |
| Source of Publication | Peer-reviewed journals and conference papers | White papers, book chapters, technical reports, dissertations, and empirical studies |
| Language | English | Not English |

The inclusion and exclusion criteria were designed to balance comprehensiveness with relevance and rigor. (a) Publication Year: Restricting the timeframe to 2019–2024 ensured that the umbrella review captured the rapid acceleration of AI research following key technological advances, including the advent of LLMs, while excluding earlier literature less reflective of the current state of the field. According to PubMed, the number of systematic reviews started to substantially increase in 2019 and every year after. (b) Study Design: Limiting to systematic reviews, meta-analyses, and scoping reviews provided a consistent level of methodological quality and transparency in search, selection, and synthesis processes, while excluding narrative reviews or technical papers that lacked systematic rigor. (c) Review Focus: Focusing specifically on AI applications in healthcare practice, rather than adjacent domains such as education or basic algorithm development, allowed for clearer insights into clinical and operational implications. In addition, reviews were excluded if they focused on COVID-19, dentistry, or psychiatry and mental health. These areas were excluded to maintain a focused scope on broader AI applications across general medical specialties. COVID-19, dentistry, and psychiatry often feature unique AI challenges and extensive dedicated review literature. (d) Source of Publication: Restricting the reviews to peer-reviewed journal articles and conference papers ensured scholarly vetting, while excluding grey literature minimized concerns about varying quality. Finally, (e) Language: We only reviewed English-language publications due to the constraints of the research team, while acknowledging that this may have excluded relevant studies in other languages. Collectively, these criteria were intended to create a dataset that was both representative of the state of knowledge and methodologically robust to support meaningful synthesis.

### 2.3. Screening Process

After we selected papers and removed the duplicates (*n* = 3140), we screened the remaining papers in two steps. First, the first and second authors independently screened the titles and abstracts of 229 articles. Title and abstract screening inter-rater agreement had a Cohen’s kappa of 0.82, which was interpreted as excellent inter-rater agreement (Landis & Koch, 1977). We resolved the discrepancies through group discussions among the three authors. With inter-rater agreement established, the first author screened the remaining articles, resulting in 252 articles for full-text screening.

The first and third authors screened seven papers independently based on the full texts. Full text screening inter-rater agreement had a Cohen’s kappa of 1.0, which was interpreted as excellent inter-rater agreement. With inter-rater agreement established, the first author screened the remaining articles, resulting in 181 articles for data extraction.

### 2.4. Data Extraction

The first author drafted the initial data extraction rubric (Table 2) based on the research questions. The research team then collaboratively refined these rubrics, pilot-tested them with 10 papers through discussion and iterative revisions, and finalized them when no more revisions were needed.

**Table 2.**
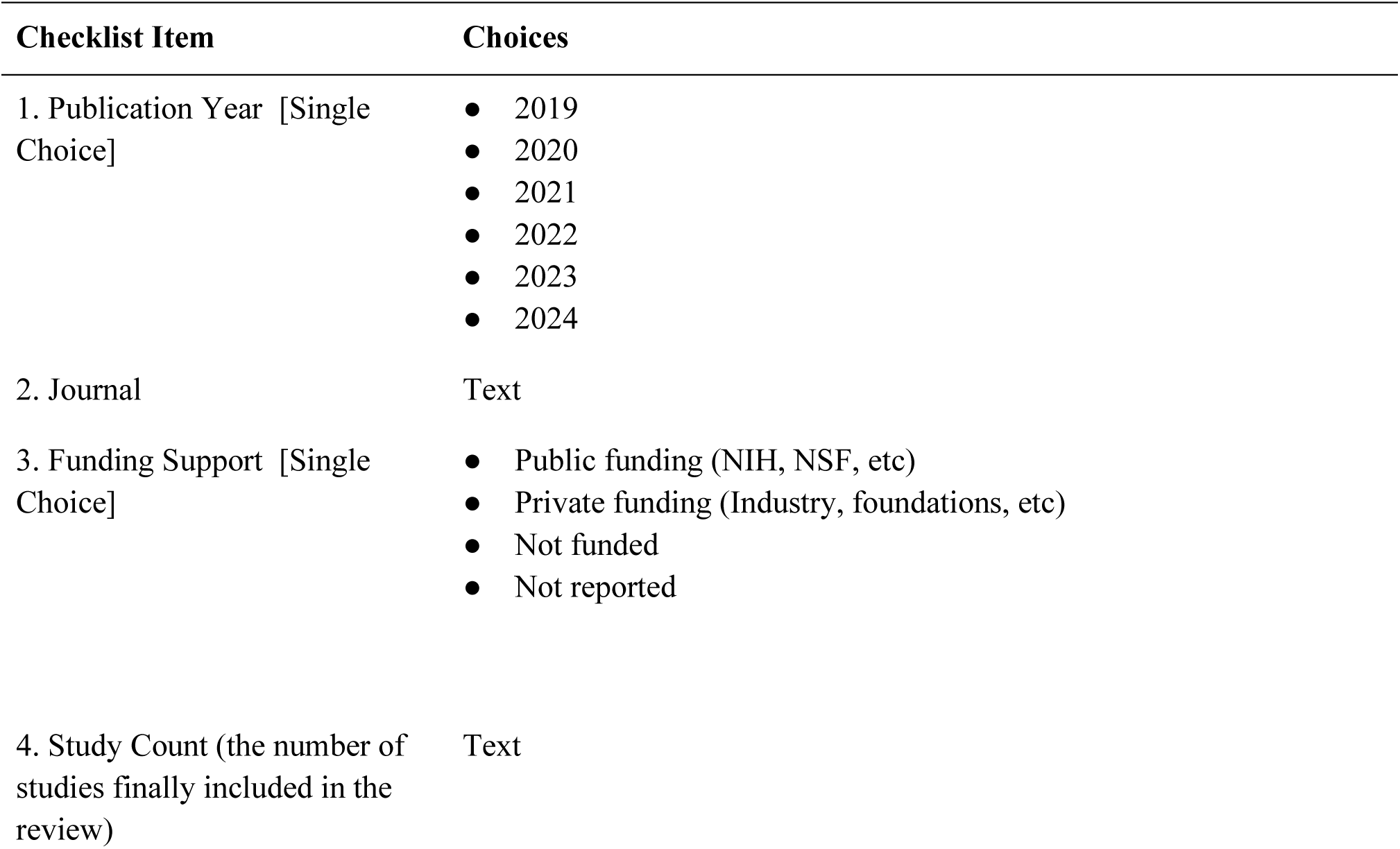
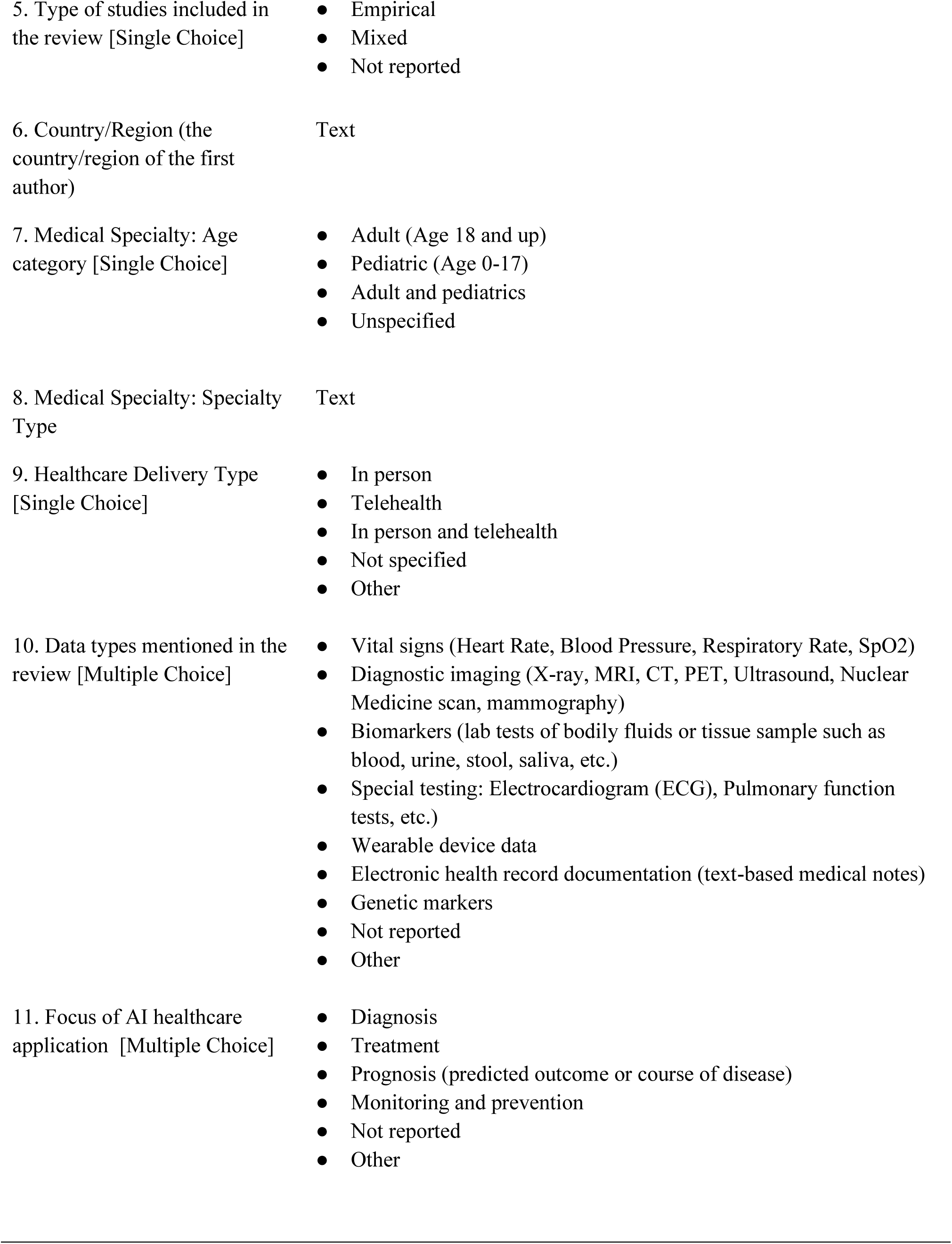
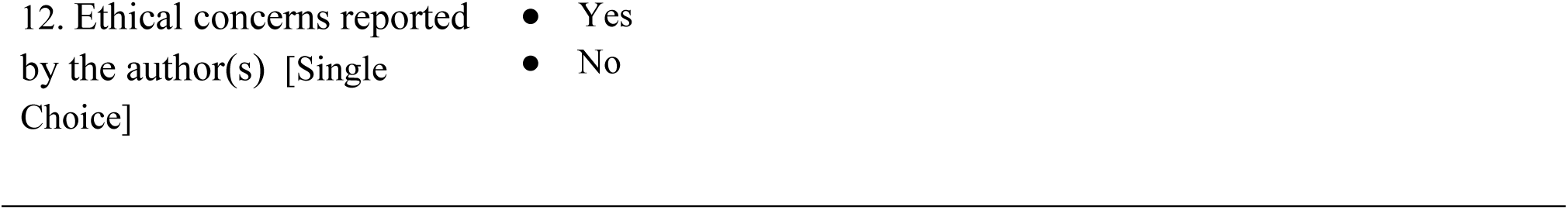
Extraction rubric.

| Checklist Item | Choices |
| --- | --- |
| 1. Publication Year [Single Choice] | ● 2019<br>● 2020<br>● 2021<br>● 2022<br>● 2023<br>● 2024 |
| 2. Journal | Text |
| 3. Funding Support [Single Choice] | ● Public funding (NIH, NSF, etc)<br>● Private funding (Industry, foundations, etc)<br>● Not funded<br>● Not reported |
| 4. Study Count (the number of studies finally included in the review) | Text |
| 5. Type of studies included in the review [Single Choice] | ● Empirical<br>● Mixed<br>● Not reported |
| 6. Country/Region (the country/region of the first author) | Text |
| 7. Medical Specialty: Age category [Single Choice] | ● Adult (Age 18 and up)<br>● Pediatric (Age 0-17)<br>● Adult and pediatrics<br>● Unspecified |
| 8. Medical Specialty: Specialty Type | Text |
| 9. Healthcare Delivery Type [Single Choice] | ● In person<br>● Telehealth<br>● In person and telehealth<br>● Not specified<br>● Other |
| 10. Data types mentioned in the review [Multiple Choice] | ● Vital signs (Heart Rate, Blood Pressure, Respiratory Rate, SpO2)<br>● Diagnostic imaging (X-ray, MRI, CT, PET, Ultrasound, Nuclear Medicine scan, mammography)<br>● Biomarkers (lab tests of bodily fluids or tissue sample such as blood, urine, stool, saliva, etc.)<br>● Special testing: Electrocardiogram (ECG), Pulmonary function tests, etc.)<br>● Wearable device data<br>● Electronic health record documentation (text-based medical notes)<br>● Genetic markers<br>● Not reported<br>● Other |
| 11. Focus of AI healthcare application [Multiple Choice] | ● Diagnosis<br>● Treatment<br>● Prognosis (predicted outcome or course of disease)<br>● Monitoring and prevention<br>● Not reported<br>● Other |

**Table 3.**
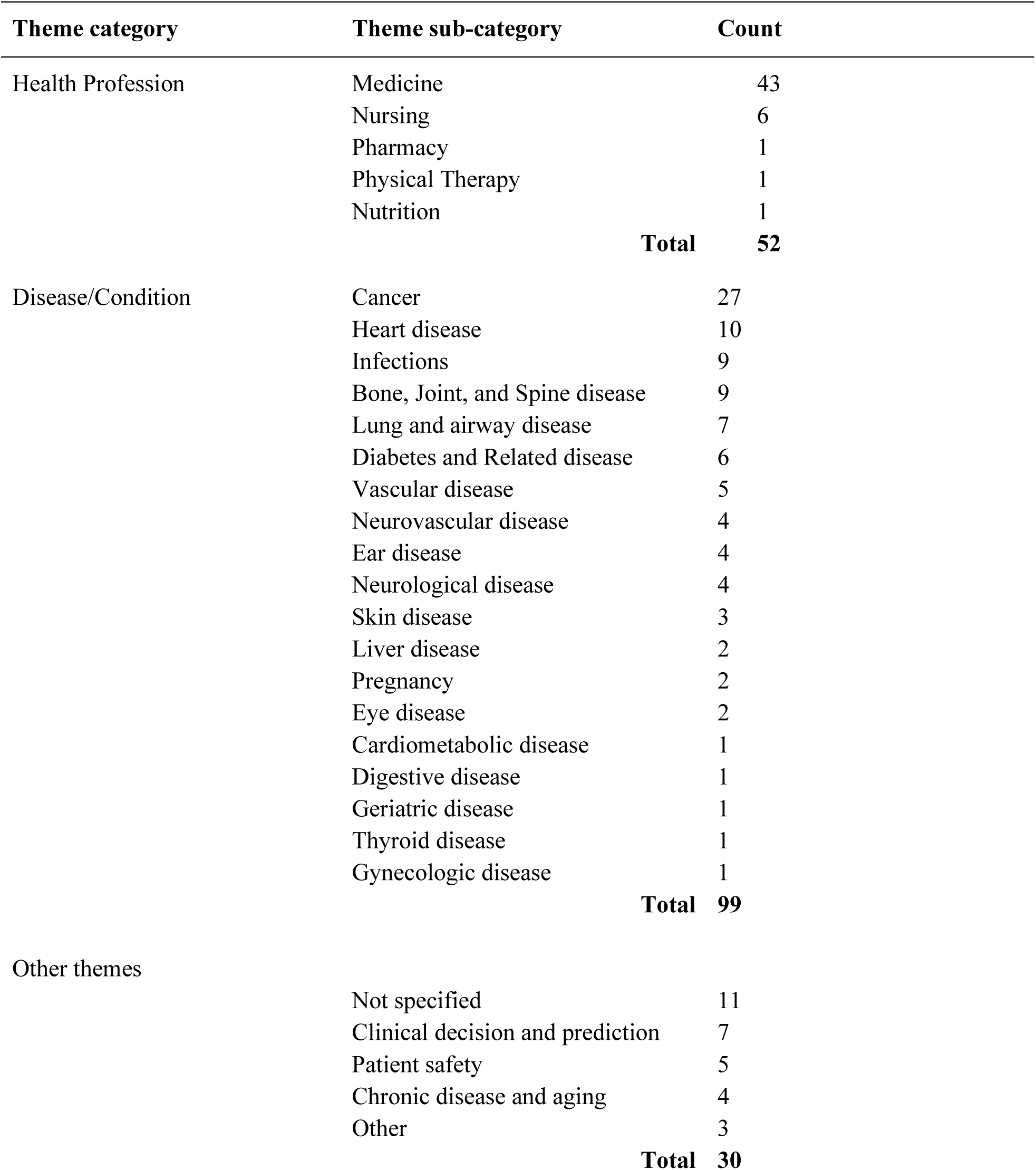
Thematic Categories of the Reviews.

| Theme category | Theme sub-category | Count |
| --- | --- | --- |
| Health Profession | Medicine | 43 |
|  | Nursing | 6 |
|  | Pharmacy | 1 |
|  | Physical Therapy | 1 |
|  | Nutrition | 1 |
|  | **Total** | **52** |
| Disease/Condition | Cancer | 27 |
|  | Heart disease | 10 |
|  | Infections | 9 |
|  | Bone, Joint, and Spine disease | 9 |
|  | Lung and airway disease | 7 |
|  | Diabetes and Related disease | 6 |
|  | Vascular disease | 5 |
|  | Neurovascular disease | 4 |
|  | Ear disease | 4 |
|  | Neurological disease | 4 |
|  | Skin disease | 3 |
|  | Liver disease | 2 |
|  | Pregnancy | 2 |
|  | Eye disease | 2 |
|  | Cardiometabolic disease | 1 |
|  | Digestive disease | 1 |
|  | Geriatric disease | 1 |
|  | Thyroid disease | 1 |
|  | Gynecologic disease | 1 |
|  | **Total** | **99** |
| Other themes | Not specified | 11 |
|  | Clinical decision and prediction | 7 |
|  | Patient safety | 5 |
|  | Chronic disease and aging | 4 |
|  | Other | 3 |
|  | **Total** | **30** |

Using the finalized rubric (Table 2), we coded the review articles to answer the research questions: (a) RQ1, Characteristics of AI applications: Publication year, journal, funding support, study count, study type, first author country of origin, medical specialty, delivery type; (b) RQ2, Data types; (c) RQ3, AI tools used in healthcare practice; and (d) RQ4, Ethical concerns: Ethical concerns reported by the author(s).

Using the finalized rubrics, the first and third authors independently coded 10 randomly selected studies to establish inter-rater agreement and resolved conflicts through group discussion. Cohen’s kappa was 0.82, indicating excellent inter-rater agreement. The first and third authors then independently coded the remaining papers, resolving ambiguities through group discussion to ensure accuracy and consistency.

### 2.5. Data Analysis

We systematically examined the selected reviews quantitatively and qualitatively. Quantitatively, we calculated frequencies and proportions for categorical variables, such as publication year, funding support, medical specialties, application, and data types, and presented the major results through tables and figures. Qualitatively, we synthesized themes and patterns that add more depth to answering the research questions. We identified the trends and gaps and recommended how to advance future research on AI applications in healthcare practice.

## 3. Results

### 3.1. RQ1: Characteristics of AI applications in healthcare practice

We described the characteristics of the reviews from multiple dimensions. Figures 2 through 5 show the general information about the reviews, including publication year, country, journals, and medical specialty. (a) The number of reviews consistently increased from 2019 to 2024, peaking at 73 in 2024 (Figure 2). The number of reviews in 2019 and 2020 is much smaller than the later years. (b) The first author’s country of origin, the top four common countries/regions for the first author were the United States (n = 21, 11.6%), the United Kingdom (n = 13, 7.2%), Australia and India (n = 10, 5.5% respectively), and China, Italy, and Saudi Arabia (n = 9, 5% respectively) (Figure 3). (c) The reviews appeared in 133 journals, with the top five journals ranging from two to 10 total reviews (Figure 4). (d) The reviews can be categorized into three main themes: health profession, disease/condition, and other. The three themes can be further classified into sub-categories, in particular, health professions into five and disease/condition into 19 sub-categories. (e) Figure 5 linked the medical specialties to disease/condition and the other themes. Many connections exist between specialties, diseases/conditions, and applications with AI tools, creating a network effect. Medical specialties such as radiology, critical care, and emergency medicine, and diseases such as cancer, heart disease, and infection have more linkages than other specialties and diseases. These highly linked themes have more AI-related research. The themes with fewer linkages indicate opportunities for more research, such as gerontology, geriatric disease, and gynecologic disease. (f) The patient age group was mostly unspecified (n = 135, 75%), with the remaining reviews focusing on adults (n = 24, 13%), pediatrics (n = 7, 4%), or adults and pediatrics (n = 15, 8%). (g) The healthcare delivery type was mostly not specified (n=176, 97%), with the remaining reviews reporting in person care or telehealth and in person care.

**Fig. 2.**
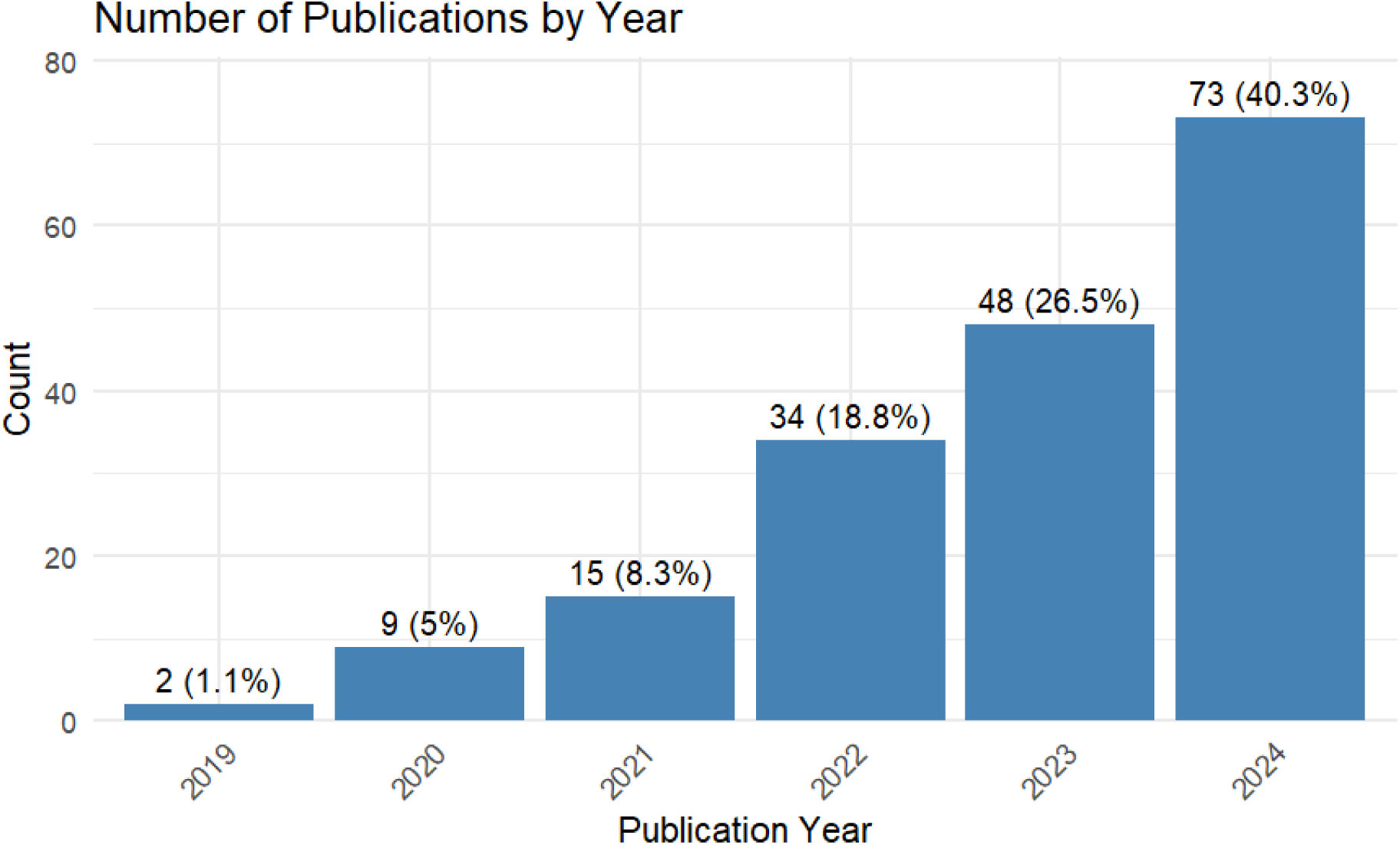
Number of publications by year (n=181)

**Fig. 3.**
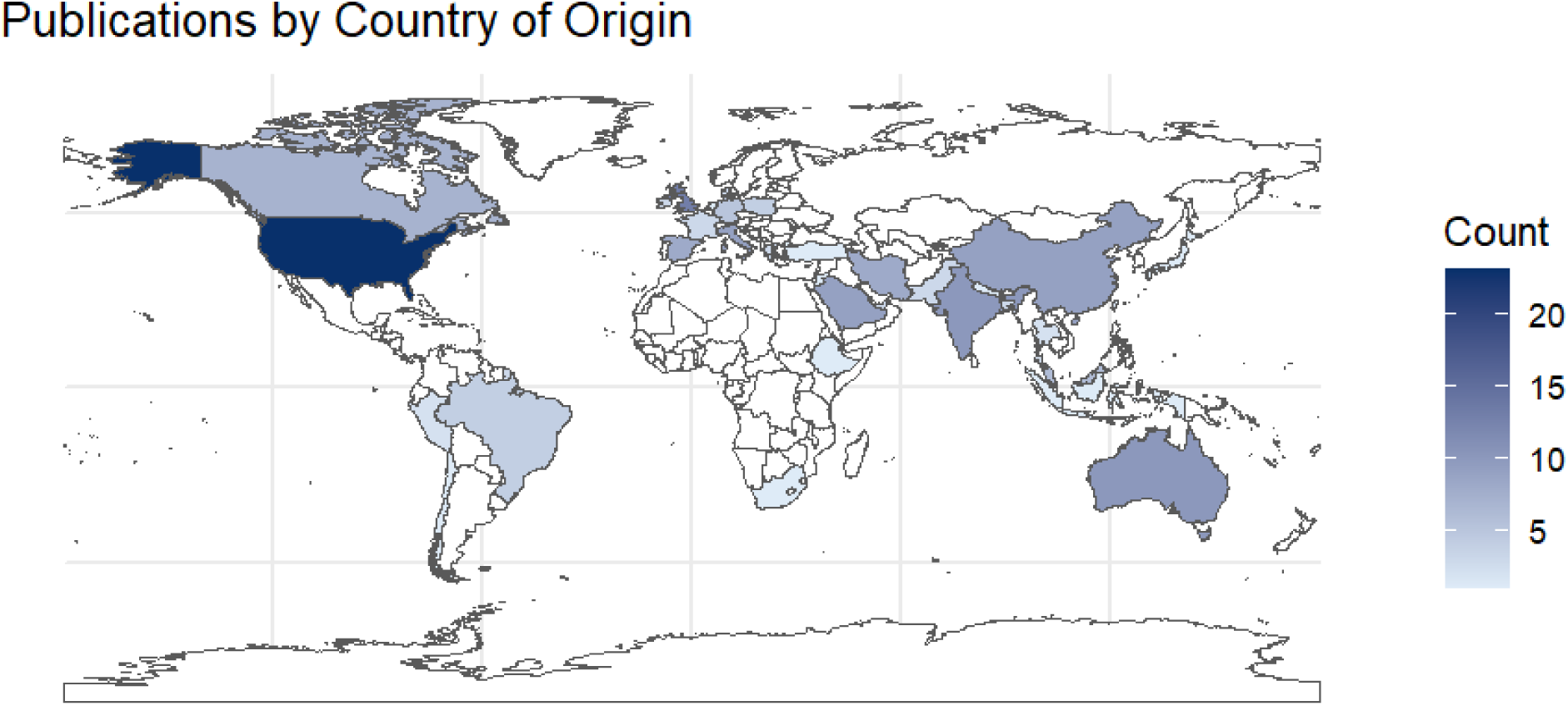
First author(s) country/region of origin

**Fig. 4.**
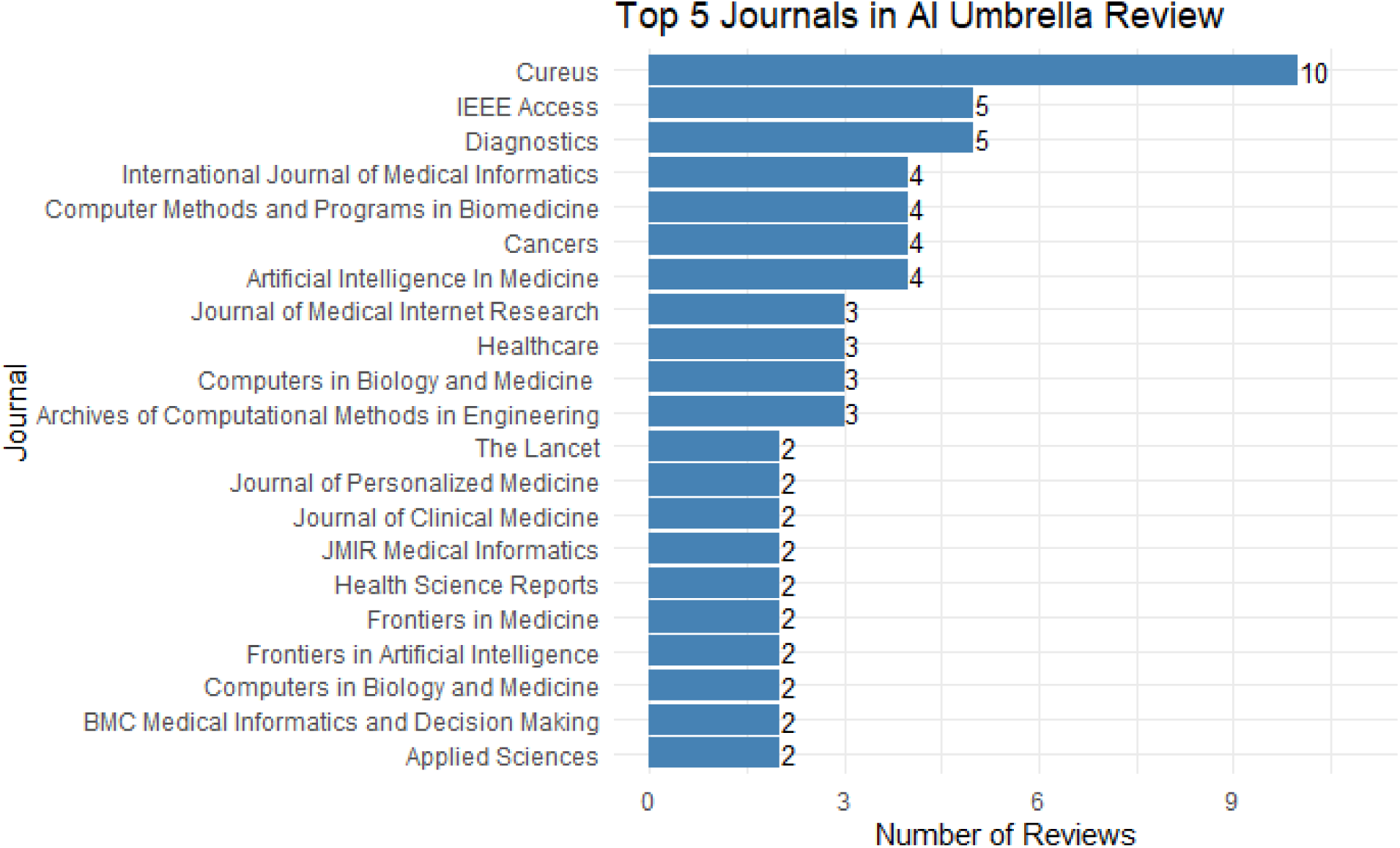
Counts of the top five journals included in the umbrella review

**Fig. 5.**
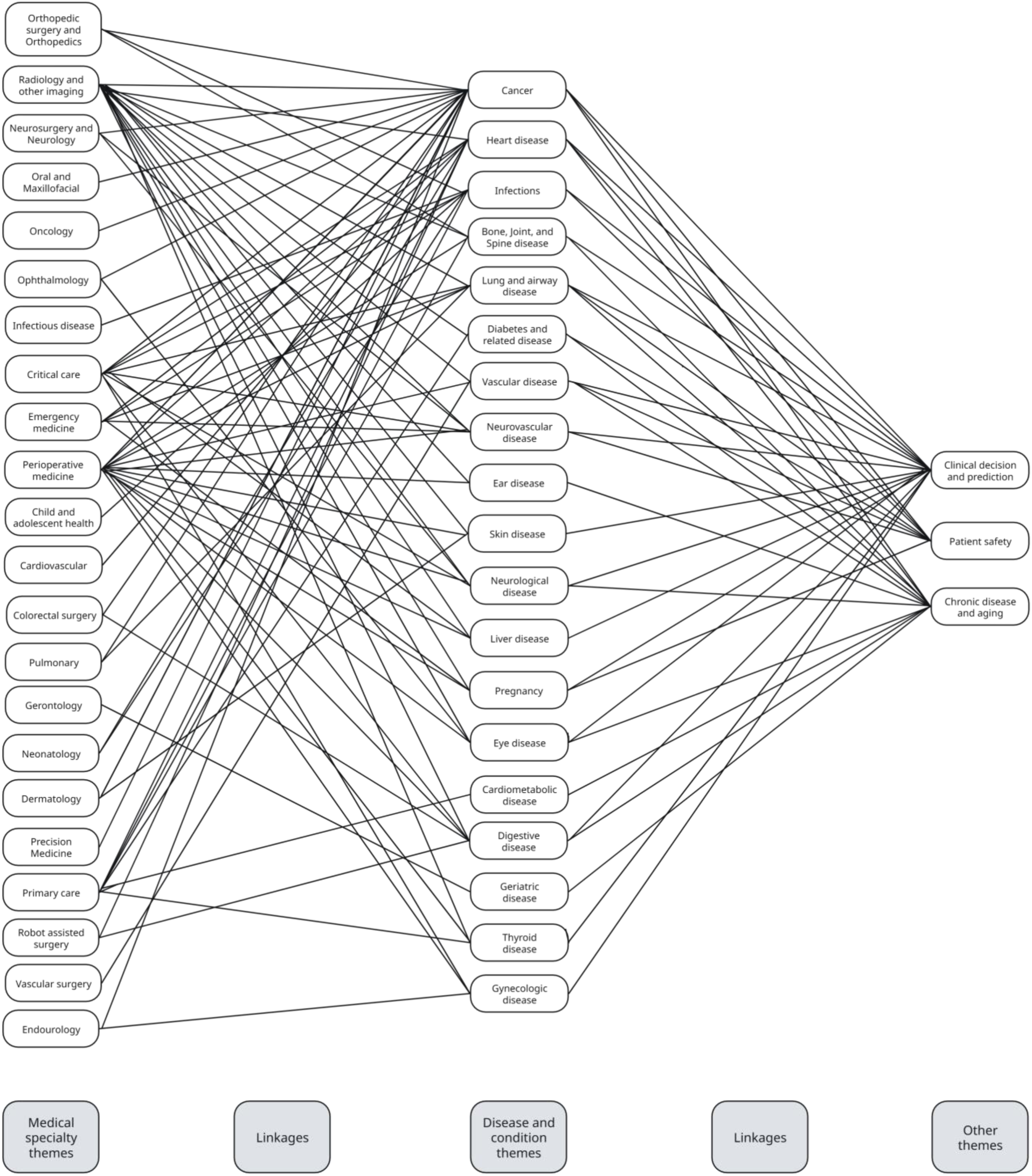
Linkages between medical specialties, disease/conditions, and other themes

### 3.2. RQ2a and b: Healthcare data types used to train AI models and change over time

The types of healthcare data used to train AI models were extracted during the full-text review phase of the umbrella review. As shown in Figure 6, three data types comprised 70% of the data types in all the reviews: diagnostic imaging, electronic health record (EHR), and biomarkers and laboratory tests. As one review may report more than one data type, the total count is above 181. Figure 7 shows how the data type changes over time. Similarly, the aforementioned three data types comprise a majority of the data types each year. Wearable device data and other data types appear in greater numbers in 2023 and 2024. This trend suggests that new data types are emerging to train AI models. This topic is explored further in the narrative review.

**Fig. 6.**
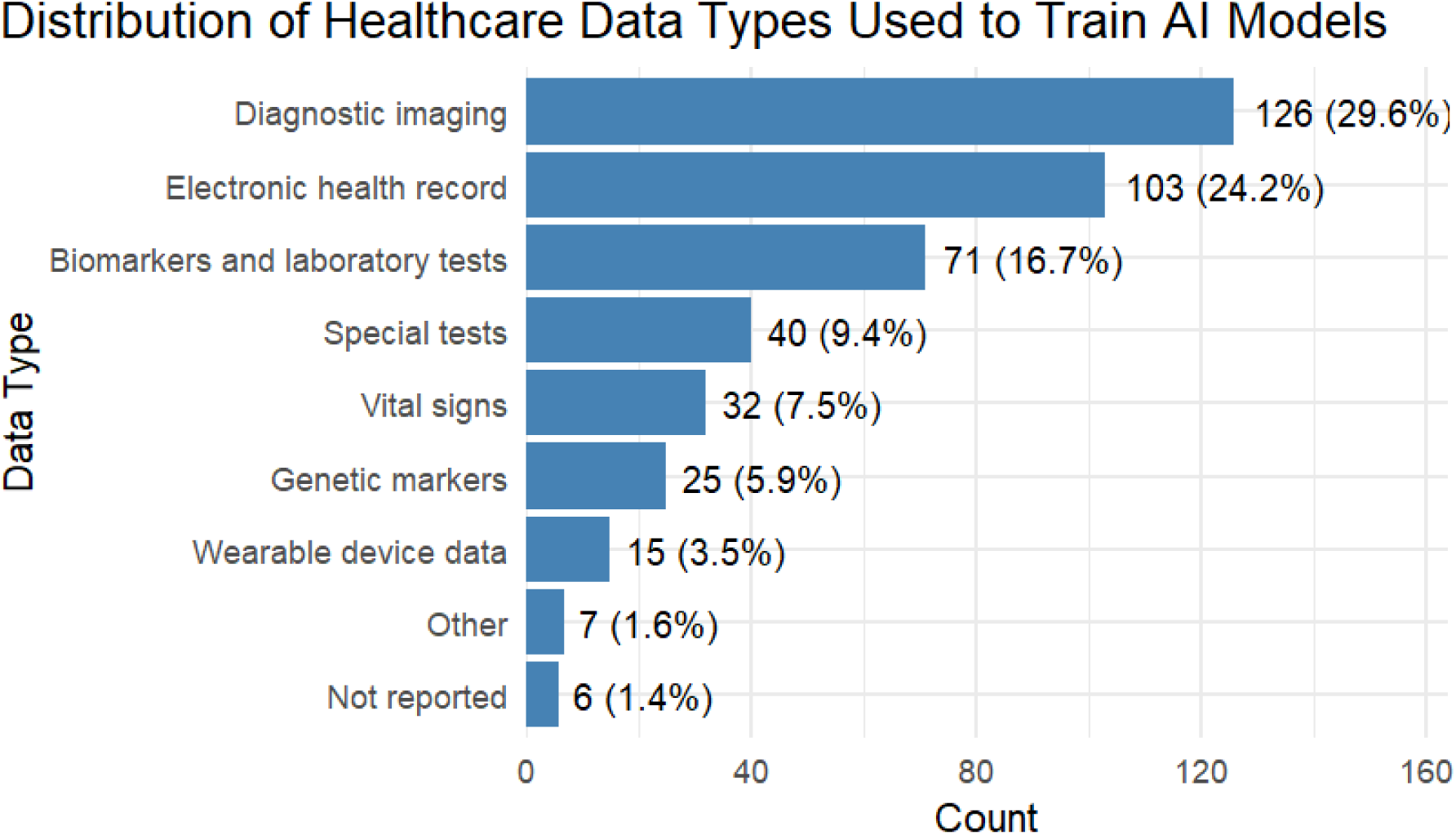
Distribution of healthcare data types used to train AI models

**Fig. 7.**
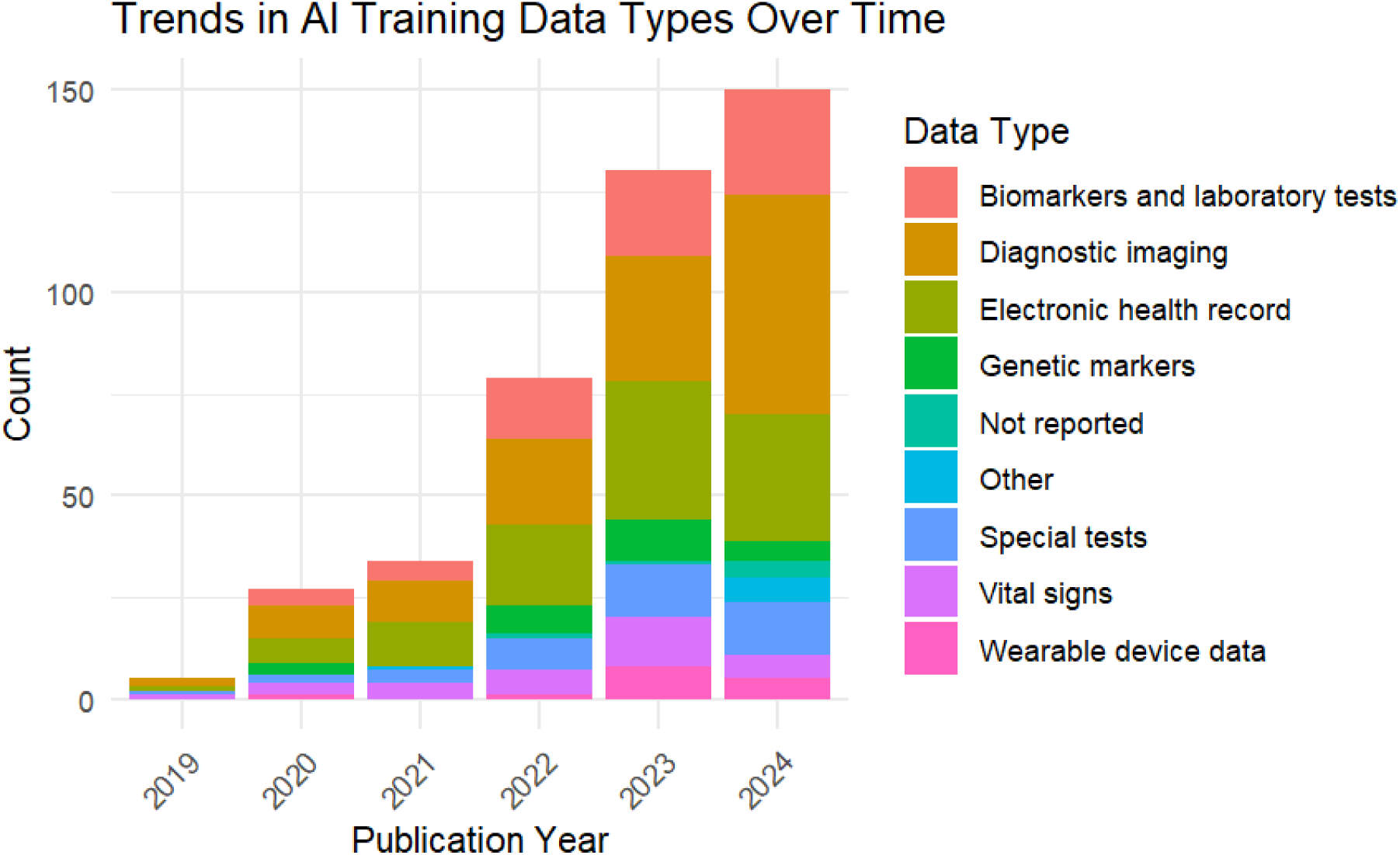
Distribution of healthcare data types over time

#### 3.2.1. Narrative review of healthcare data types used to train AI models

Across systematic reviews of AI applications in healthcare, diagnostic imaging, EHR, and biomarkers/laboratory test results consistently dominate model development. Diagnostic imaging was the leading source of data used to train AI models in this review. Imaging data includes radiographs, CT scans, MRIs, photographs, and other images. Islam et al. (2020) investigated the accuracy of deep learning (DL) algorithms to detect diabetic retinopathy in retinal fundus photograph. The results showed that DL algorithms had high sensitivity and specificity for detecting retinopathy. Sensitivity and specificity, the proportion of true positives and true negatives, respectively, are critical measures for diagnostic tests (Pacurari, 2023). A later review by Grzybowski et al. (2024) reported that several AI models used to detect diabetic retinopathy have received approval from the US Food and Drug Administration (FDA) and have been implemented in clinical practice. This study, among many others, demonstrates how significant advances in imaging data-trained models are now augmenting or automating the interpretation of imaging data.

Another leading source of data is from EHRs. EHRs are data-rich, containing sources of medical, demographic, and billing data. Jones et al. (2021) investigated AI models using EHR primary care data to facilitate early cancer diagnosis. The results were inconclusive, but the authors endorsed continued work and research with models based on EHR data because of the strong potential to improve patient outcomes. Ruksakulpiwat et al. (2024) systematically reviewed the use of AI in nursing care. They found that EHR data, when paired with ML approaches, can effectively identify patients at high risk for hospital readmission. The review concluded that most models remain at the proof-of-concept or feasibility stage, with limited evidence of widespread, routine use in nursing practice. The authors emphasized the need for prospective validation and real-world implementation studies before these tools can be reliably adopted in daily care. Early identification of sepsis is another growth area using EHR data. Rao et al. (2024) reviewed 16 studies focused on early diagnosis of neonatal sepsis. They found that ML models using EHR and laboratory data consistently outperformed traditional diagnostic methods, with some approaches predicting sepsis hours before clinical recognition and achieving accuracies above 99%. However, most studies relied on retrospective datasets and few offered prospective validation, underscoring the need for further real-world testing. Other empirical research, outside the scope of this umbrella review, indicates that AI tools for the early detection of sepsis in adults have received FDA approval and are already being implemented in clinical practice (Bhargava et al., 2024).

Other data sources have emerged in recent years, such as wearable device data, sensor data, and other health data. Gudigar et al. (2024) investigated the use of AI to detect and assess hypertension. The traditional methods of measuring blood pressure are now being complemented with wearable device data. Wearable devices include smart watches, patch-based monitors, and other devices. These devices allow for continuous remote monitoring that can facilitate earlier detection of hypertension and more proactive management of existing hypertension. In addition, Buijs et al. (2024) investigated the use of scheduling data. They found that historical scheduling data can be used to train models that identify patients at higher risk of missing their appointment. The model can then identify certain time slots that can be overbooked based on this information and prevent outpatient appointment no-shows. Increasingly diverse data types present more opportunities to build AI models that can improve patient care and clinical operations.

### 3.3. RQ3a and b: Application of AI tools in healthcare practice and change over time

The application of AI tools in healthcare practice was extracted during the full-text review phase of the umbrella review. As shown in Figure 8, the distribution of AI healthcare applications, diagnosis, prognosis, and treatment comprised over 80% of the applications in all the reviews. This pattern takes into account that one review may have multiple applications reported. Figure 9 shows the application of AI in healthcare over time. Similarly, the aforementioned three applications comprise a majority of the data types each year. An additional trend is the appearance of other applications in 2023 through 2024, showing that new applications are emerging.

**Fig. 8.**
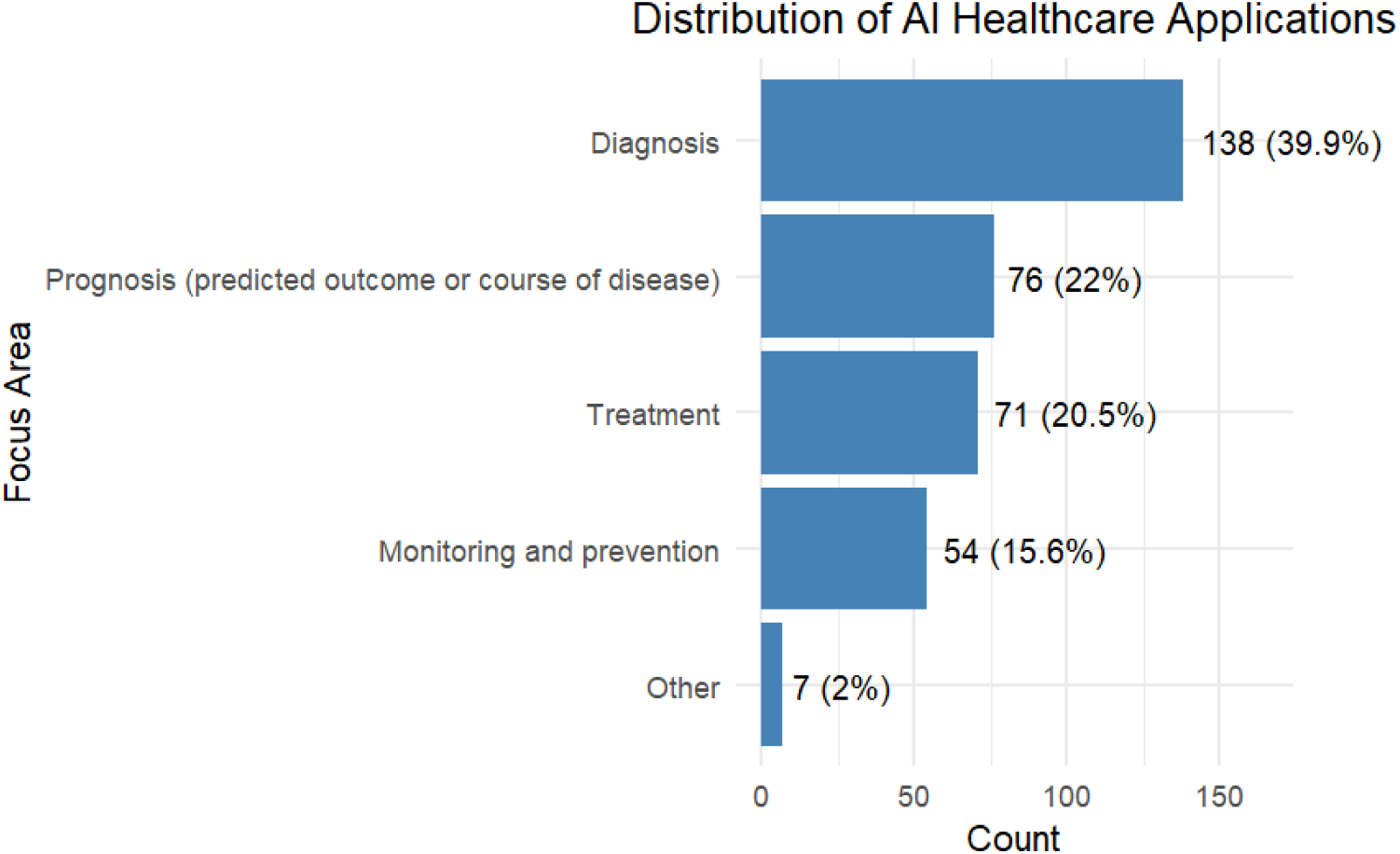
Distribution of AI healthcare applications

**Fig. 9.**
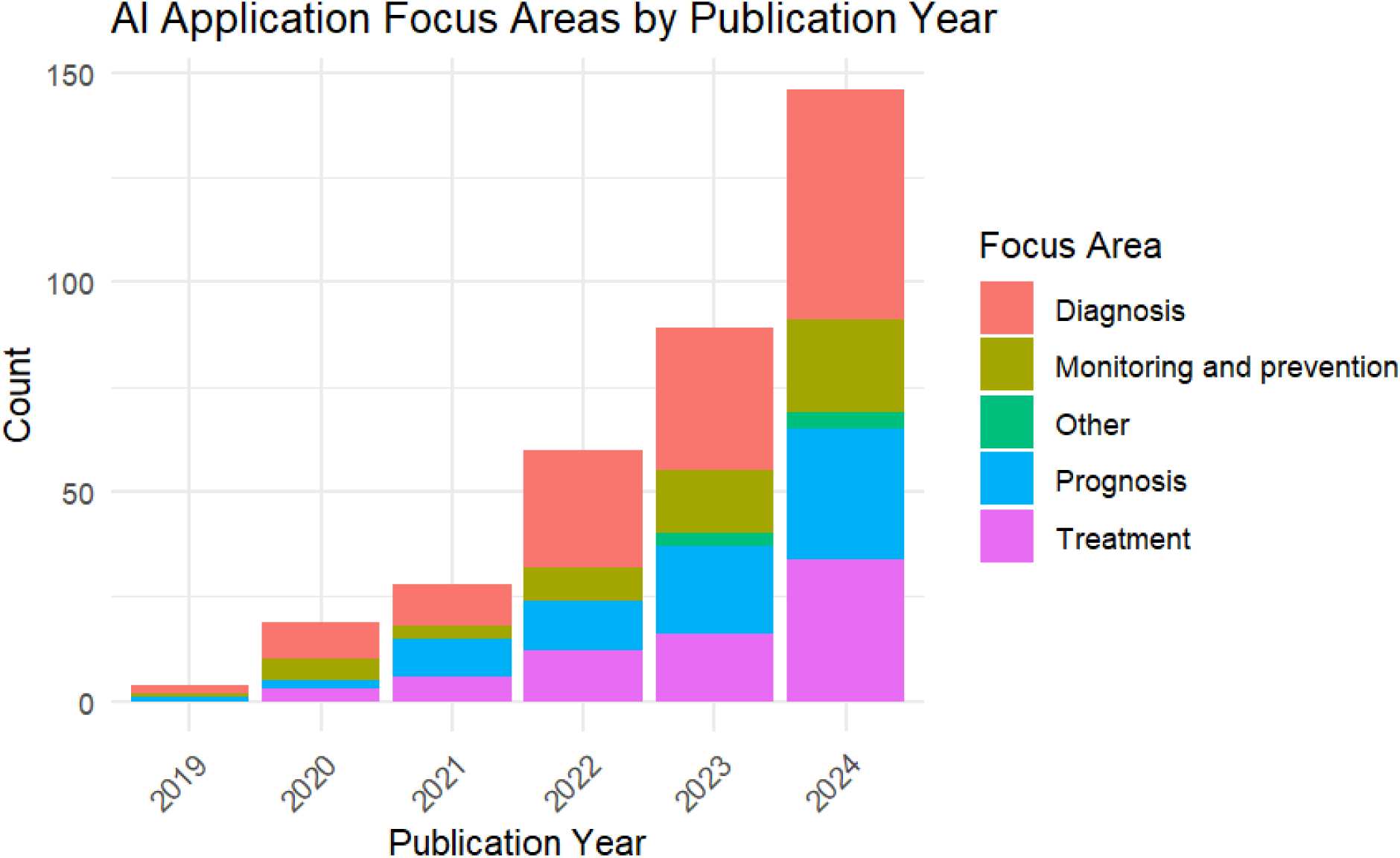
Distribution of AI healthcare applications over time

#### 3.3.1. Narrative review of the application of AI tools in healthcare practice

AI applications in healthcare have rapidly expanded, with diagnosis, prognosis, and treatment dominating the landscape. This trend is reflected consistently across umbrella review data, where these three categories comprise over 80% of applications. Diagnosis, defined as identifying a disease, condition, or injury based on signs and symptoms, was the leading AI application every year (NCI, 2011). For example, AI tools in radiology have demonstrated strong diagnostic capabilities in fracture detection. A meta-analysis by Wong et al. (2024) reported that AI models for diagnosing hand and wrist fracture and dislocation yielded high accuracy, with an Area under the curve (AUC) of 0.946. They recommended prospective external validation to further test the accuracy of these models. Other articles (e.g., Voelker, 2018; Zech et al., 2022), outside the scope of this umbrella review, indicate that fracture detection models have received FDA approval and the models are used to aid clinical decision-making, not replace it.

The second most common AI application was prognosis, which is defined as the predicted outcome or course of a disease, including the chance of recovery or recurrence (NCI, 2011). For example, ML applications for prognosis in lung cancer have demonstrated higher discriminative ability than traditional logistic regression models, with analyses showing improved survival prediction accuracy (Didier, 2024). Ullah et al. (2024) showed that ML had similar discriminative ability in prognosing abdominal aortic aneurysms. The authors of both reviews reported that ML holds promise for more accurate prognosis and enhancing personalized treatment strategies. In addition, both reviews emphasized that further validation using prospective data is required before widespread clinical adoption.

Treatment-based AI models research has increased dramatically in recent years. Obimba et al. (2024) reviewed AI technologies in cancer treatment for older adults, synthesizing evidence across seven studies on lung, breast, and gastrointestinal cancers. The authors found that while AI-assisted approaches such as treatment planning tools (e.g., Watson for Oncology, ChatGPT) show promise, their effectiveness for older adults was generally comparable to or less effective than standard care. Pressman et al. (2024) reported that LLMs have significant potential to assist during the preoperative, intraoperative, and postoperative phases of surgical care, including risk assessment, patient education, and real-time surgical guidance. Other empirical research, outside the scope of this umbrella review, indicates that an AI-powered surgical robot and platform has recently received FDA approval and has been implemented in clinical practice (Cadière et al., 2024).

AI model research focused on monitoring and prevention has also increased in recent years. For example, Moffat and Xu (2022) compared the performance of ML models with the Modified Early Warning Score (MEWS) to monitor for risk of in-hospital cardiac arrest (IHCA). The MEWS score is a risk assessment tool used to identify patients who may need an increased level of care. Utilizing MEWS can allow for earlier detection or prevention of adverse medical events such as IHCA. The results showed ML models had similar or superior performance compared to MEWS; however, variability in study design and data sources limits generalizability. Leghissa et al. (2023) investigated ML models’ ability to detect or predict frailty in elderly people. Earlier detection or prediction of frailty can lead to more effective treatment and more optimal allocation of resources. The results were inconclusive due to varying definitions of frailty and data sources to build models.

Beyond traditional clinical applications, new domains are emerging. For example, Salman et al. (2023) investigated the use of AI models to predict total knee arthroplasty implant component sizes. The authors reported moderate to high accuracy in the review. Sacoransky et al. (2024) investigated the use of LLMs to generate structured radiology reports. The authors concluded that LLMs demonstrate the capability to generate reports and save radiologists’ time and resources. Overall, the use of LLMs to assist in clinical documentation has advanced significantly in the past two years. New applications of AI tools are emerging in healthcare practice, and further growth of this trend is possible moving forward.

### 3.4. RQ4a and b: Ethical concerns and change over time

We extracted the ethical concerns reported in the results, discussion, or conclusion of the reviews. Ethical concerns include data privacy, model accuracy, data and algorithm bias, and explainability. Figure 10 shows that the majority of reviews report ethical concerns (64.6% Yes and 35.4% No). The count of ethical concerns reported increased each year from 2021 through 2024(Figure 11). The increasing proportion of reviews reporting ethical concerns suggests growing awareness and perceived challenges.

**Fig. 10.**
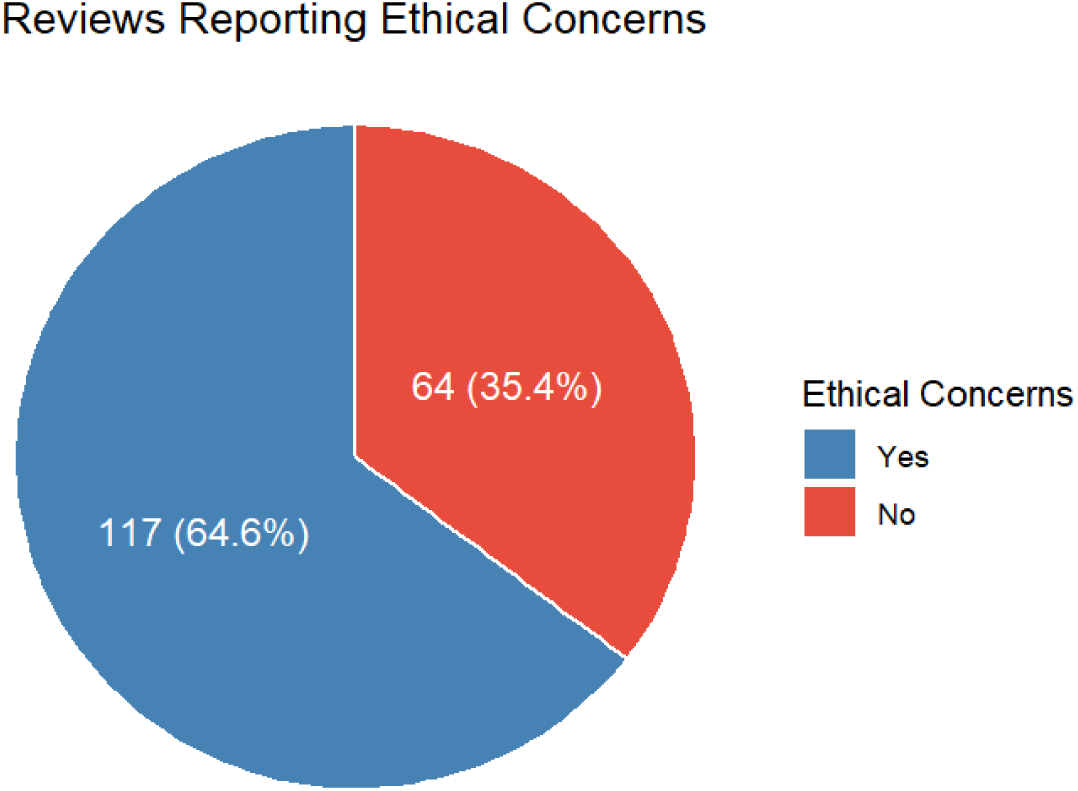
Ethical concerns reported by the review authors

**Fig. 11.**
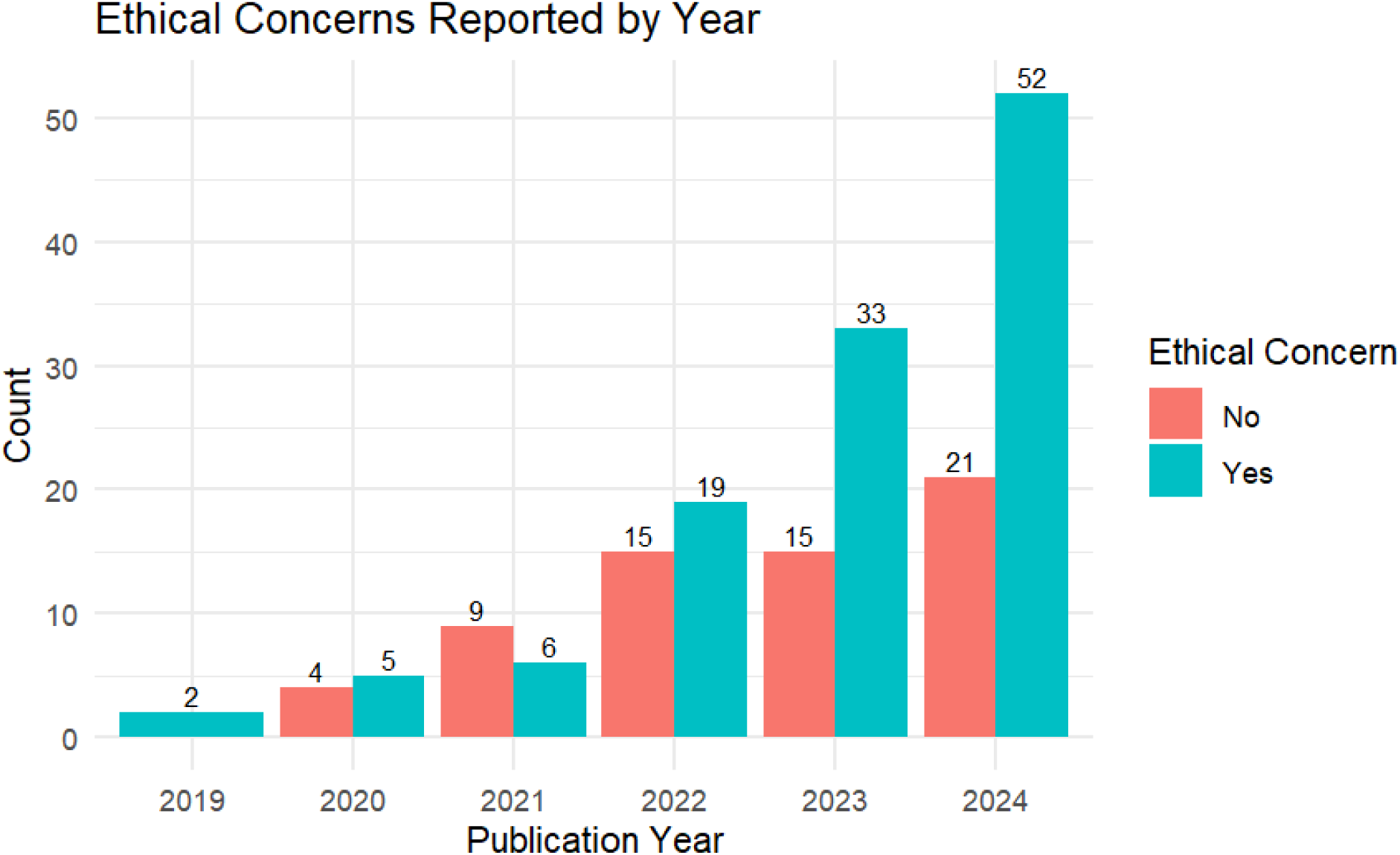
Ethical concerns reported by the review authors over time

#### 3.4.1. Narrative review of ethical concerns

Ethical concerns in the reviews ranged from data privacy issues, model accuracy, data and algorithmic bias, and explainability concerns. Each of these ethical concerns has the potential to compromise patient safety.

##### Data Privacy

AI models require extensive use of healthcare data. In the United States, some of this data may be protected by legislation such as the Health Insurance Portability and Accountability Act (HIPAA) (HHS, 2021). In the review of Kitsios et al. (2023), the authors emphasized that healthcare organizations should ensure that protocols and protections are in place to gain consent for and secure the collection and use of data to train models. Ali et al. (2023) discussed concerns about protecting patient data that is fed into the model once it is implemented. The risk of a data breach is a serious threat that has unfortunately become commonplace. The authors emphasized the importance of regulation and safeguards to ensure that health data is secure.

##### Model Accuracy

In their review of early cancer diagnosis using primary care data, Jones et al. (2021) recommended that prospective data is needed to further validate AI models for implementation in practice, in addition to the retrospective data. Plana et al. (2022) also recommended more rigorous validation in their review. Specifically, they suggested that more randomized clinical trials (RCTs) are needed to further test the accuracy of AI models, given that the FDA has approved 343 AI applications, but they found that only 41 RCTs have been reported. Xavier et al. (2024) suggested that algorithm performance should be available for healthcare professionals and leaders to make informed decisions regarding which models are the most accurate. This is especially relevant for LLMs given their tendency to hallucinate or generate false content. Veneziani et al. (2024) reported additional concerns about model accuracy. Specifically, the algorithms to interpret subtle emotional and behavioral cues are limited during patient evaluations. The authors recommended that clinicians must find a balance between integrating AI technology without sacrificing the human connection and reasoning that are essential to clinical decision making.

##### Data and Algorithmic Bias

In their review of AI applications used in the detection, management, and prognosis of bone metastases, Papalia et al. (2024) reported that ensuring diverse data sets representing different patient populations is essential for safe adoption, because the diverse data allows for generalizability and widespread implementation. Radaelli et al. (2024) also reported that the effectiveness of AI models is limited by the quality of data used to train the models. For example, if a dataset does not contain certain variables, e.g.,blood culture data to predict infections, this missing data may lead to bias and limit the generalizability of the model. In their review of AI applications for clinical decision support in acute ischemic stroke, Akay et al. (2023) emphasized the need for high-quality training and validation data. Biased or incomplete data could lead to flawed predictions and potential patient harm. They recommended policy requirements for data used to build and validate AI models.

##### Explainability

Javidan et al. (2022) reported concerns about the “black box” dilemma that algorithms present. That is, many AI applications lack explainability. Explainability is especially challenging with deep learning models, which often make it difficult to interpret how predictors contribute to a specific output. As AI models become more advanced and complex, explainability decreases about how an output is generated. The decrease in explainability can make decision-making more challenging than ever.

## 4. Discussion

### 4.1. RQ1: Characteristics of AI applications in healthcare practice

The steady increase in the number of systematic reviews from 2019 to 2024 highlights the accelerating interest in and adoption of AI applications in healthcare. While earlier years (2019– 2020) may have smaller sample sizes due to the emerging nature of this research field, the peak in 2024 suggests that AI has transitioned from a niche focus to a mainstream research priority. This trend mirrors broader trends in AI adoption across industries, particularly as healthcare organizations seek solutions for diagnostic efficiency, predictive modeling, and operational improvement.

Geographically, the dominance of first authors from the United States, United Kingdom, Australia, India, China, Italy, and Saudi Arabia reflects both the concentration of AI research infrastructure and the policy environments that support innovation in these countries. The breadth of 133 journals publishing on this topic underscores the interdisciplinary nature of AI in healthcare and indicates that the research community is distributed across clinical, technical, and policy-oriented outlets.

The network effect observed between medical specialties, diseases/conditions, and other themes suggests that certain domains (e.g., radiology, critical care, cancer) act as “AI research hubs” where tool development and validation are more advanced. Underrepresented areas, such as geriatric and gynecologic disease, may represent untapped opportunities. Targeting these gaps could broaden the impact of AI in healthcare beyond its current strongholds.

### 4.2. RQ2a and b: Healthcare data types used to train AI models and change over time

Diagnostic imaging, EHR data, and biomarkers/laboratory results consistently dominate as training data sources, accounting for about 70% of all data types across reviews. This dominance reflects their availability, structure, and established role in clinical decision-making. Kolasa et al. (2024) reported a similar finding in their umbrella review, with the three data types accounting for 75% of all data types. Imaging data, in particular, benefits from decades of digitization, making it ideal for deep learning applications. EHR data offers breadth, capturing demographic, clinical, and billing information, but challenges remain in standardization, interoperability, and data quality.

The emergence of wearable device data, sensor data, and other nontraditional sources from 2022 onward represents a shift toward continuous, real-world data capture. Emerging data types, such as AI-assisted hypertension detection and predictive models for appointment no-shows, illustrate the expanding scope of what constitutes “healthcare data.” The expansion in healthcare data could enable more proactive, precise, convenient, and personalized healthcare, because it integrates continuous, real-world physiological data rather than only relying on traditional health data. However, the new data types also raise issues of integration, privacy, regulatory oversight, access to devices, and bias mitigation.

### 4.3. RQ3a and b: Application of AI tools in healthcare practice and change over time

Diagnosis, prognosis, and treatment remain the dominant AI applications, which align with the clinical imperative to improve patient outcomes through better detection, risk stratification, and intervention planning. The high accuracy reported for diagnostic tasks, such as fracture detection and diabetic retinopathy screening, demonstrates AI’s potential to aid or partially automate specialized tasks. Prognostic models, particularly in critical care settings, show promise for enabling earlier, targeted interventions. Using AI for treatment has increased every year but remains at an early stage, with research focusing on more retrospective data. However, promising advances are happening with surgical care and AI-powered robotics.

The appearance of newer applications in 2023-2024, such as LLMs for documentation and AI-assisted surgical planning, suggests that AI’s role is expanding beyond traditional clinical endpoints. These applications could have downstream benefits in workflow efficiency, patient communication, and cost containment. Consequently, these applications can significantly reduce administrative burden for clinicians, allow more time for direct patient interaction, and improve the quality of patient records for subsequent care. However, the translation of novel applications into routine practice will depend on robust validation, regulatory approval, and integration into existing clinical workflows.

### 4.4. RQ4a and b: Ethical concerns and change over time

The upward trend in both the number and proportion of reviews citing ethical concerns reflects growing awareness of the risks and unintended consequences of AI in healthcare. The most frequently cited issues include data privacy, model inaccuracy, data and algorithmic bias, and explainability, which align with themes in broader AI ethics discourse. The “black box” challenge or the lack of explainability, in particular, underscores tension between accuracy and interpretability, especially for deep learning models.

The persistence of these concerns suggests that technical advances alone will be insufficient. The H-AI-H model of human + AI integration is essential to ensure ethical oversight. Progress in governance, regulation, and stakeholder engagement is essential. Strategies such as bias audits, explainable AI methods, and benchmarking standards for validation could help address these concerns. Moreover, ethical considerations are likely to become more complex as data sources diversify and applications expand, making interdisciplinary collaboration critical between clinicians, data scientists, ethicists, and policymakers. Overall, further research is warranted to ensure safety and earn the trust of regulators, clinicians, and patients.

## 5. Conclusions and Limitations

### 5.1. Major Findings

This umbrella review makes several key contributions to the understanding of AI applications in healthcare practice. First, we synthesized evidence from 181 systematic reviews to characterize AI in healthcare across multiple dimensions, including publication trends, geographic distribution, thematic focus, and medical specialties. We identified high-density research areas, such as radiology, oncology, and critical care, alongside underrepresented specialties and conditions, highlighting opportunities for future investigation.

Second, we analyzed the types of healthcare data used to train AI models. Diagnostic imaging, EHR data, and biomarkers/laboratory results accounted for approximately 70% of data sources, reflecting their accessibility and clinical relevance. However, emerging sources such as wearable device and sensor data indicate a shift toward continuous, real-world data capture, with implications for personalized care and operational efficiency.

Third, we examined AI applications in healthcare practice and found that diagnosis, prognosis, and treatment dominate the current landscape. Reviews in later years showed the emergence of novel applications, including the use of LLMs for clinical documentation and AI tools for operational decision-making.

Fourth, we documented an upward trend in reported ethical concerns, with bias, privacy, safety, transparency, and explainability emerging as recurring themes. The growing prevalence of such concerns suggests an increasing awareness of the governance, regulatory, and trust-building requirements for successful AI adoption in healthcare.

Collectively, these findings provide a panoramic view of the AI healthcare literature and can help researchers, clinicians, and policymakers identify both the existing AI applications in healthcare practices and critical gaps that warrant further exploration.

### 5.2 Limitations and Implications

Despite efforts to ensure a comprehensive synthesis, several limitations should be acknowledged. First, while our search spanned multiple major academic databases and was updated through November 2024, relevant systematic reviews may have been missed if they were indexed outside these sources, published after our cutoff date, written in languages other than English, or reported in the grey literature. As AI in healthcare is evolving rapidly, future umbrella reviews should incorporate incoming publications to capture new developments.

Second, we focused exclusively on systematic reviews that reported applications of AI in healthcare practice. Reviews that examined AI algorithms without discussing their clinical or operational applications were excluded. Although this exclusion aligns with our practice-oriented scope, reviews on AI technical development merit separate synthesis to provide complementary insights into algorithmic innovation.

Third, the heterogeneity of included reviews, spanning diverse clinical specialties, data types, and evaluation methods, limited our ability to perform pooled quantitative analyses. Our synthesis relied on reported findings, which may be influenced by varying review quality, incomplete reporting, and differing definitions of AI. While we described thematic and temporal trends, the absence of standardized metrics across reviews constrains direct comparisons among these reviews.

Finally, our scope excluded AI applications in adjacent domains such as public health, healthcare research, healthcare administration, and health professions education. These areas may represent significant opportunities for AI-driven applications and should be explored in future reviews. In sum, this umbrella review underscores the rapid expansion, concentration, and diversification of AI applications in healthcare. Meanwhile, it also draws attention to the ethical, methodological, and policy challenges that must be addressed to successfully integrate AI into healthcare practice and improve health outcomes and efficiency.

## Data Availability

All data produced in the present study are available upon reasonable request to the authors.

## Statement on open data and ethics

The data supporting the findings of this study are available from the corresponding author upon reasonable request. As this study is an umbrella review, it does not report or involve the use of any animal or human data and therefore did not require ethics approval or informed consent.

## CRediT authorship contribution statement

**Adam Andersen**: Conceptualization, Data curation, Formal analysis, Methodology, Coding for data traction and quality assessment, Project administration, Software, Validation, Visualization, Writing –original draft, Writing –review & editing, Managing references.

**Ruiping Huang**: Conceptualization, Writing –review & editing, Coding for data extraction and quality assessment, Software, Validation, Project administration.

**Edward Jiusi Liu**: Writing –review & editing, Coding for data extraction and quality assessment.

## Acknowledgments

The authors would like to acknowledge the various contributions made by Dr. Yue Yin during the preparation of this paper.

## Declaration of generative AI in scientific writing

During the preparation of this work, the authors used ChatGPT to proofread grammar. After using this tool/service, the authors reviewed and edited the content as needed and take full responsibility for the content of the publication.

## Appendix A. The systematic reviews included in this umbrella review

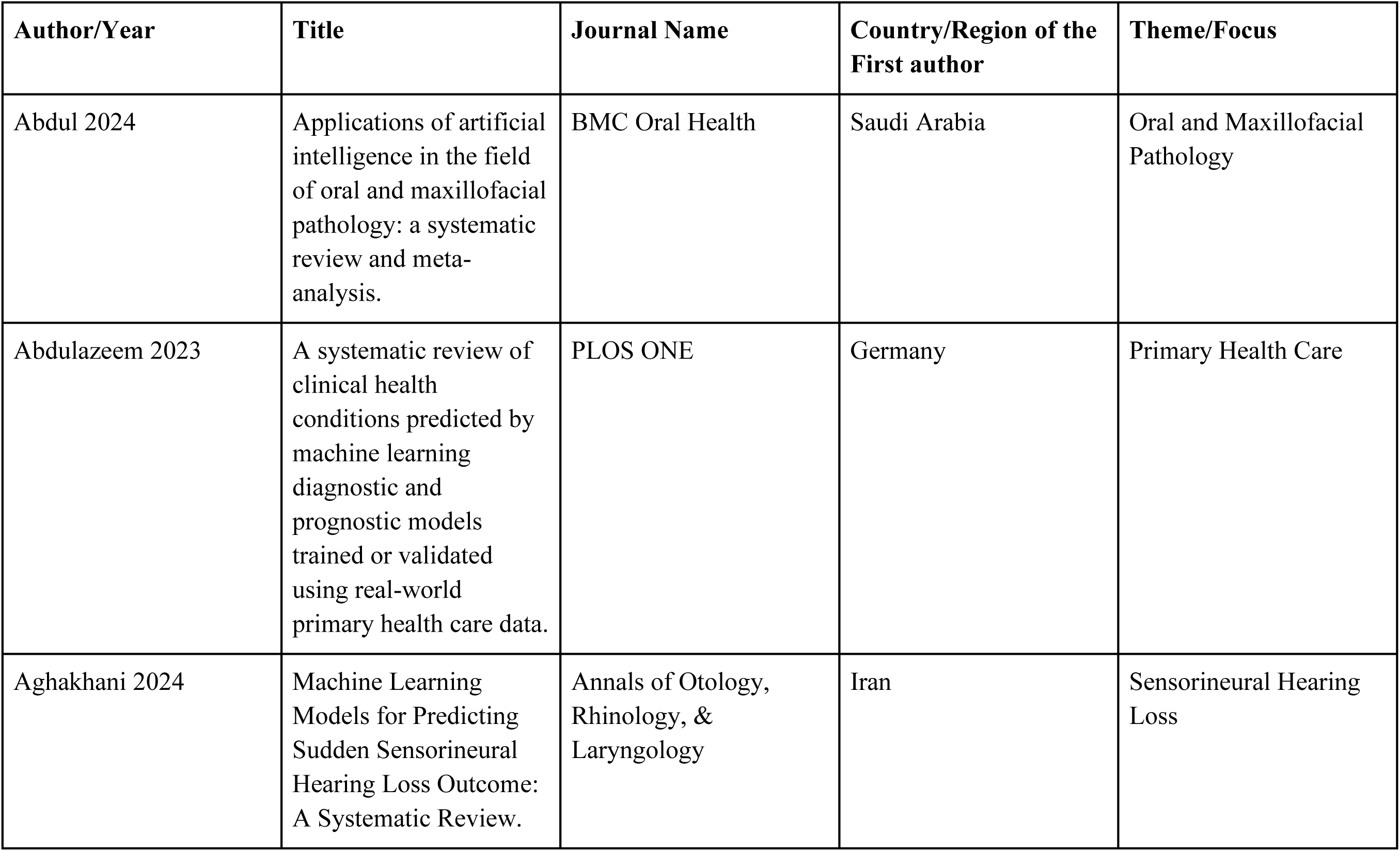
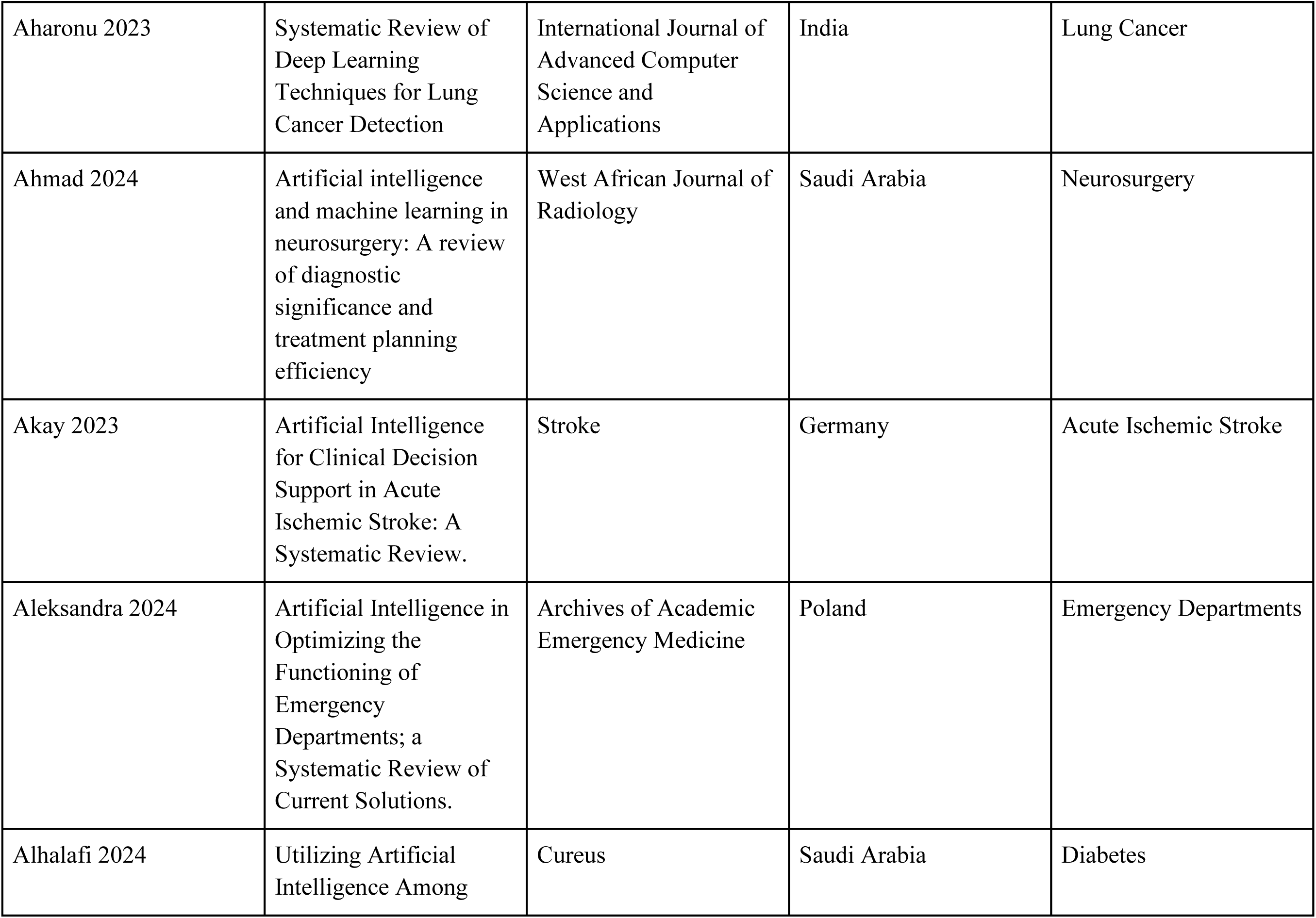
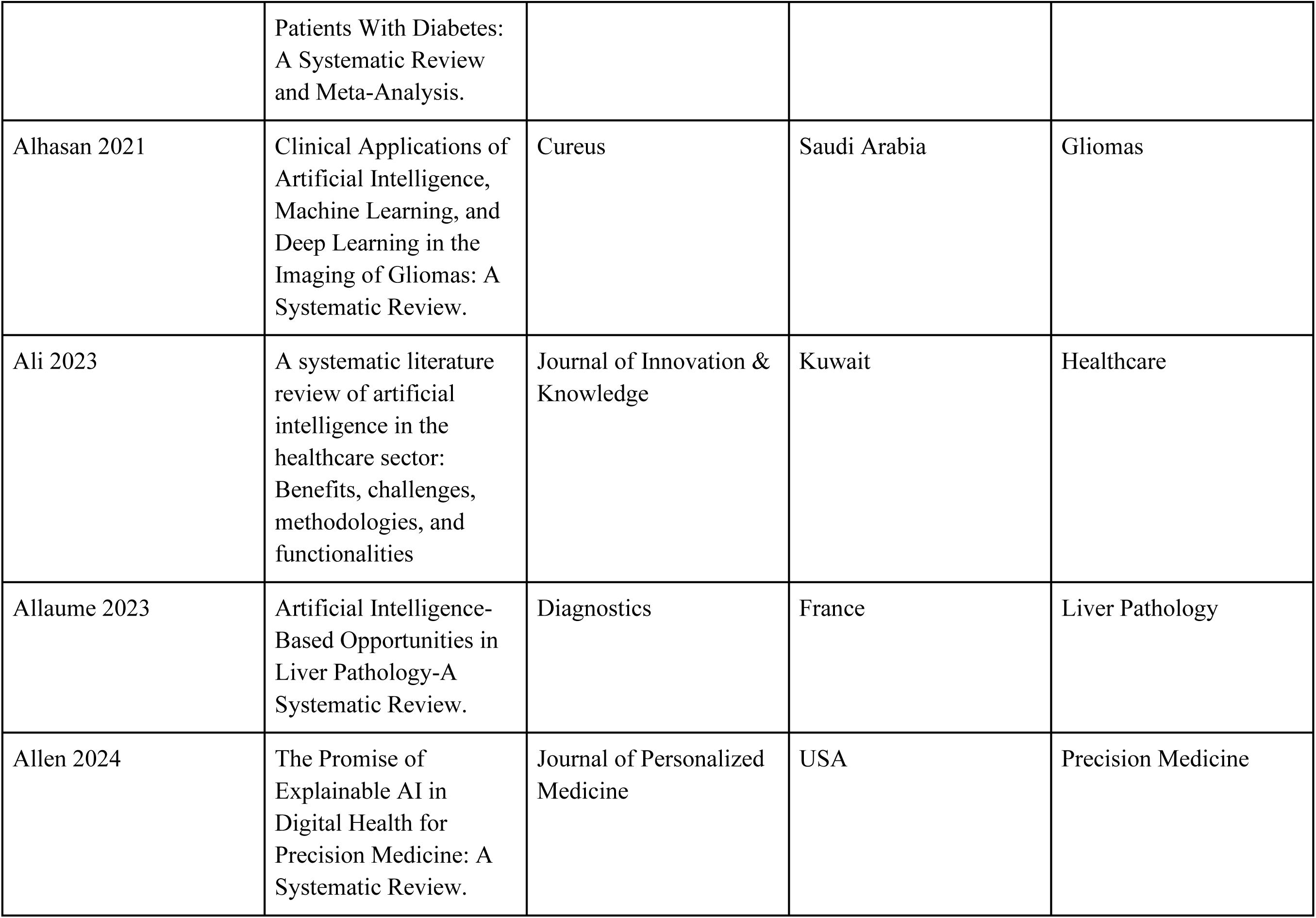
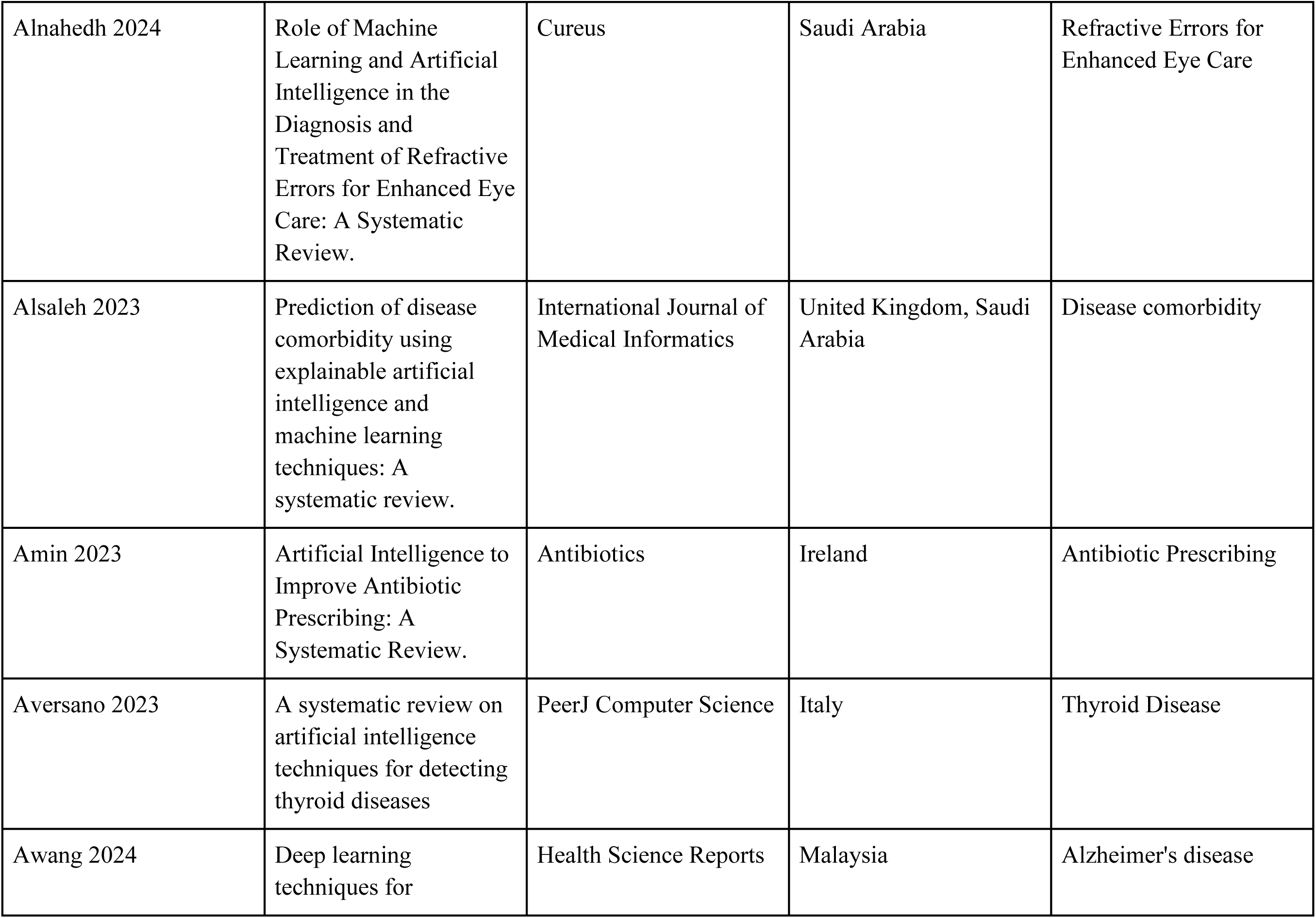
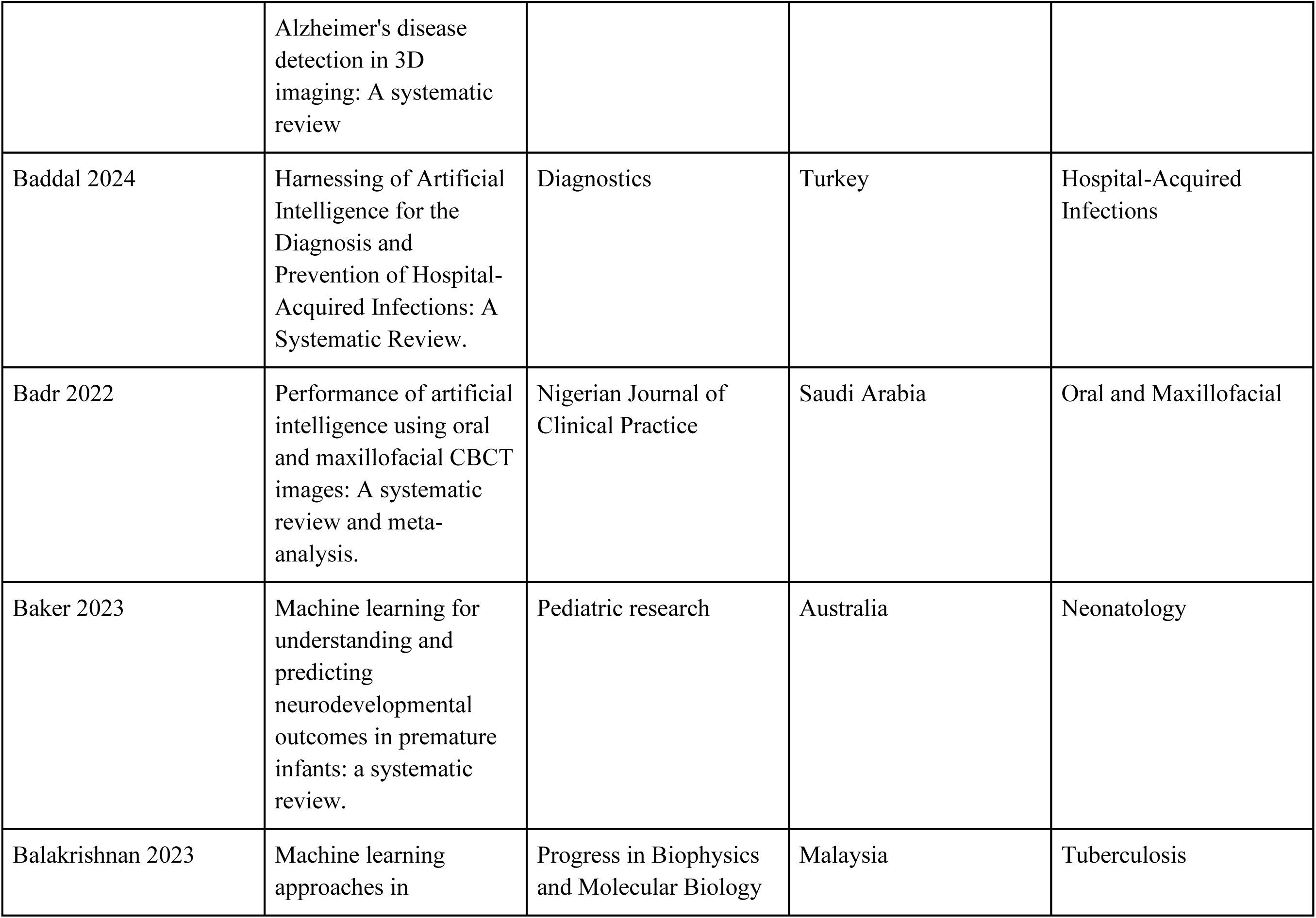
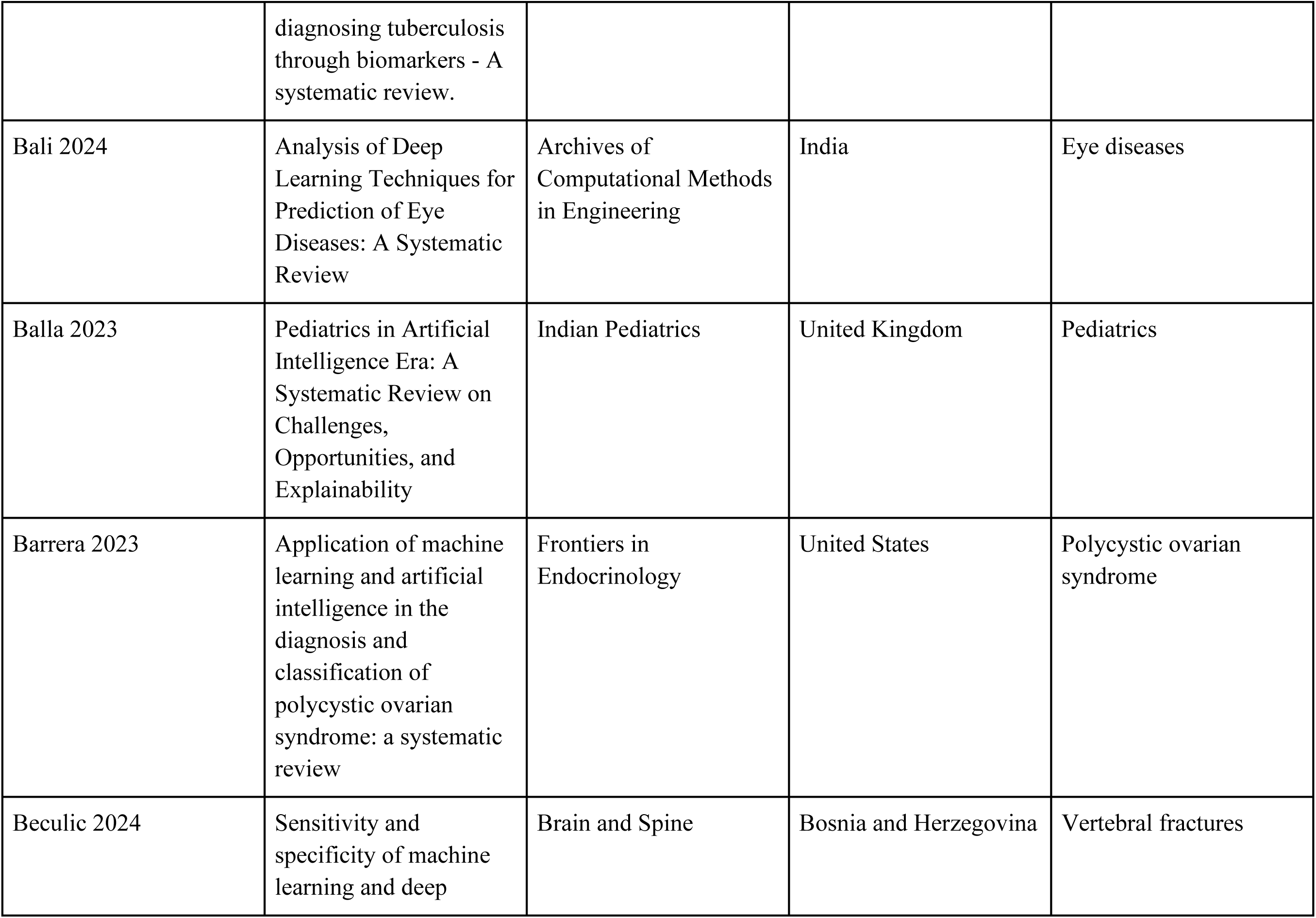
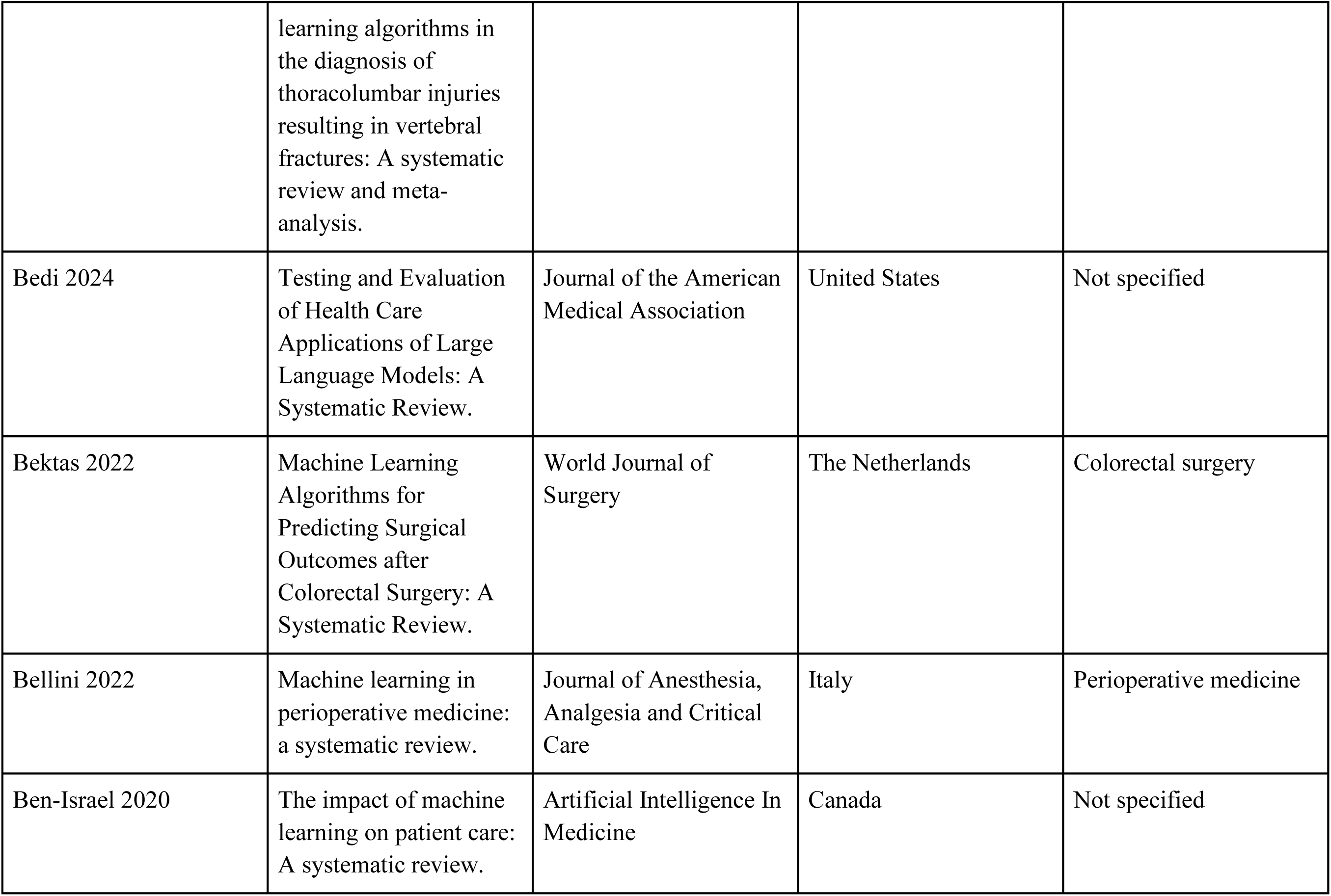
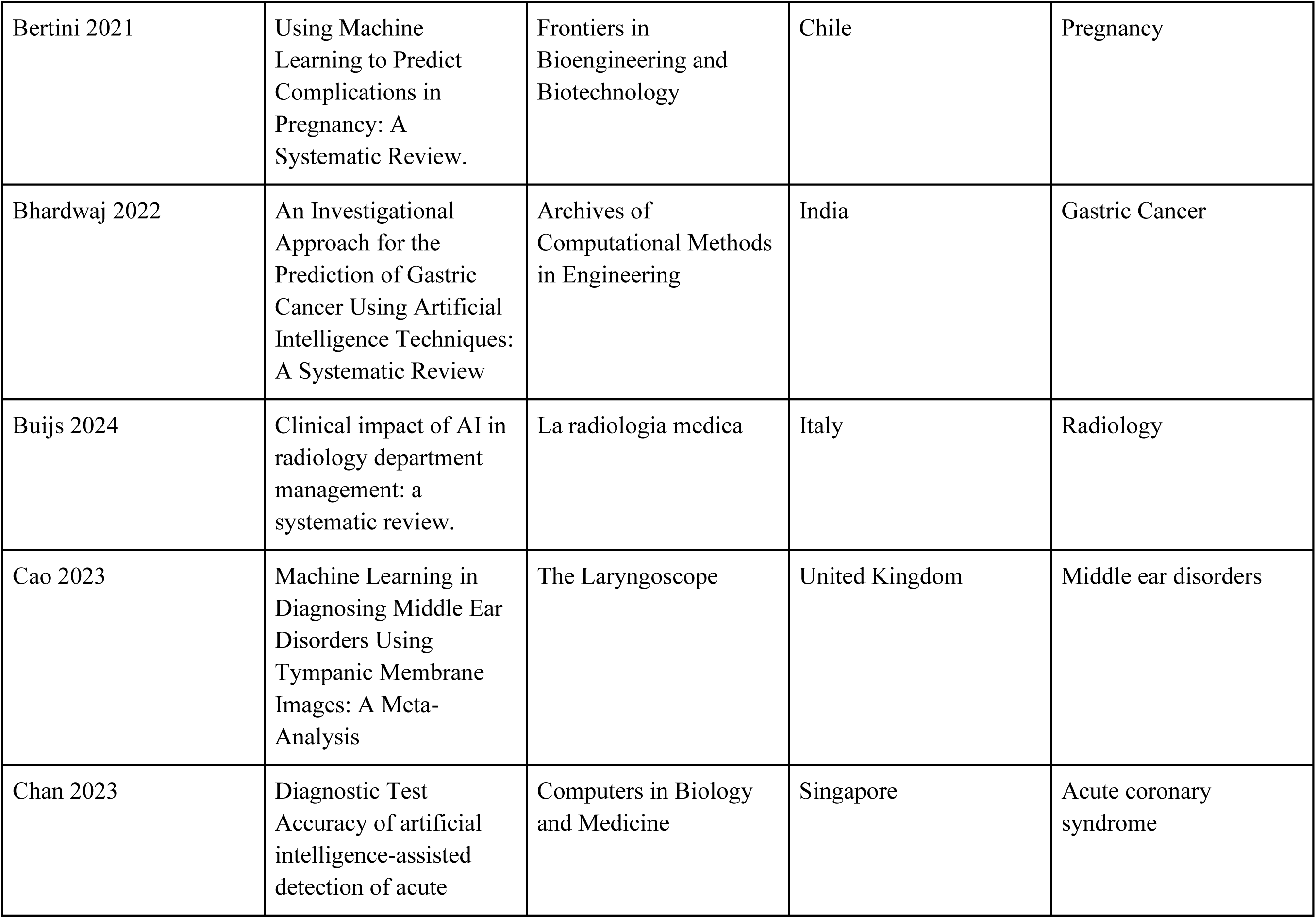
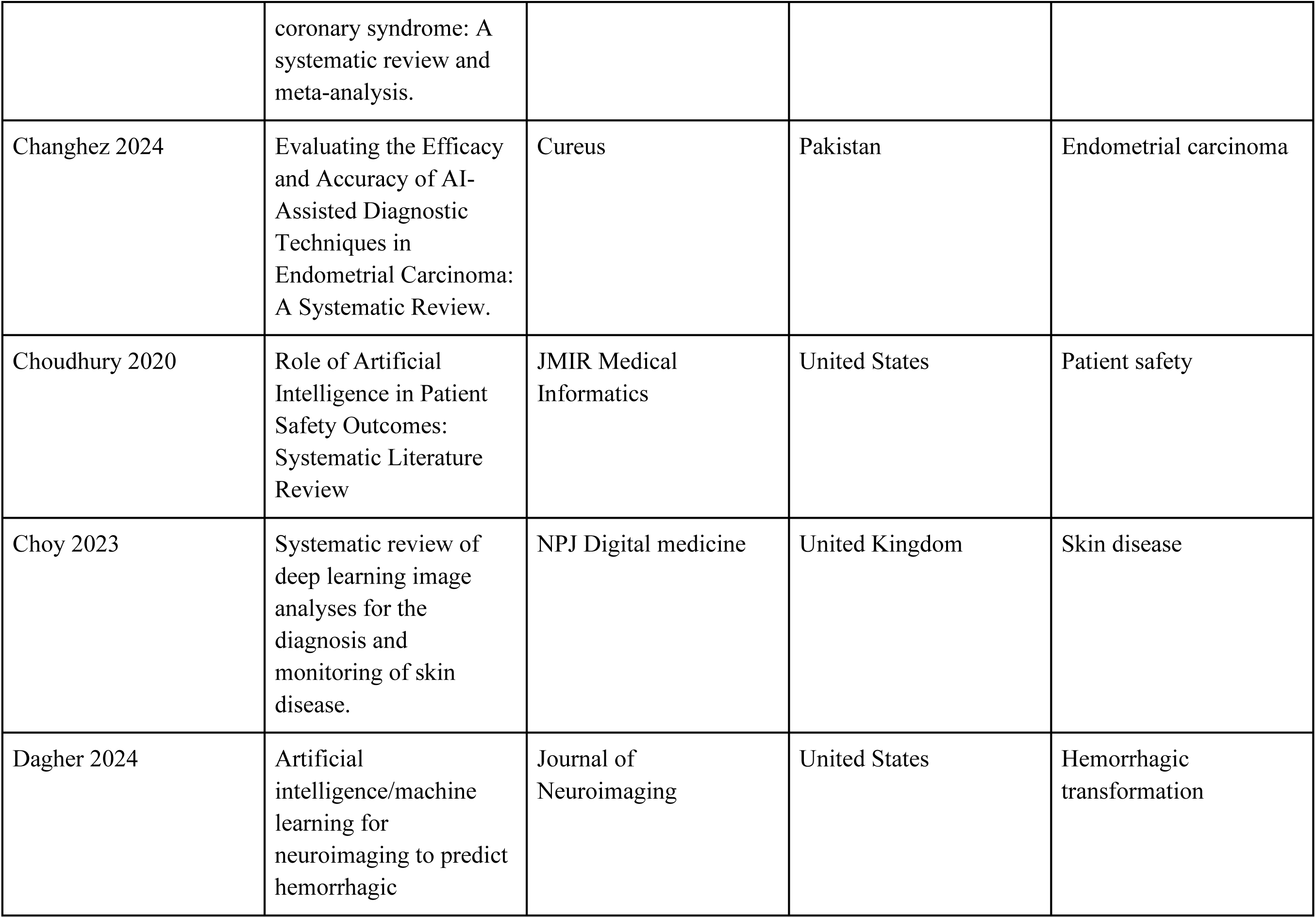
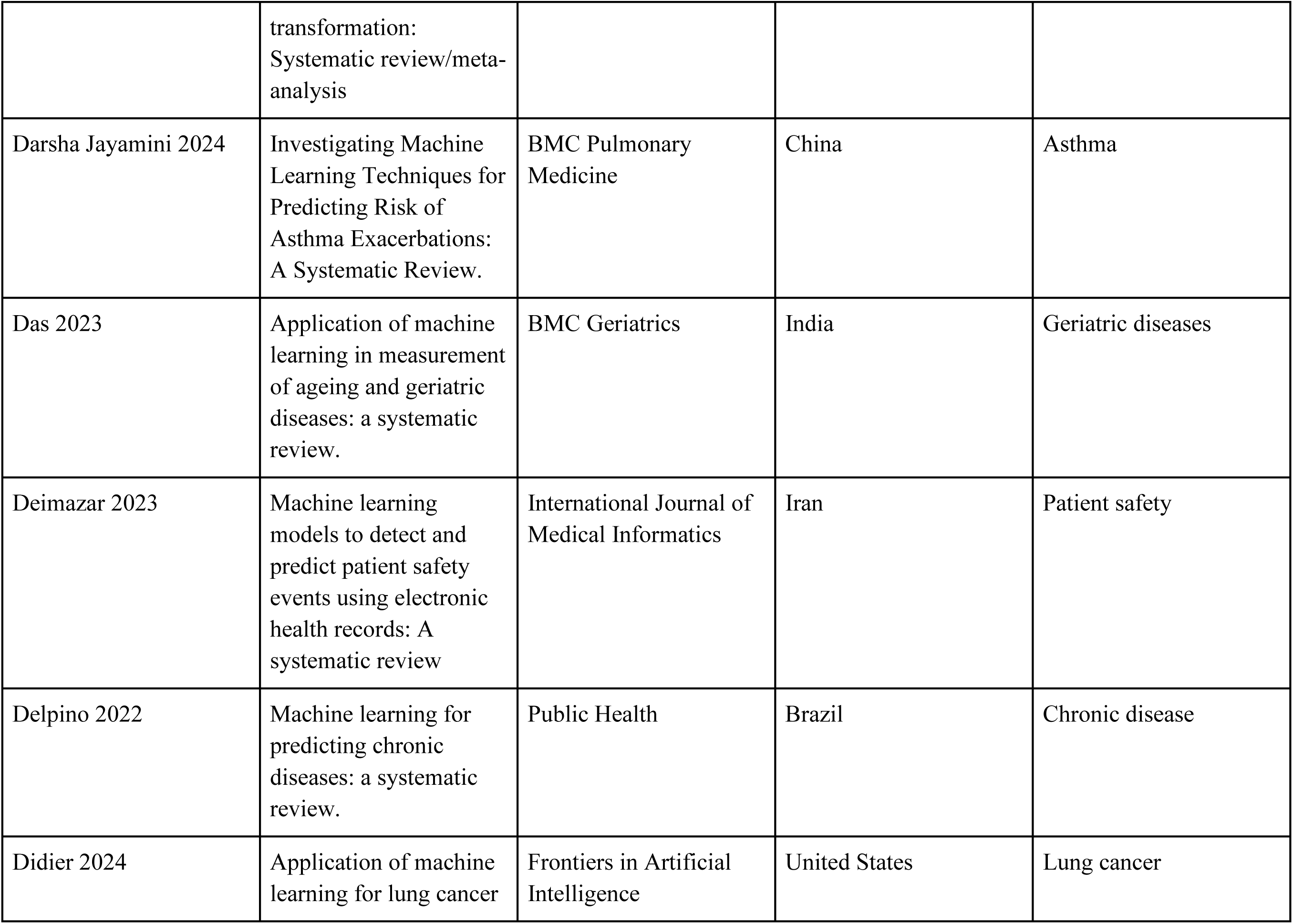
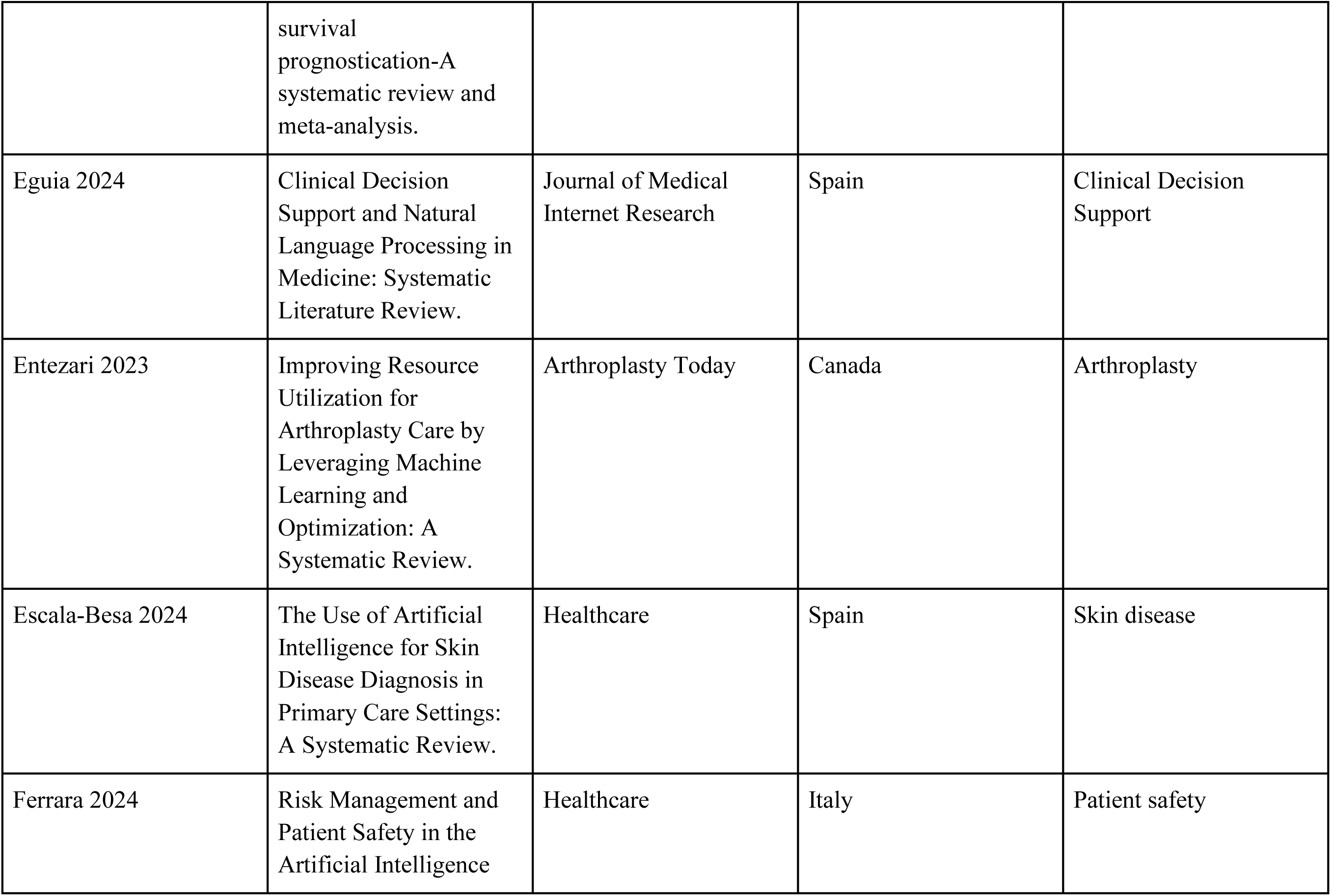
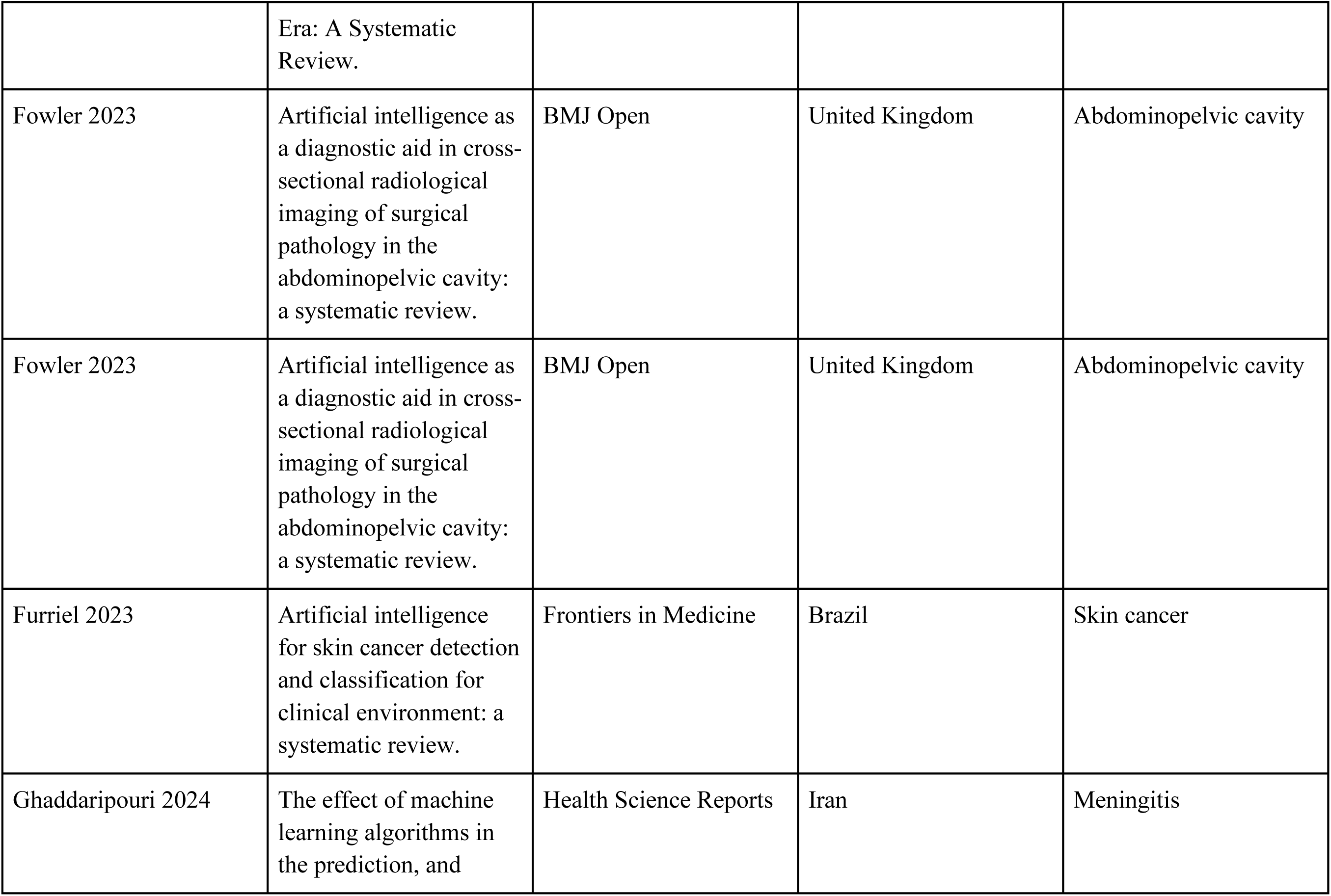
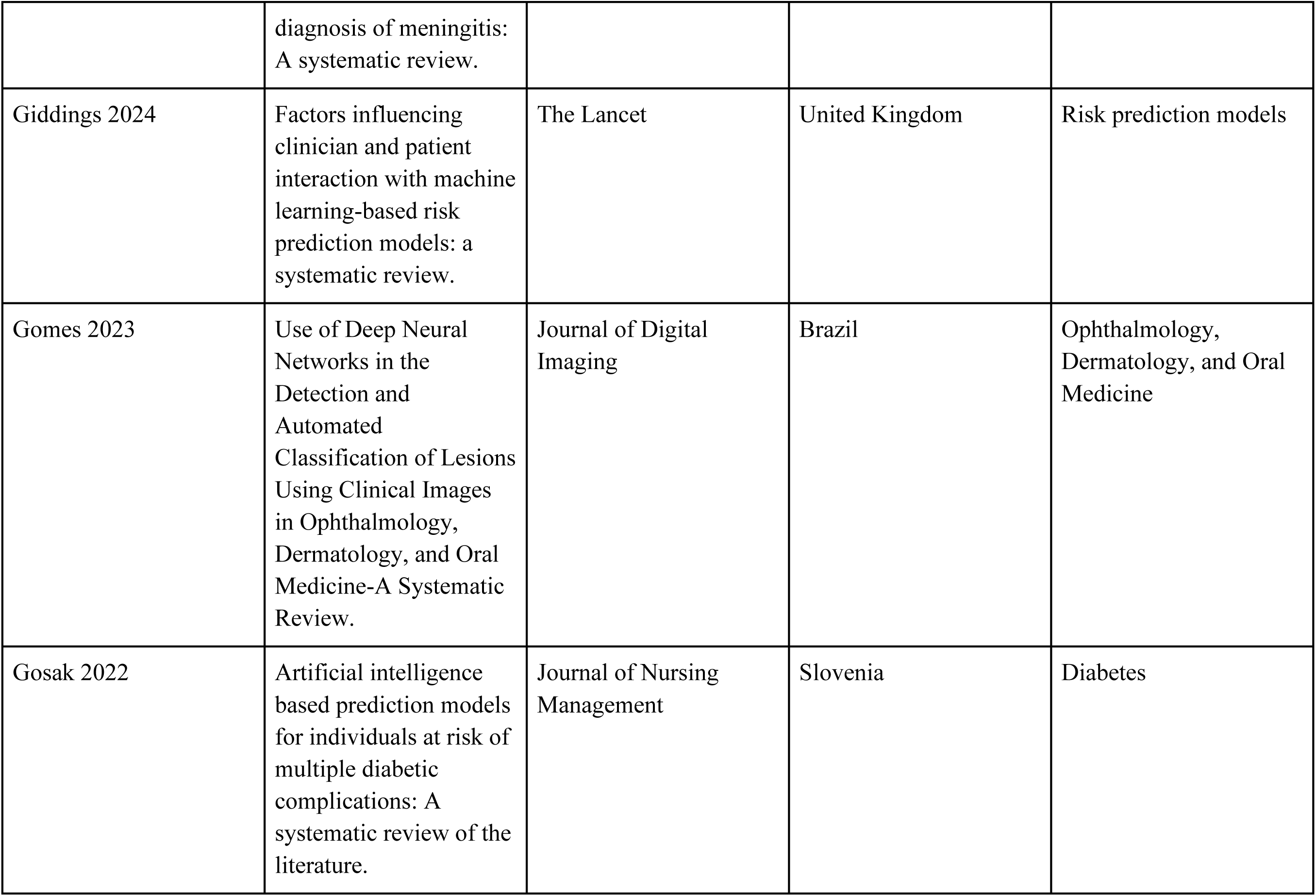
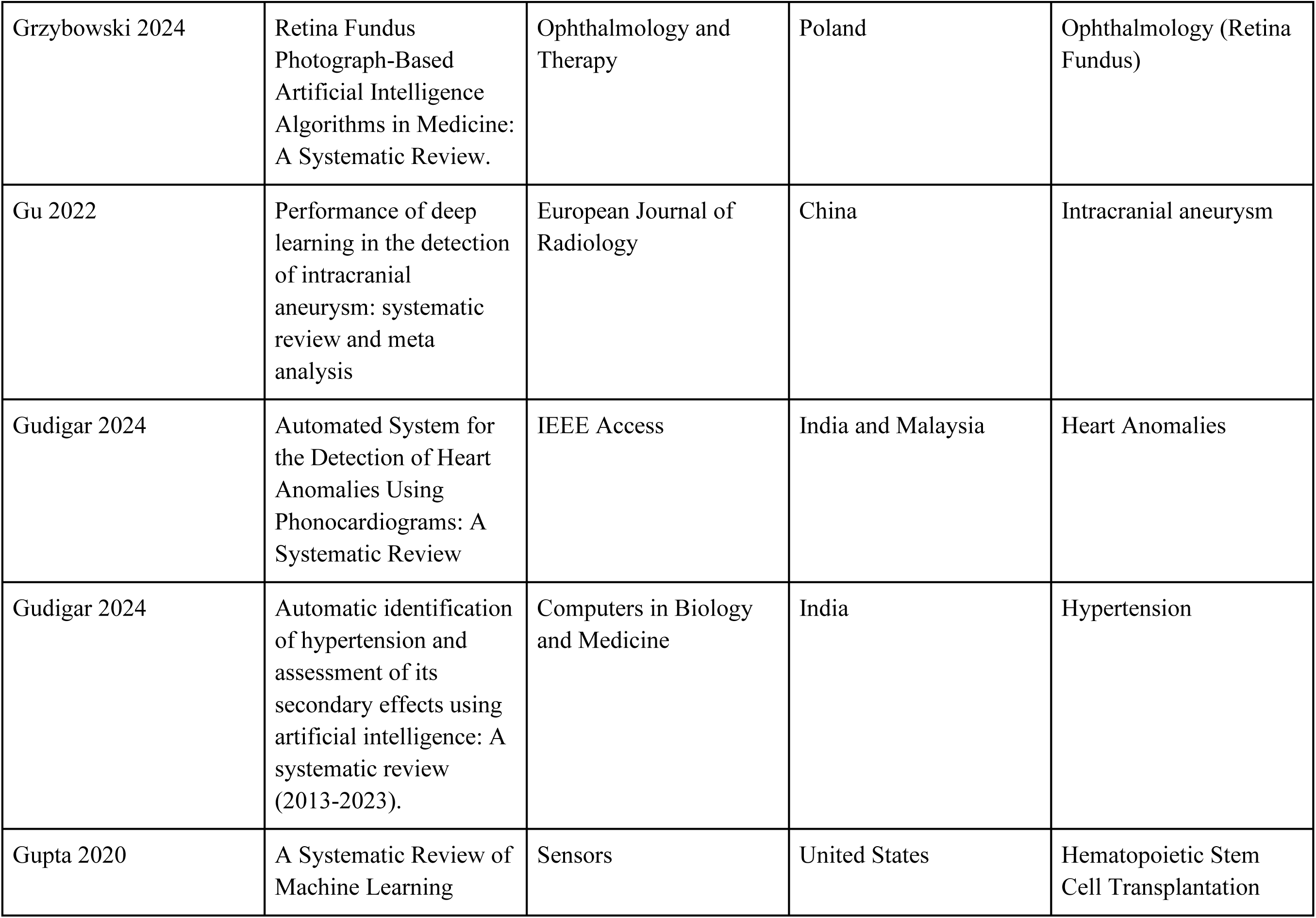
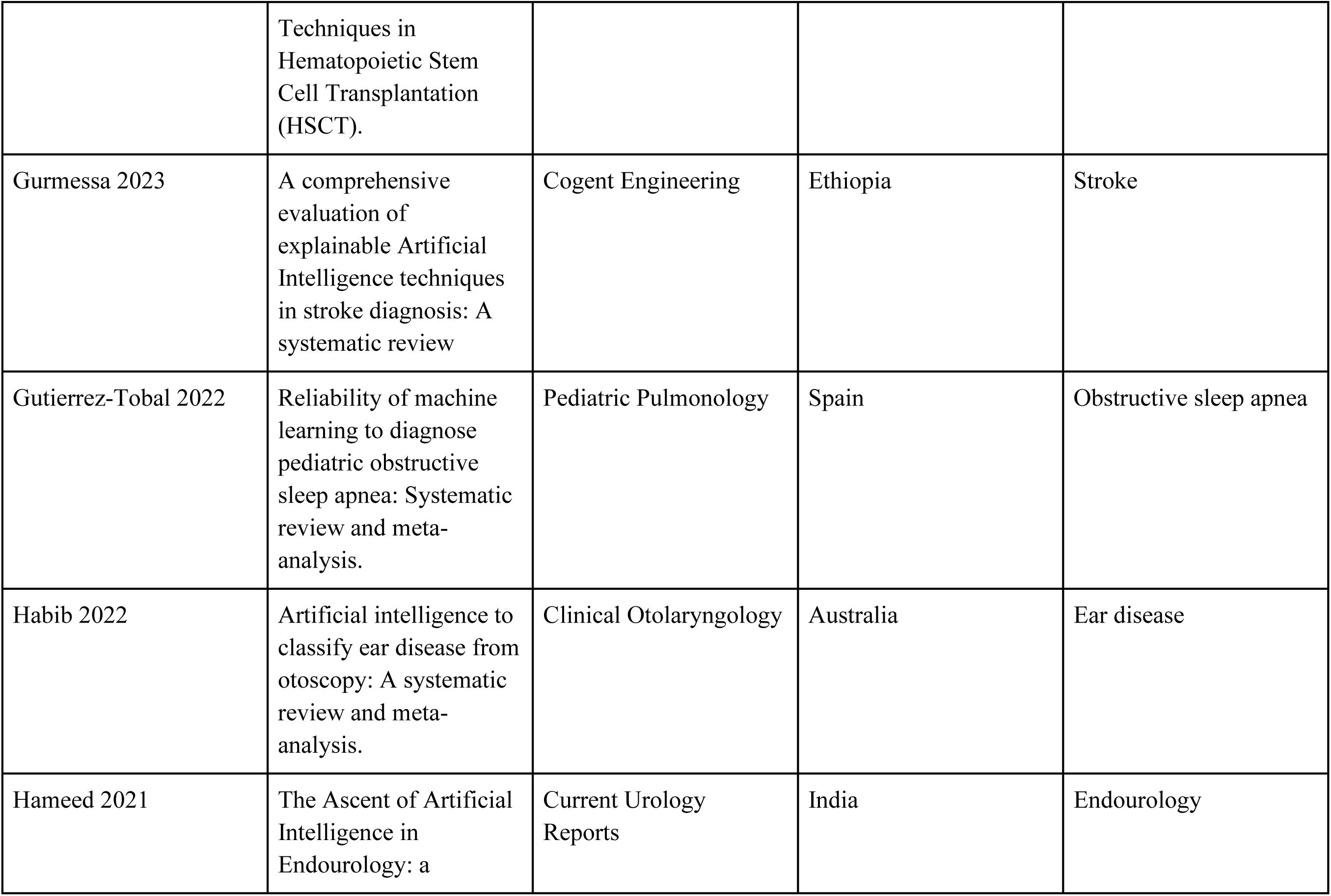
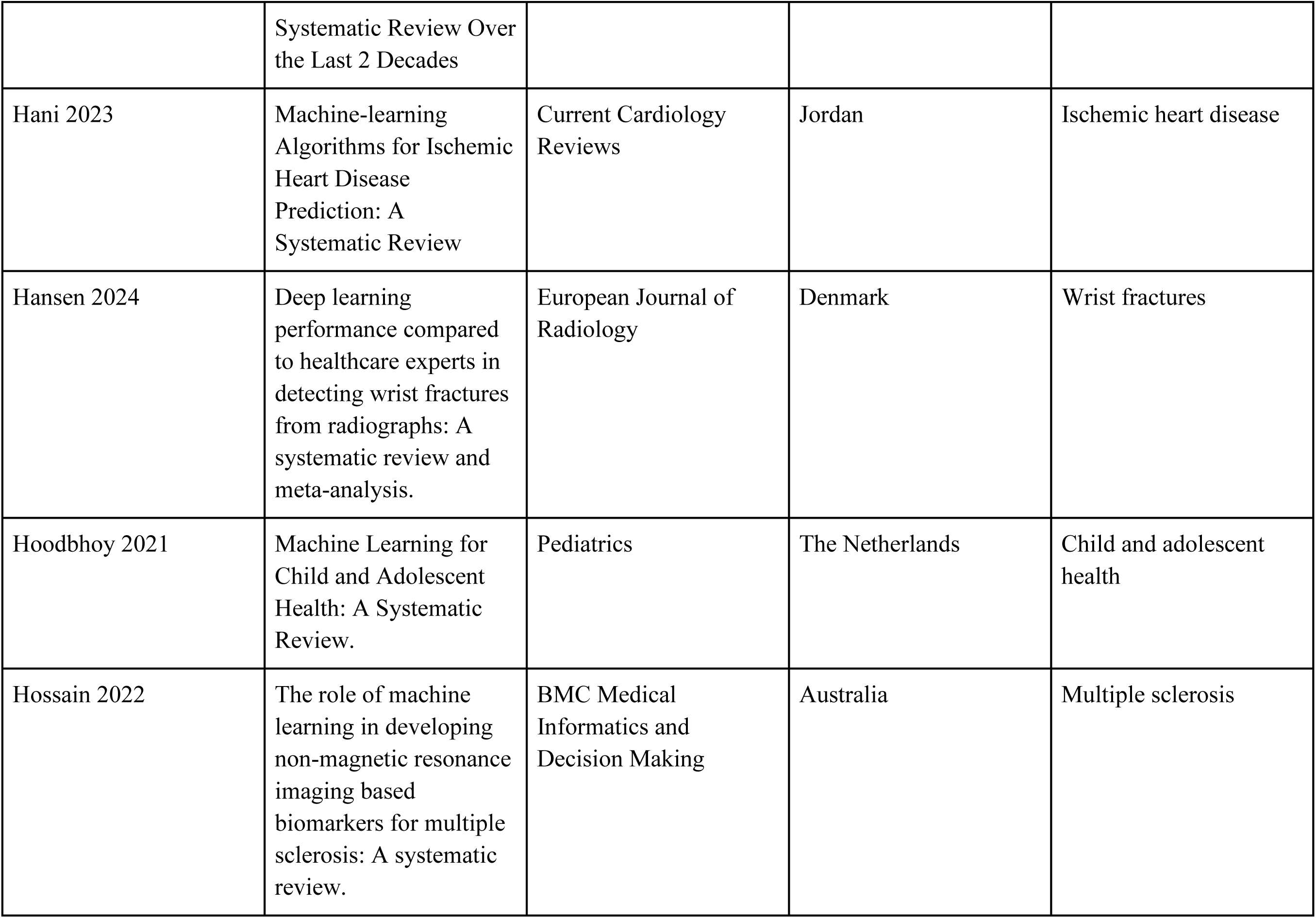
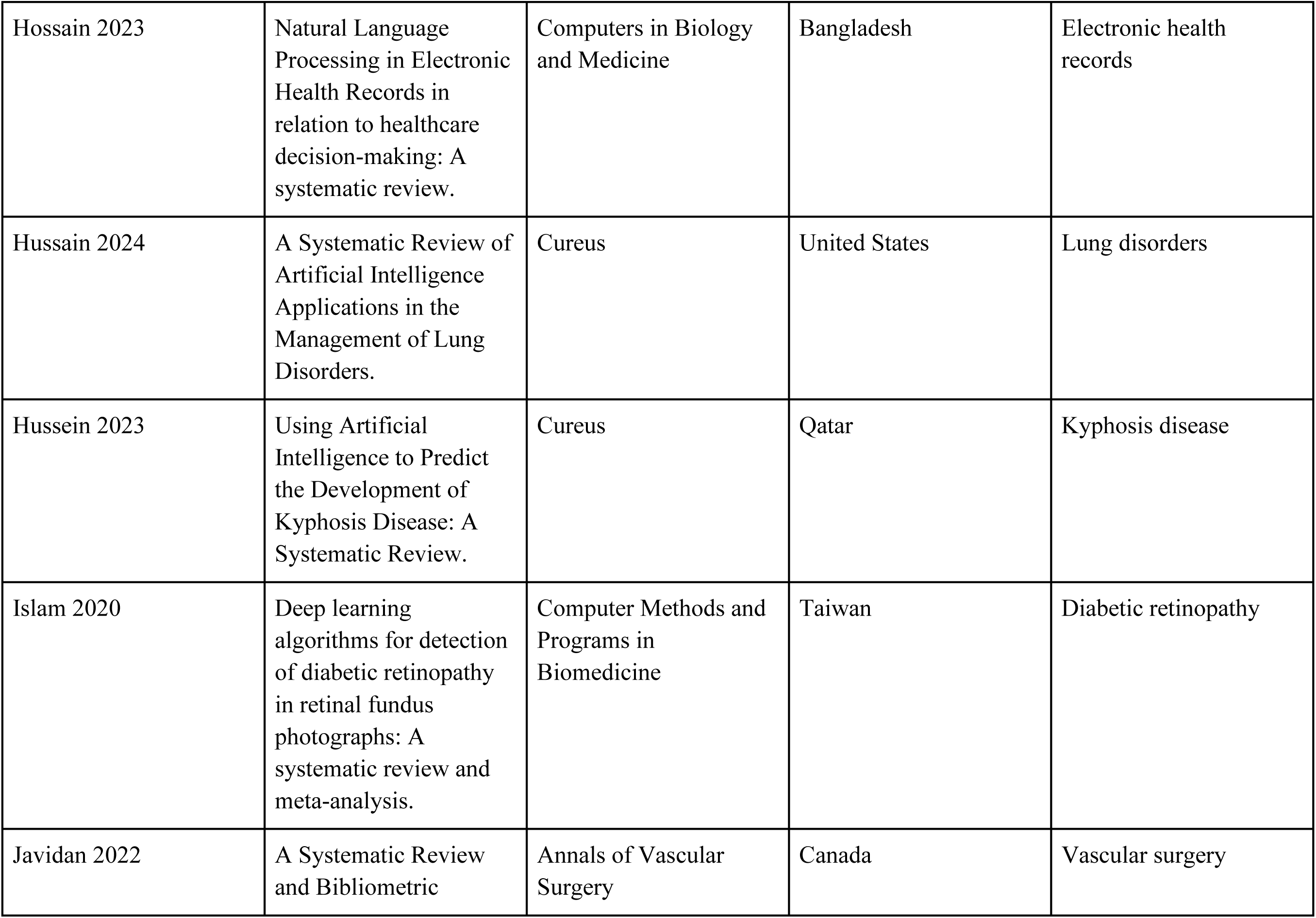
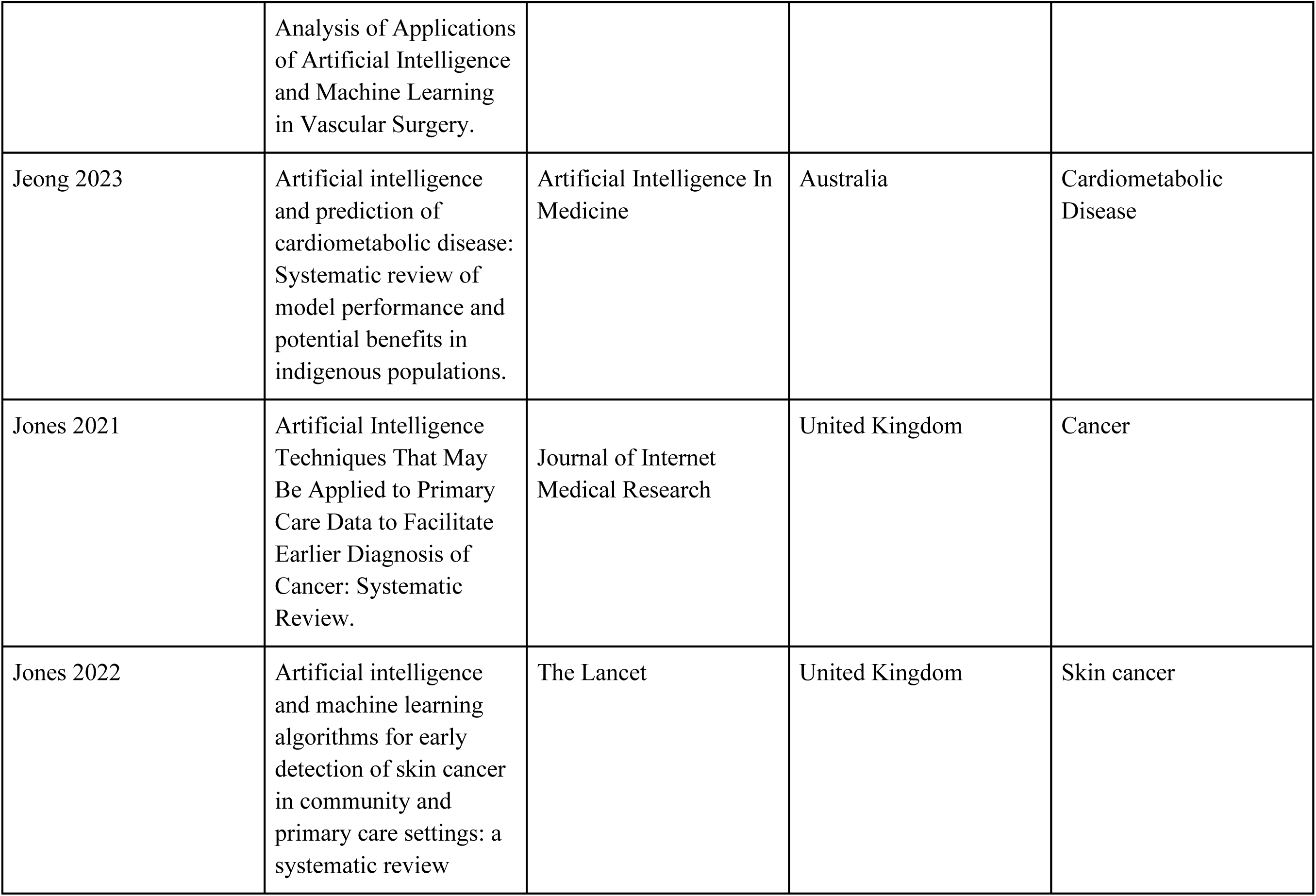
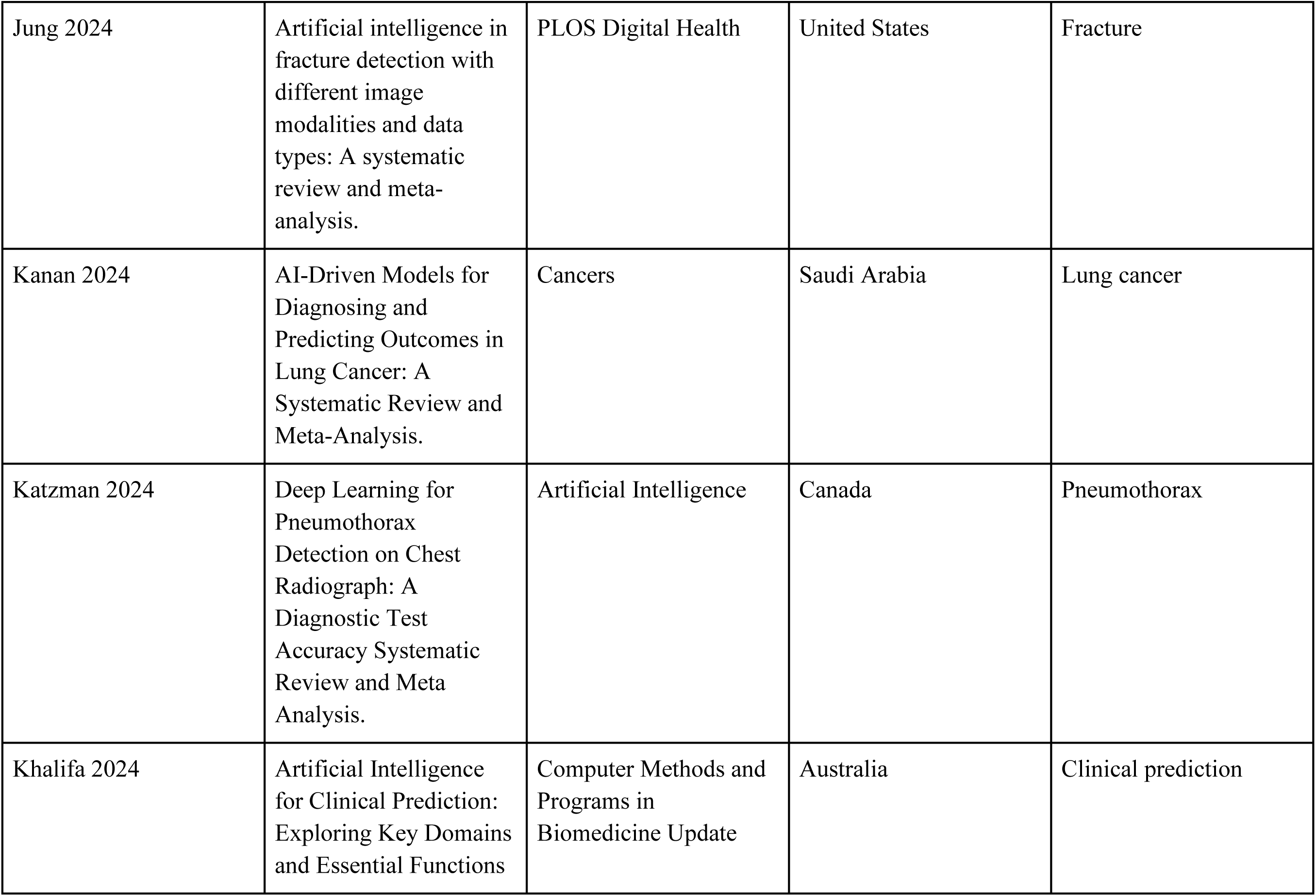
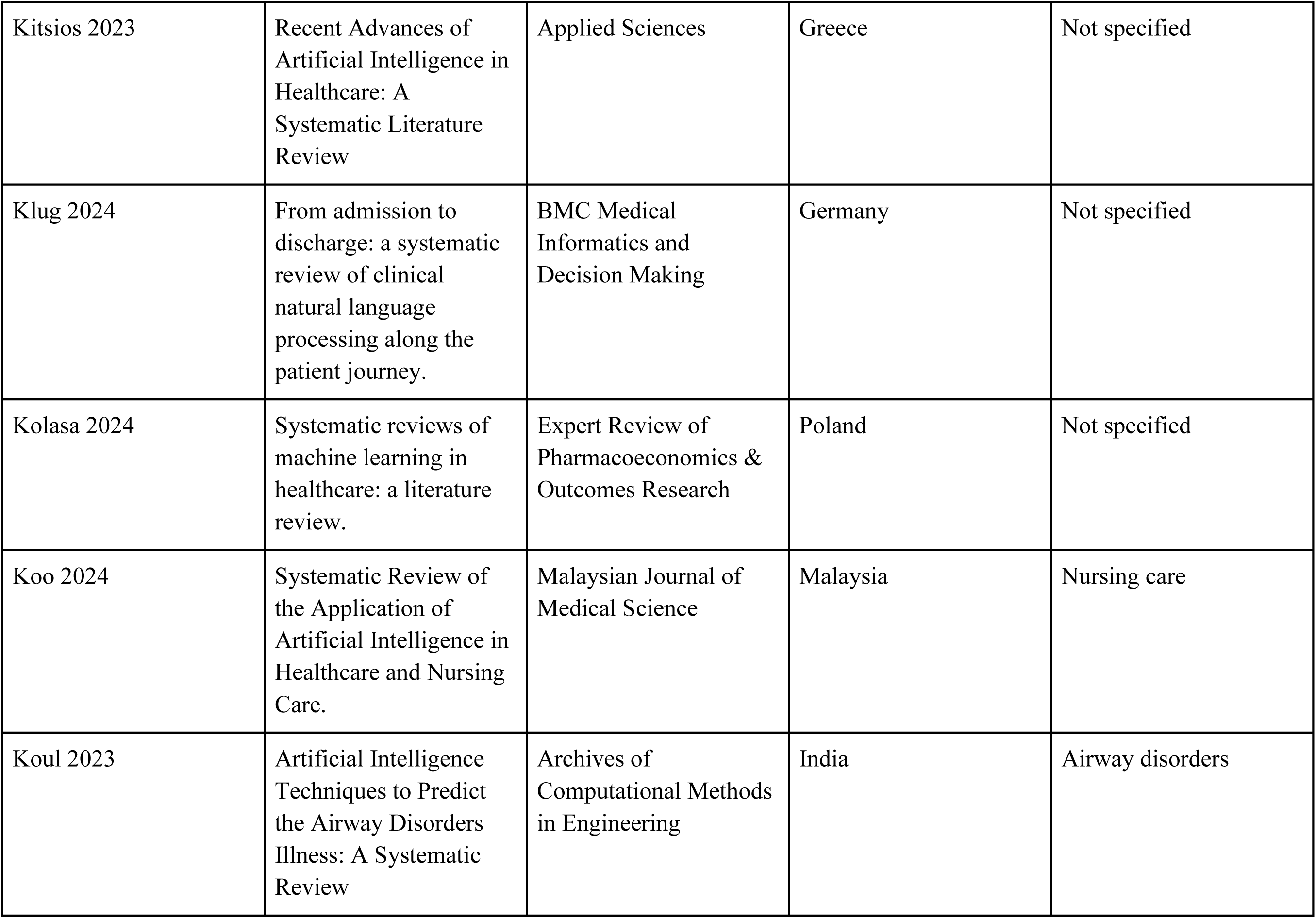
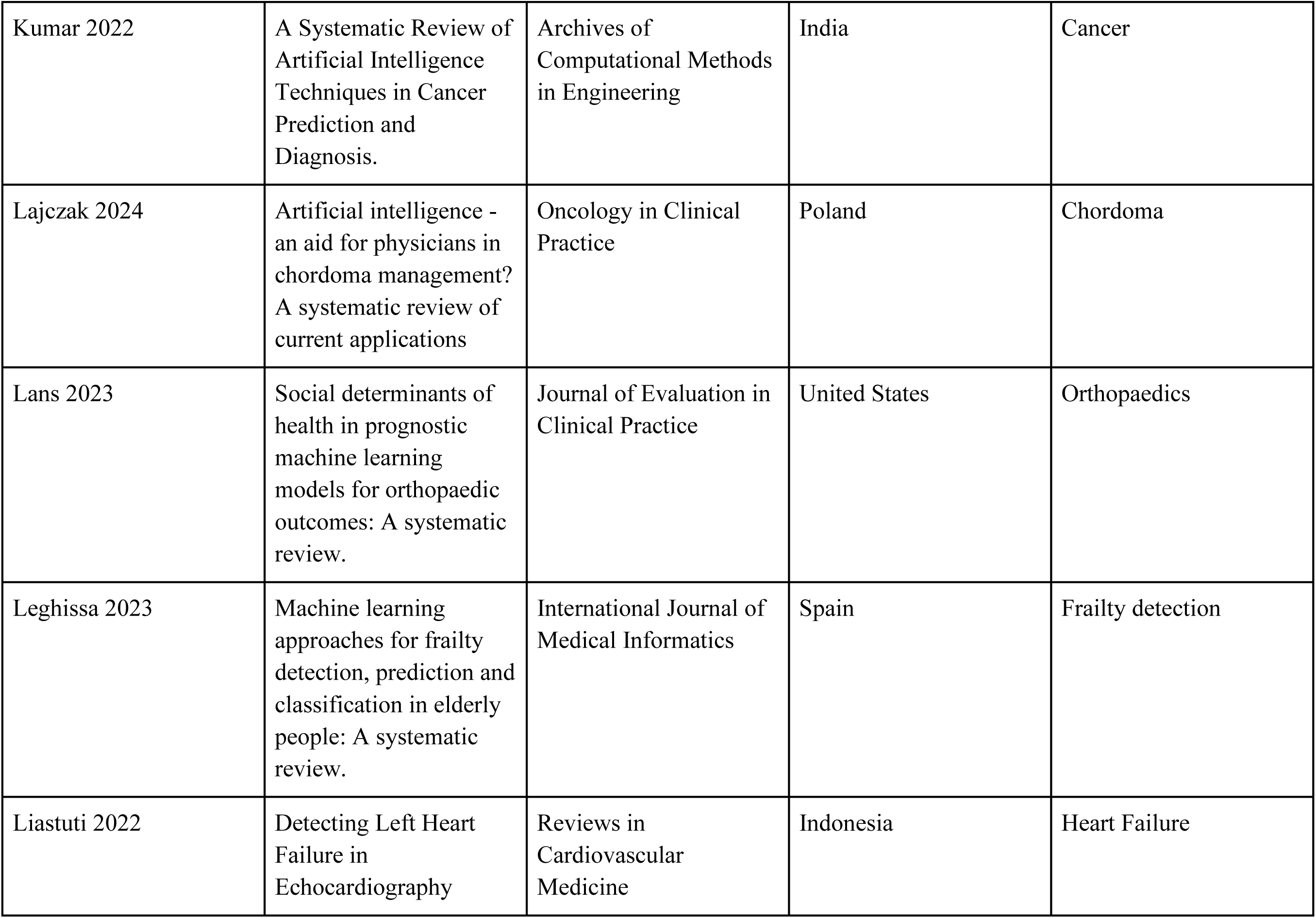
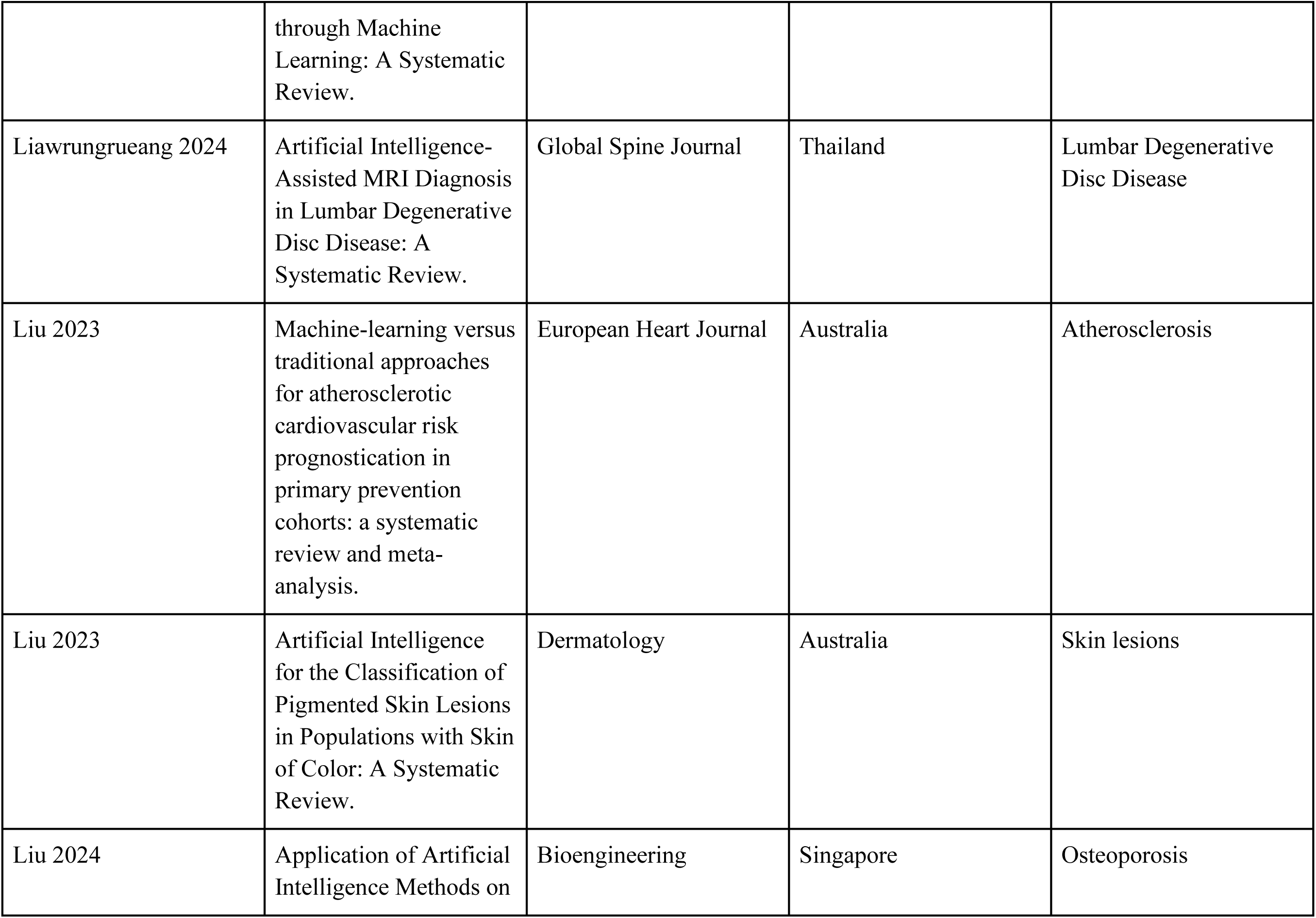
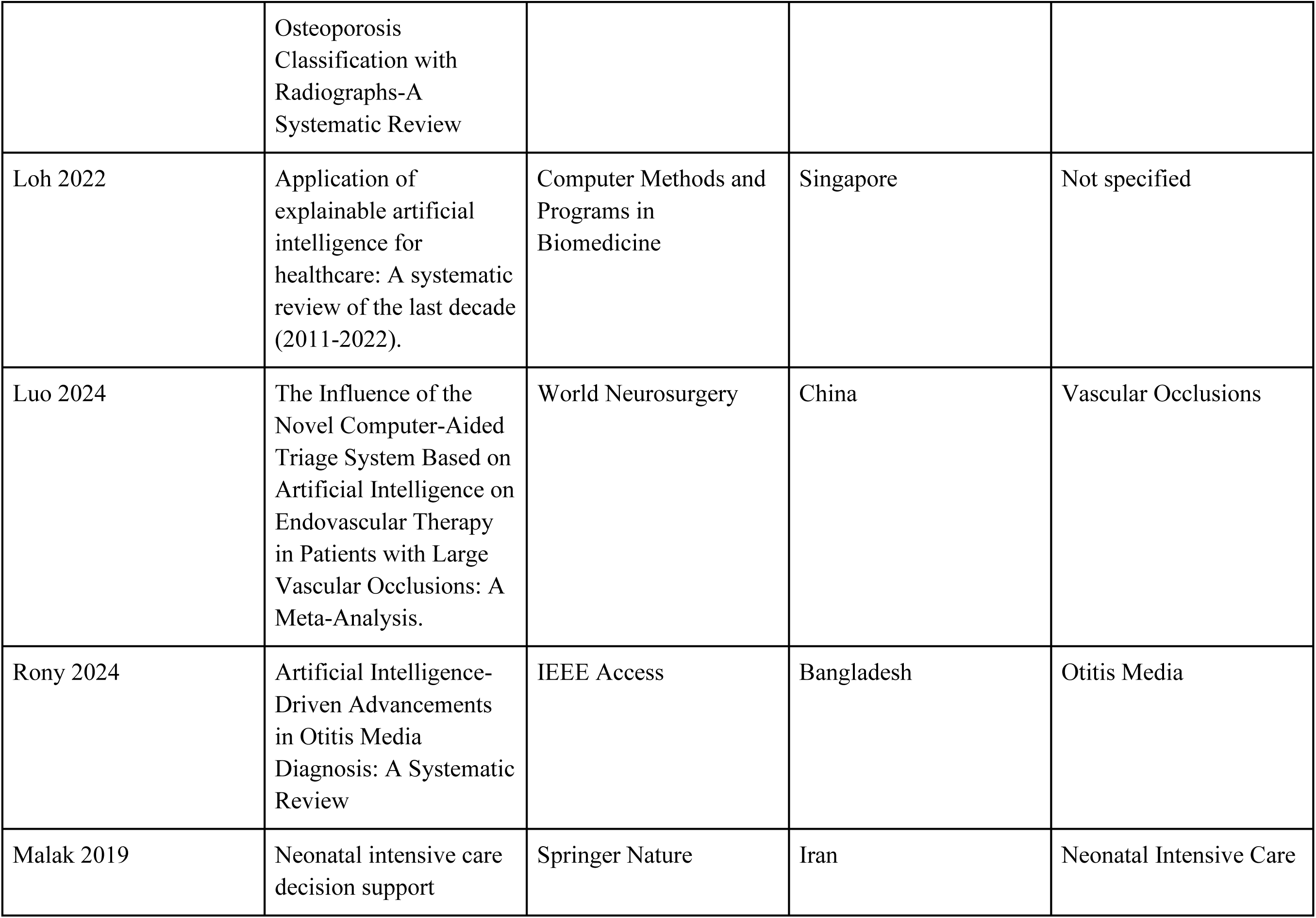
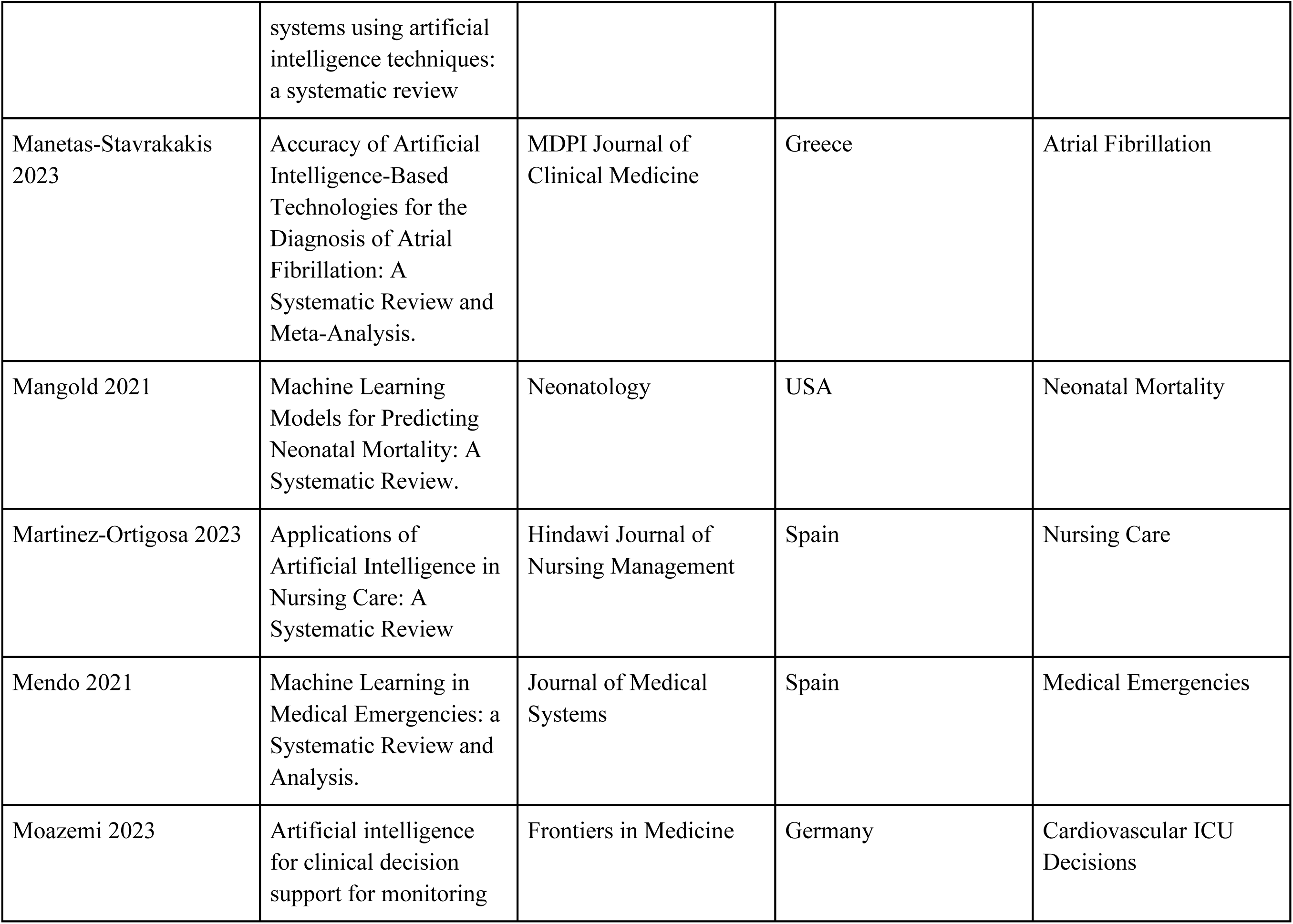
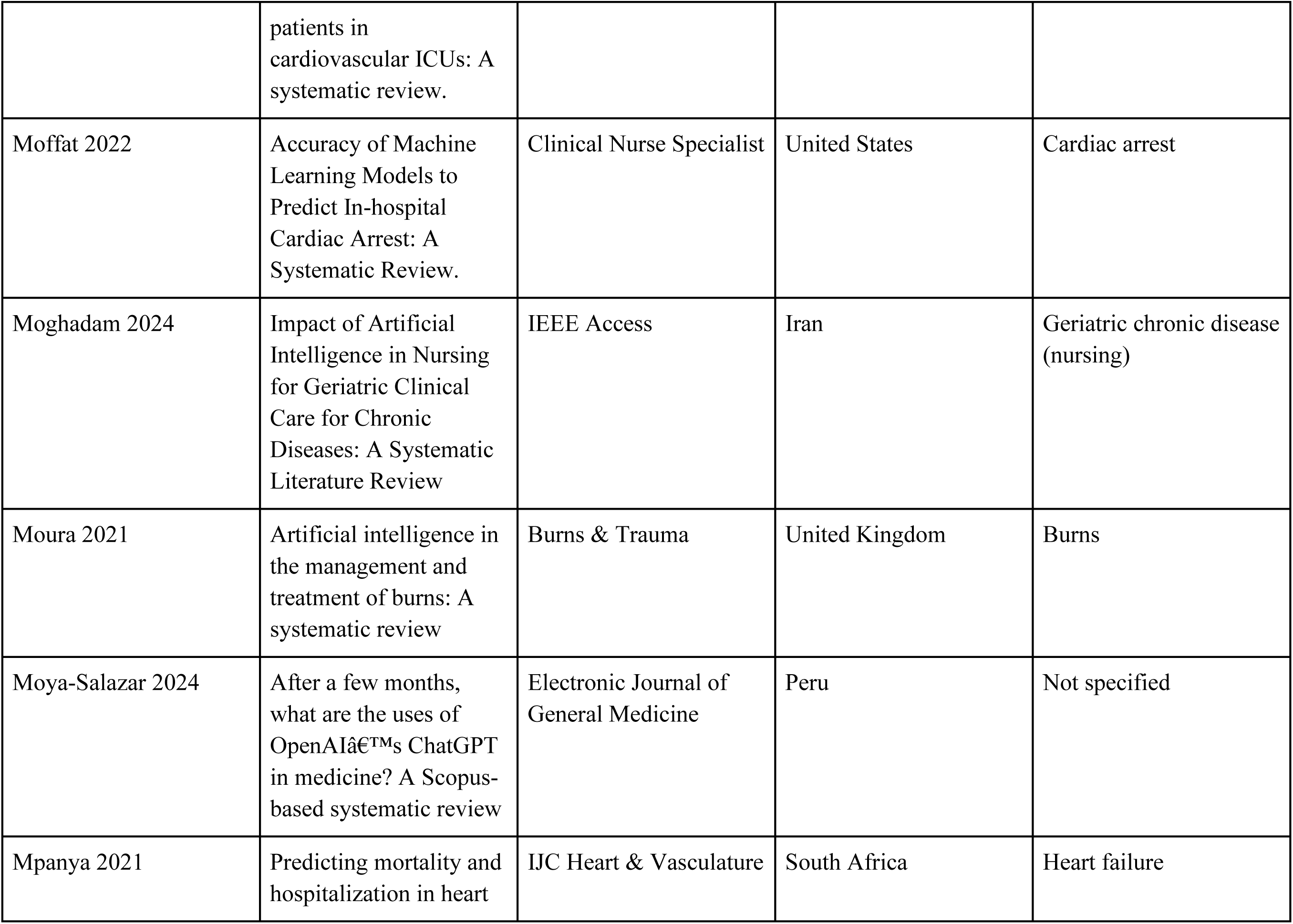
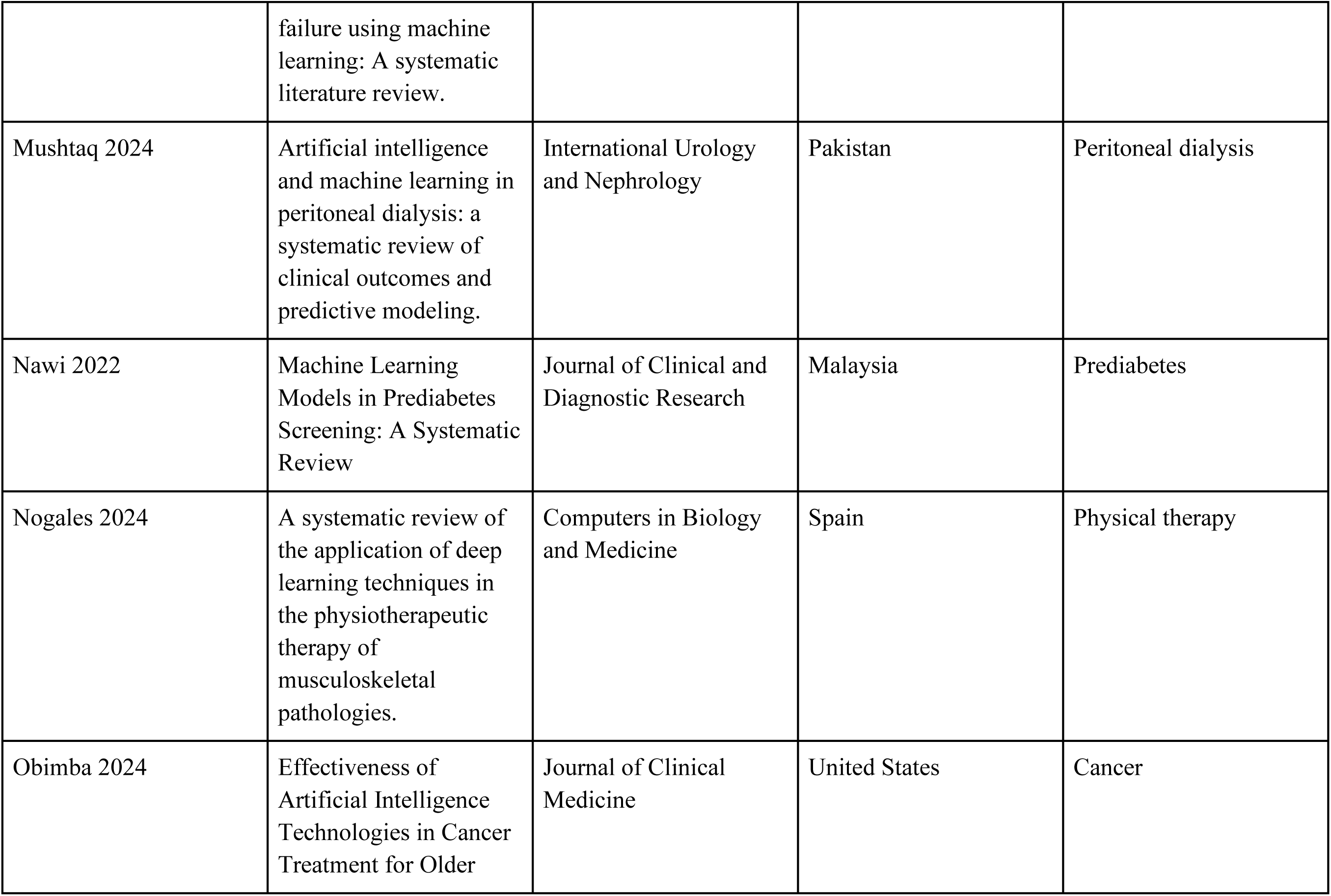
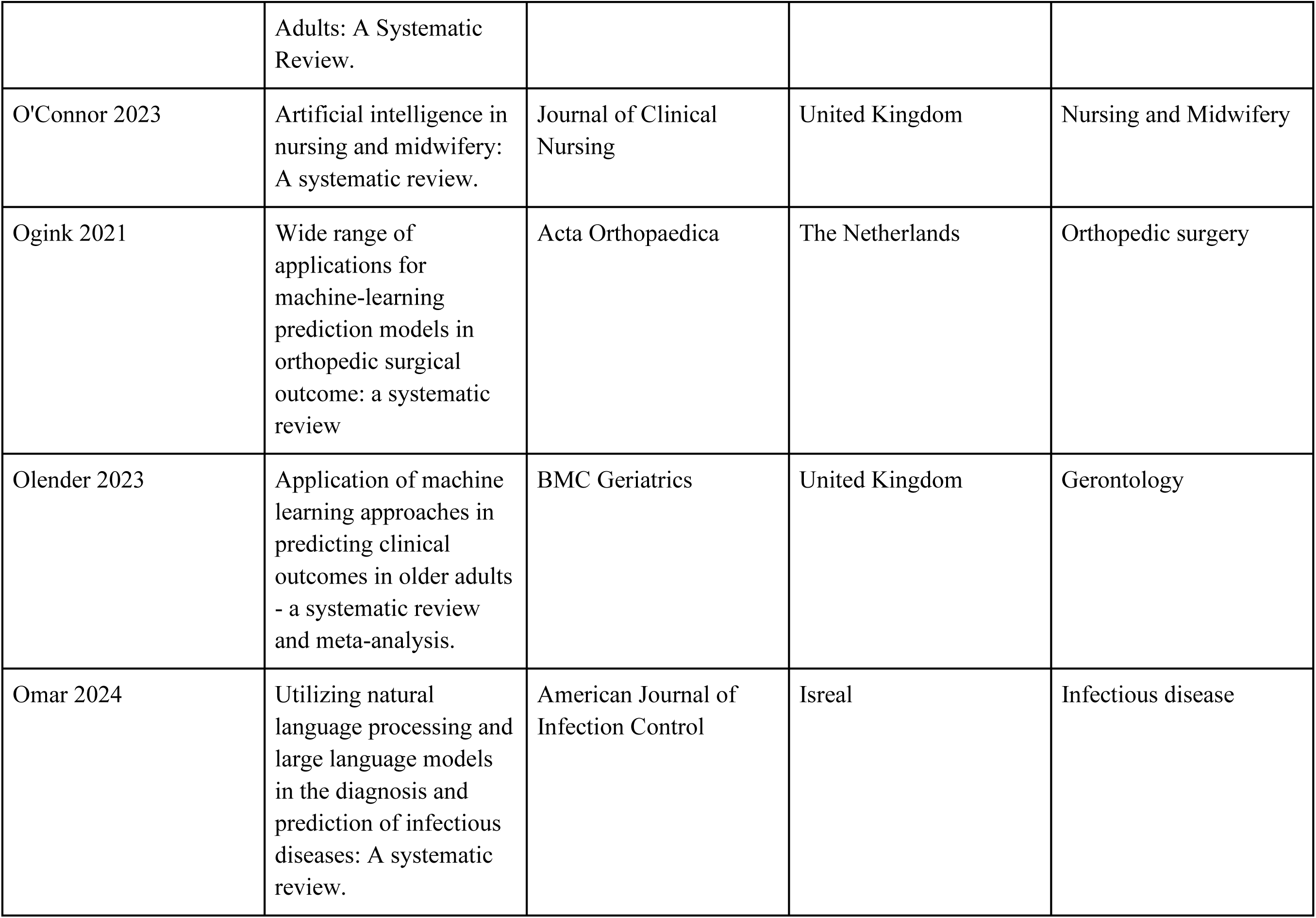
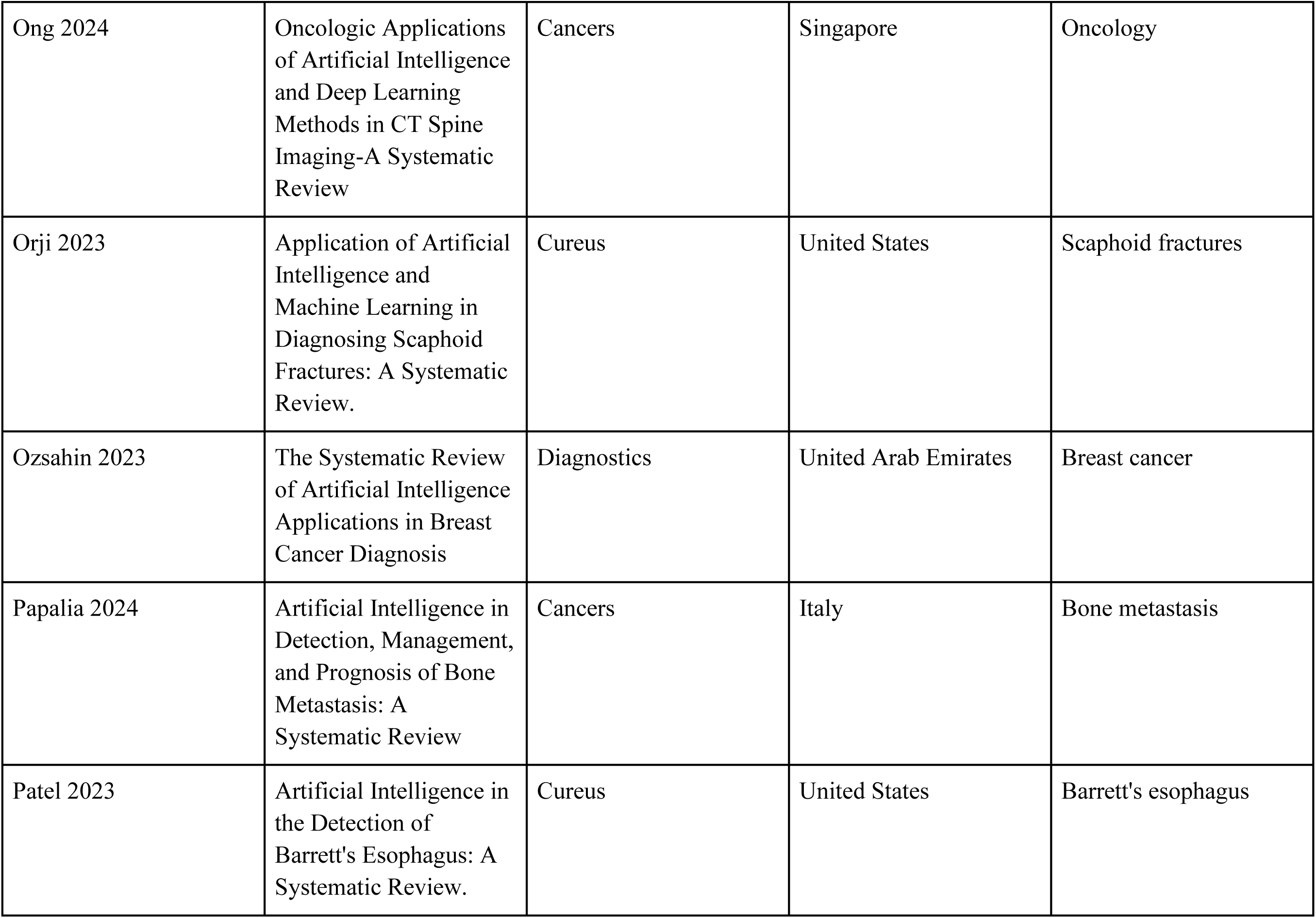
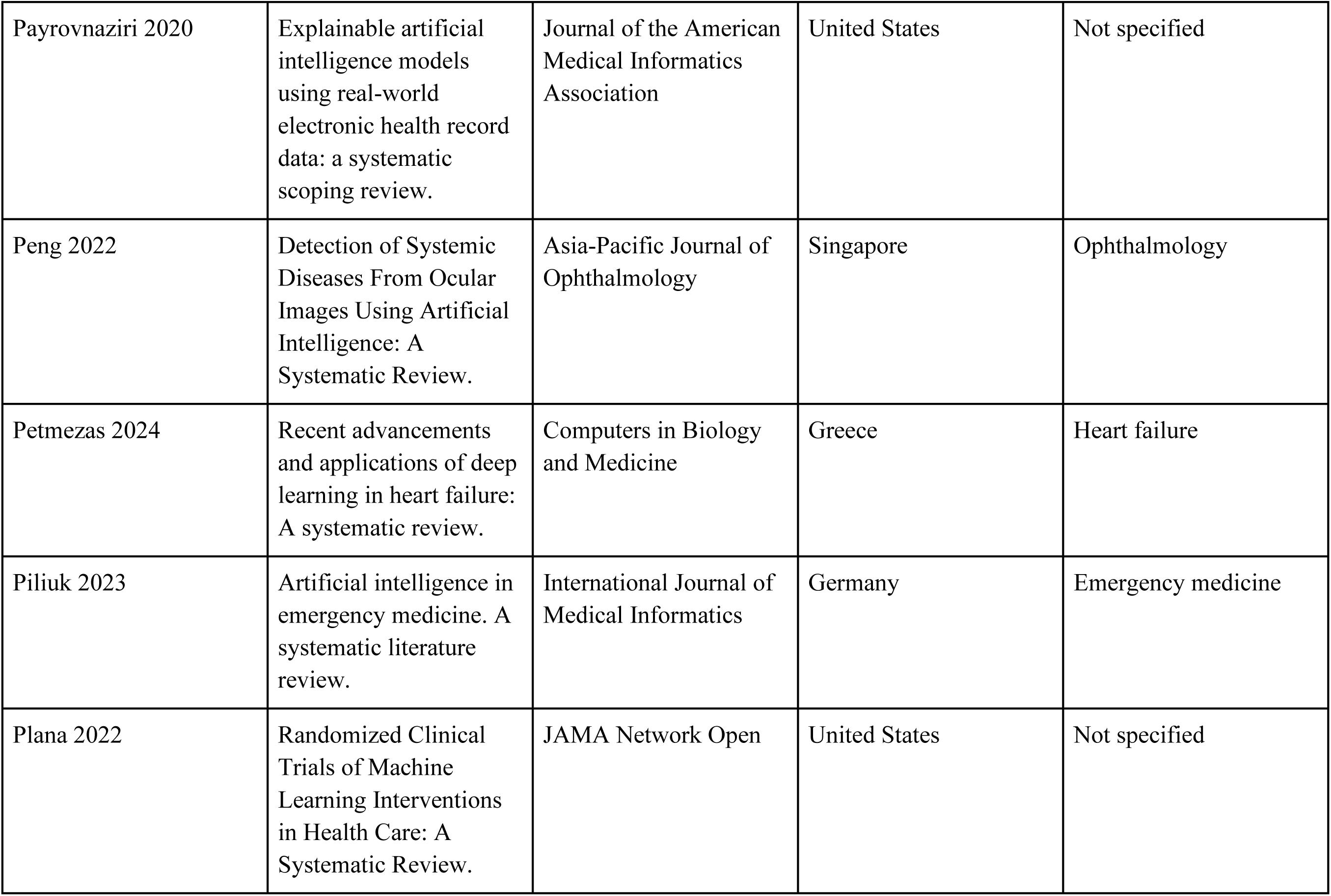
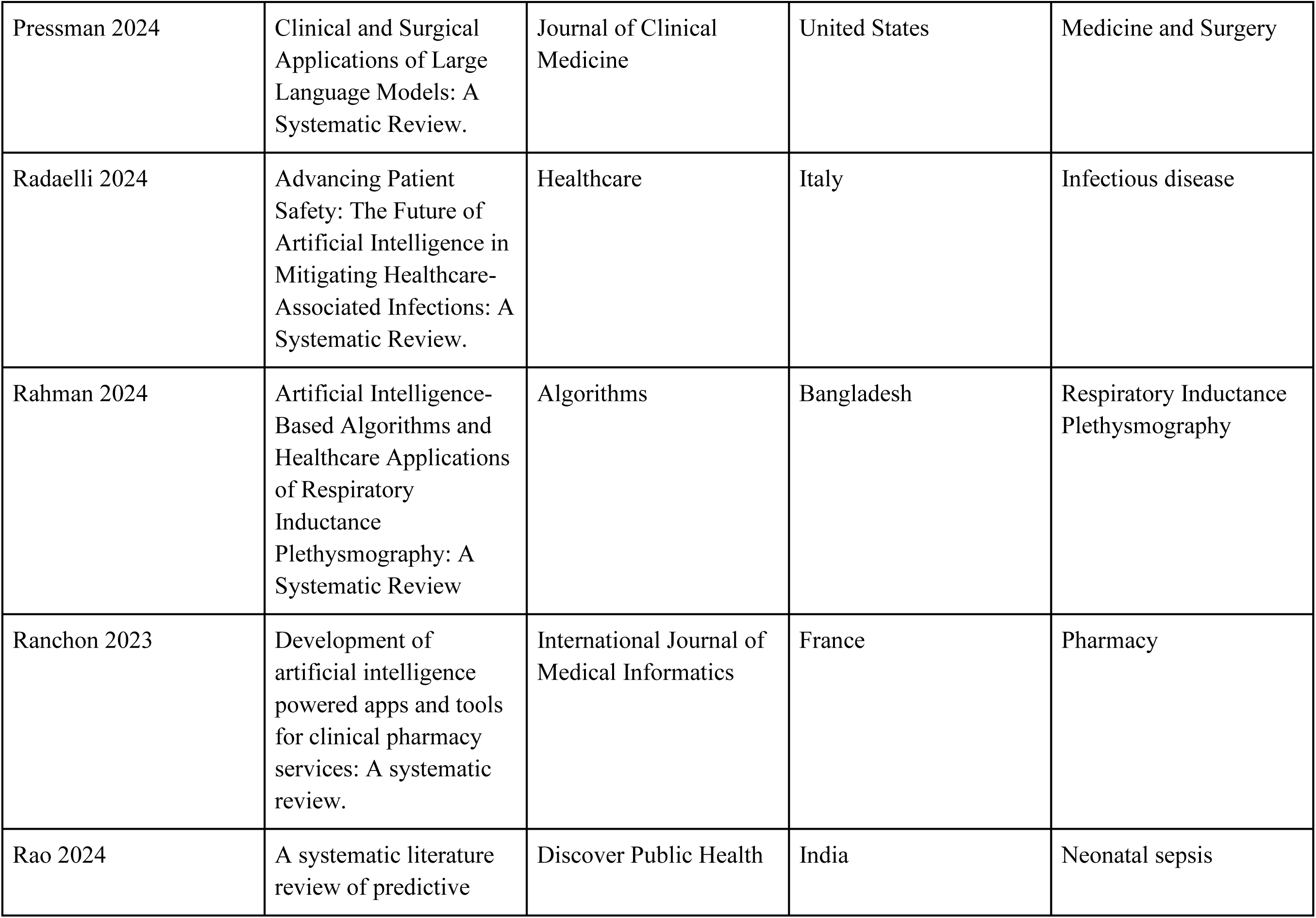
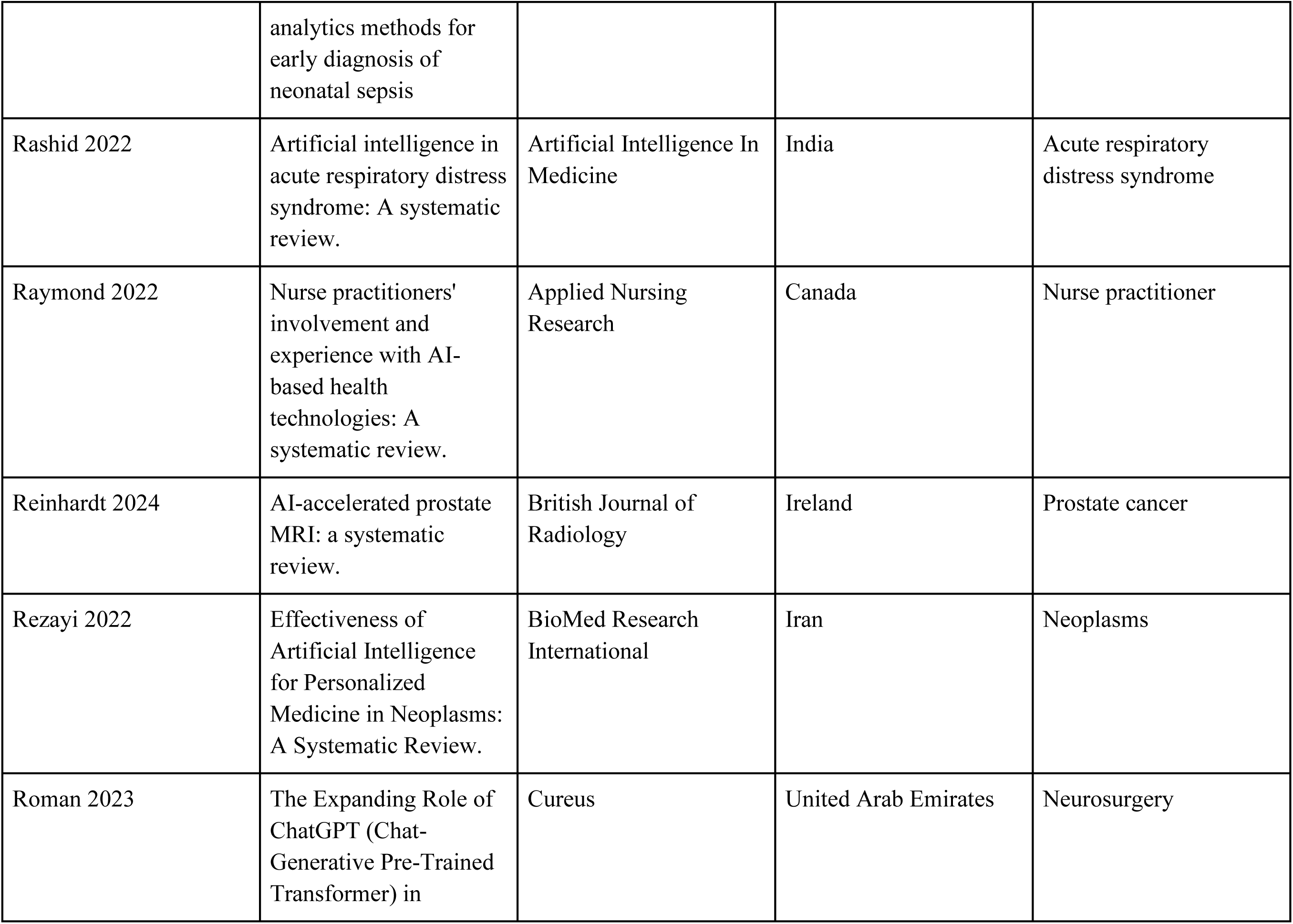
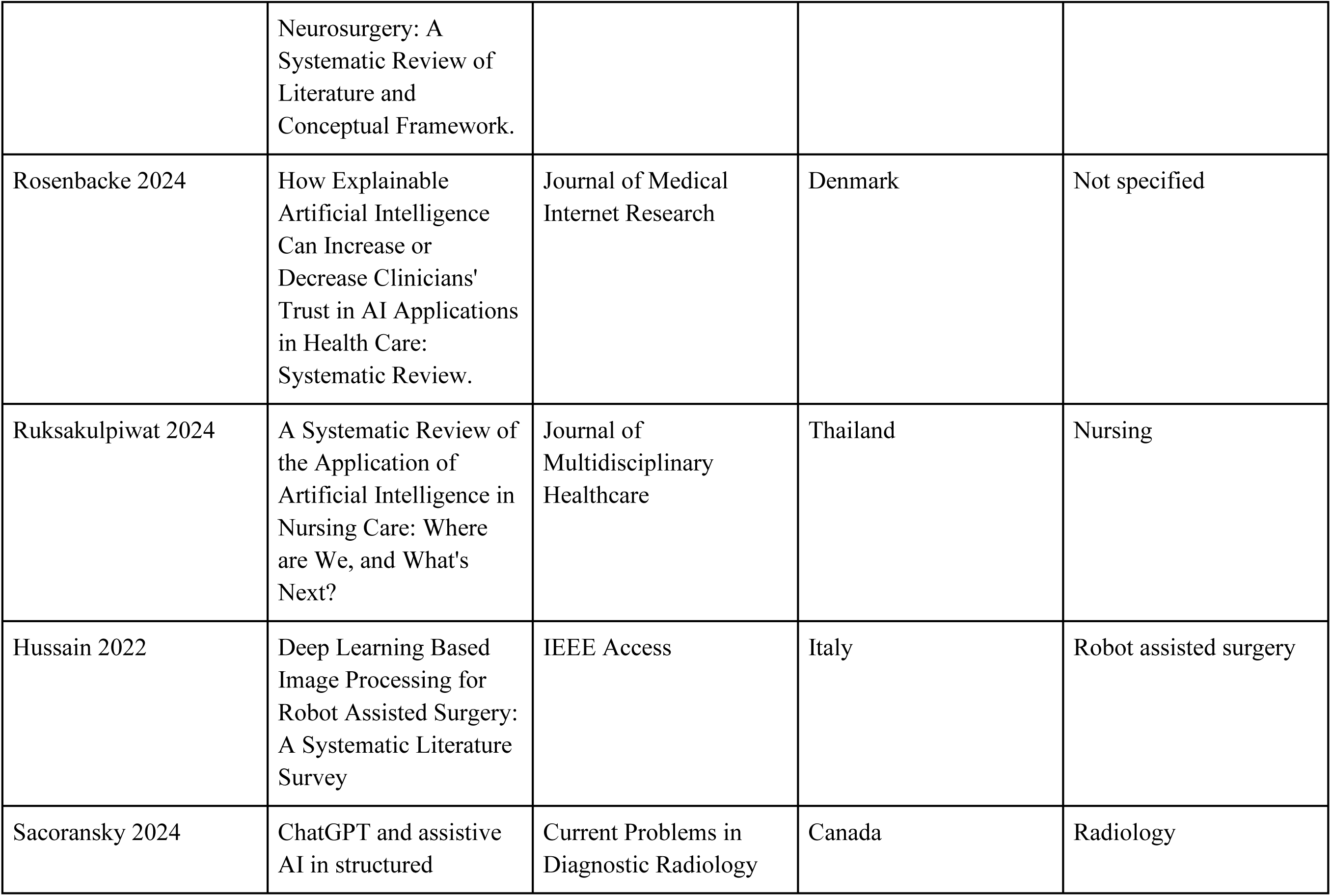
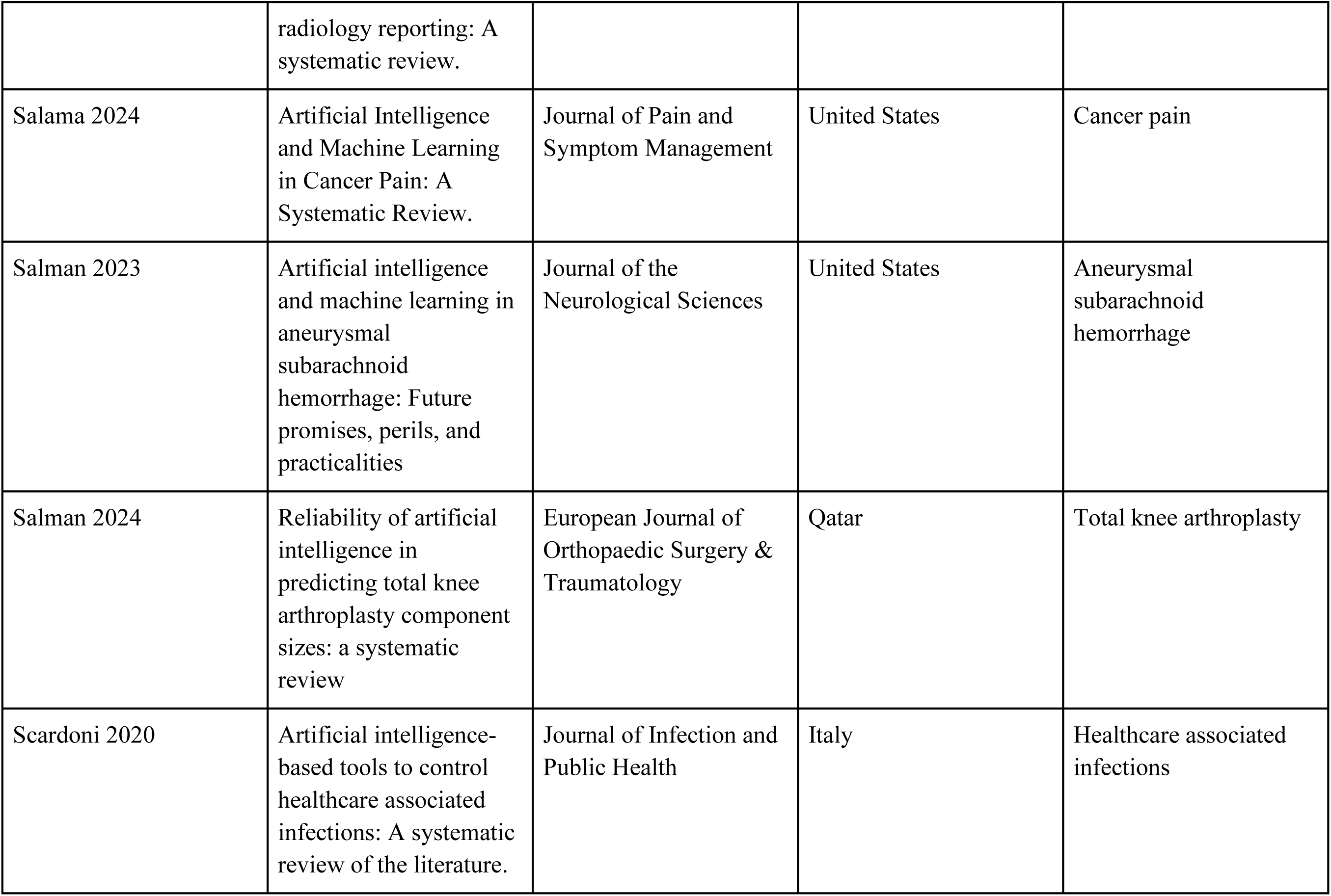
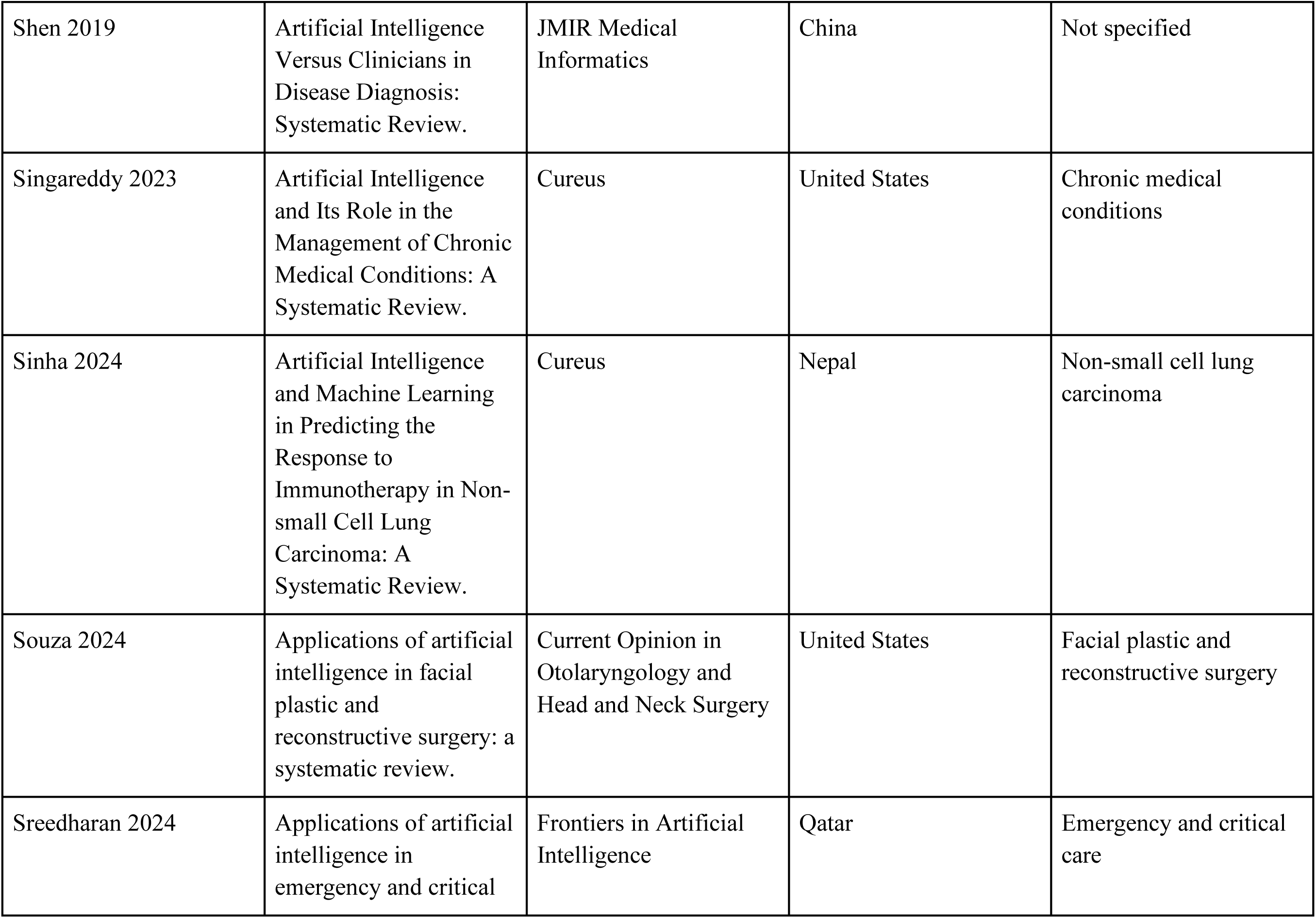
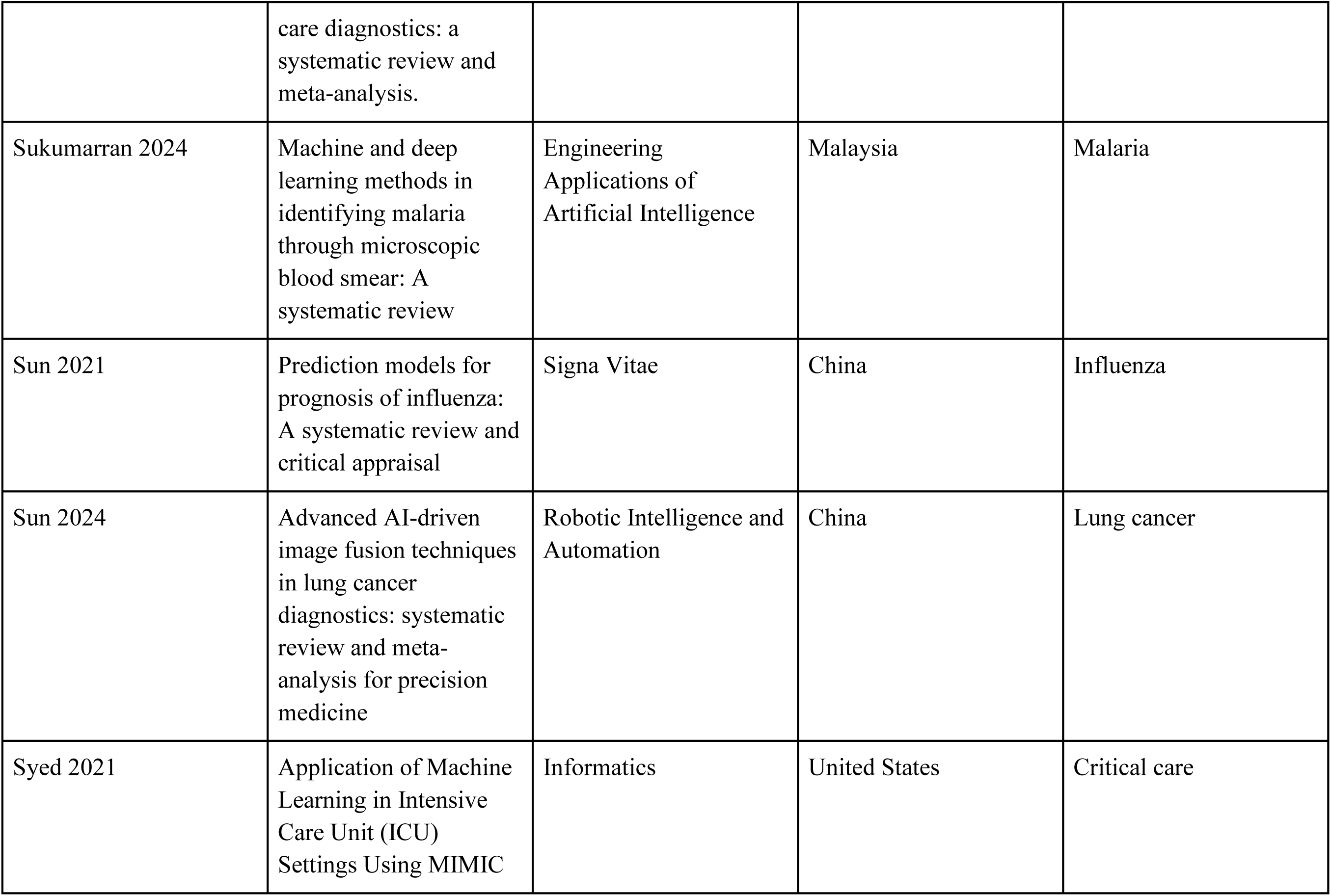
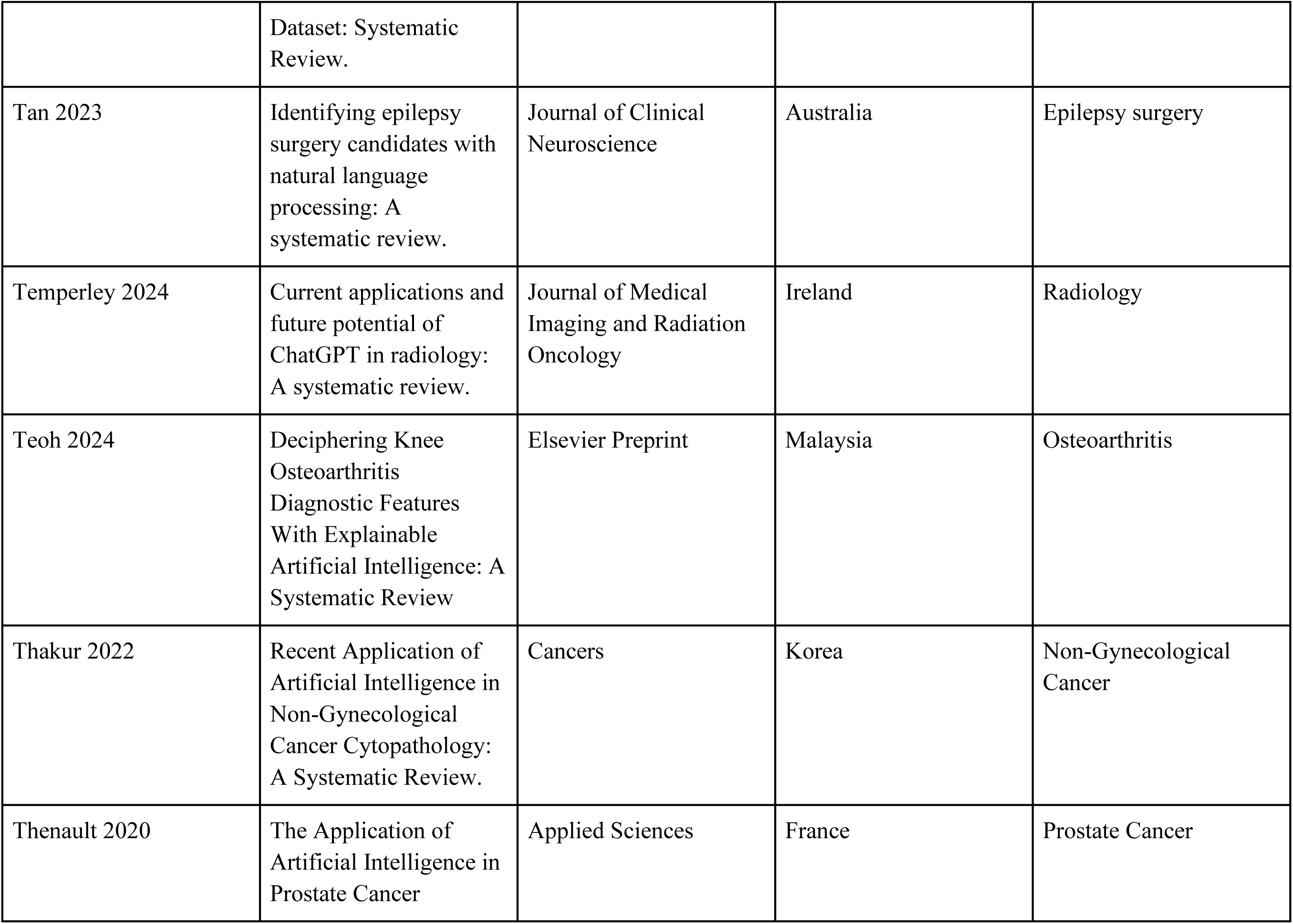
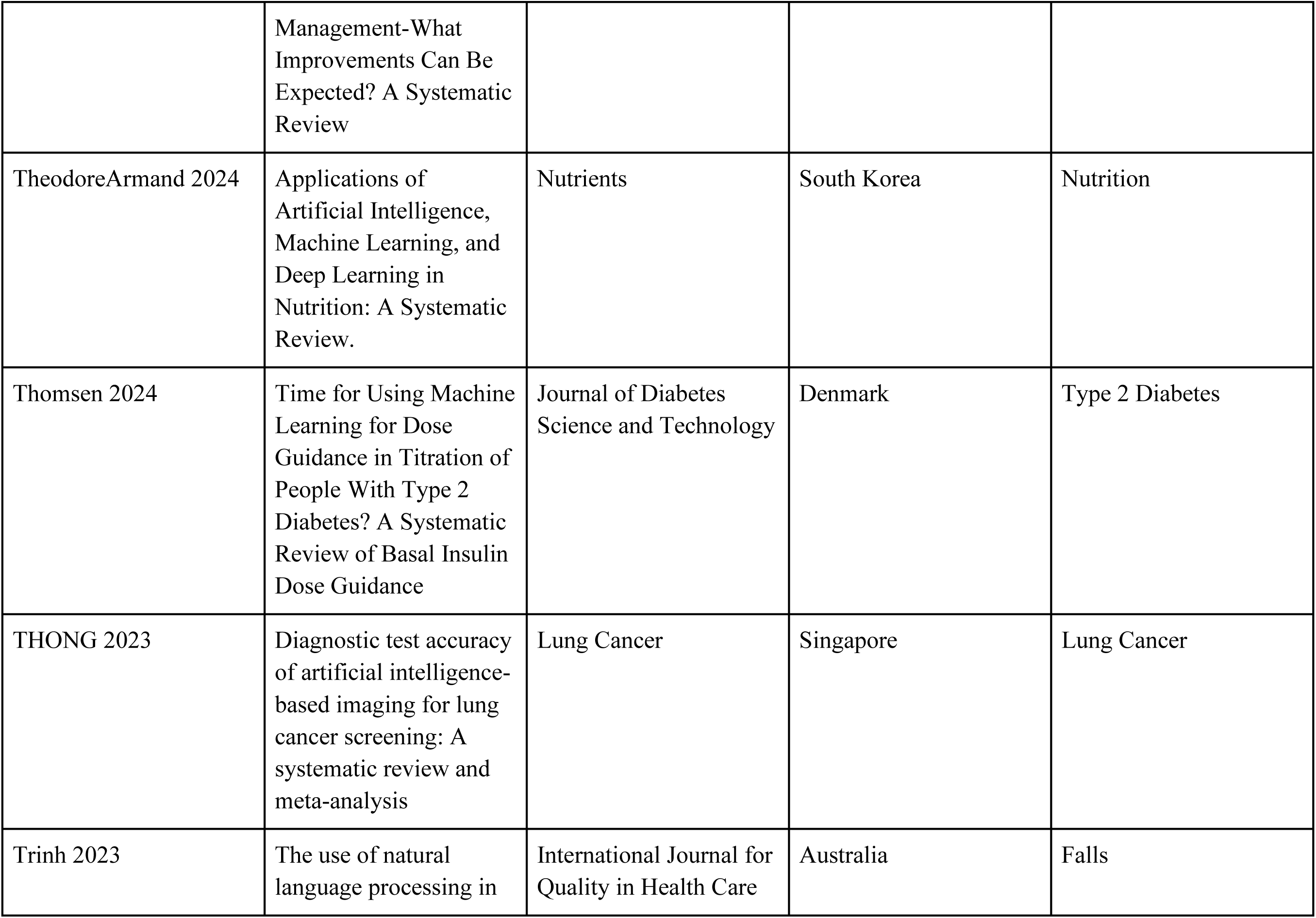
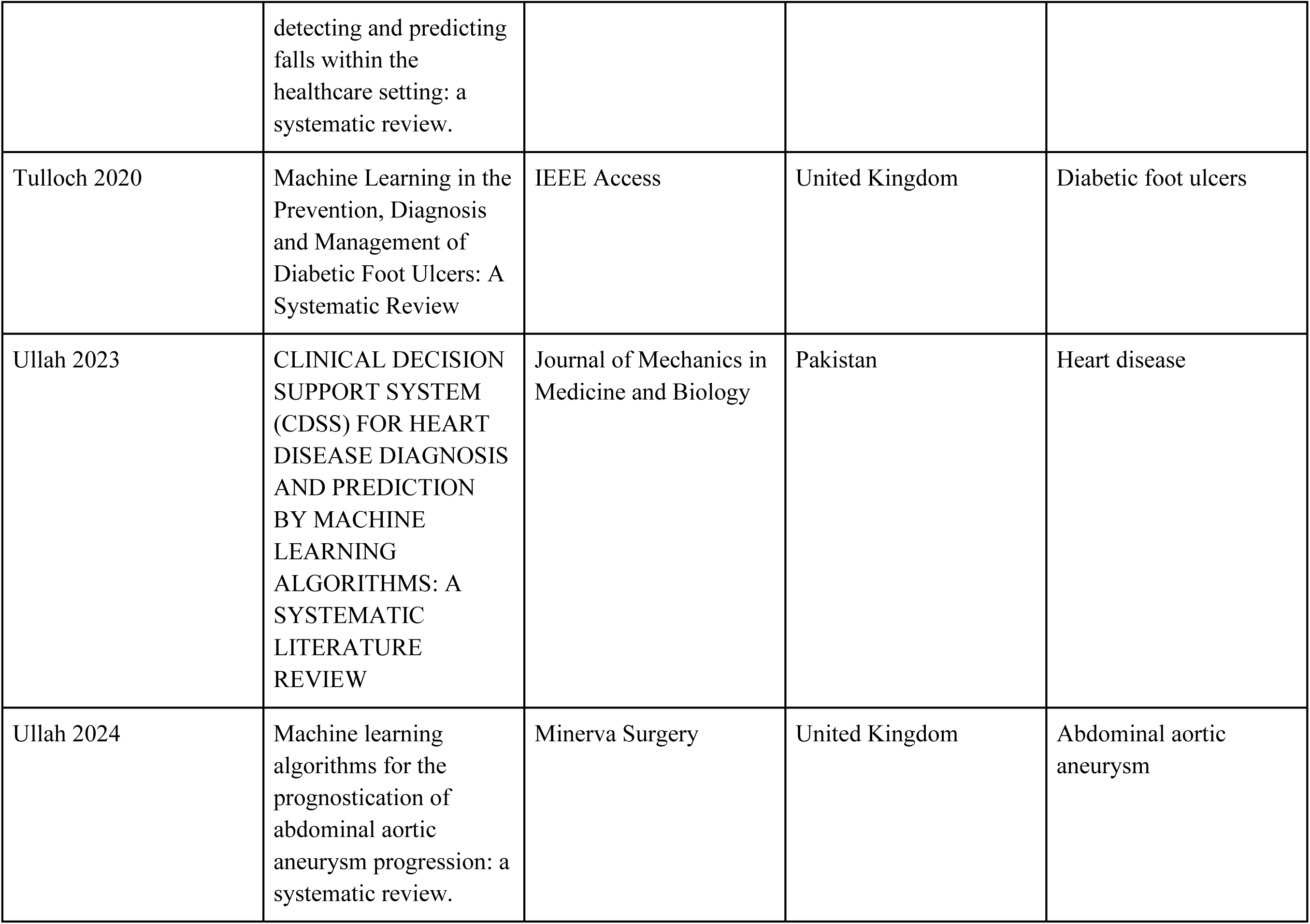
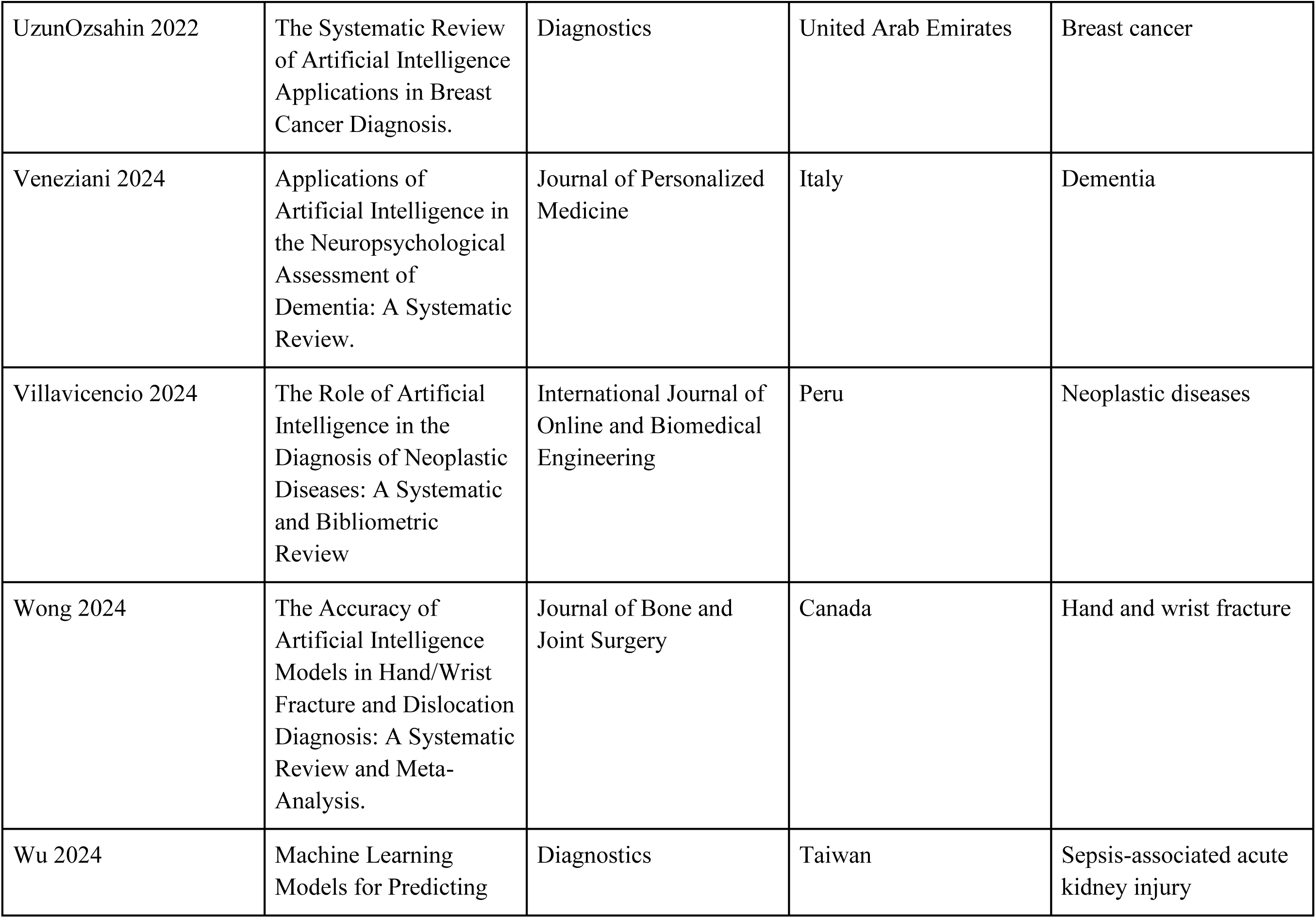
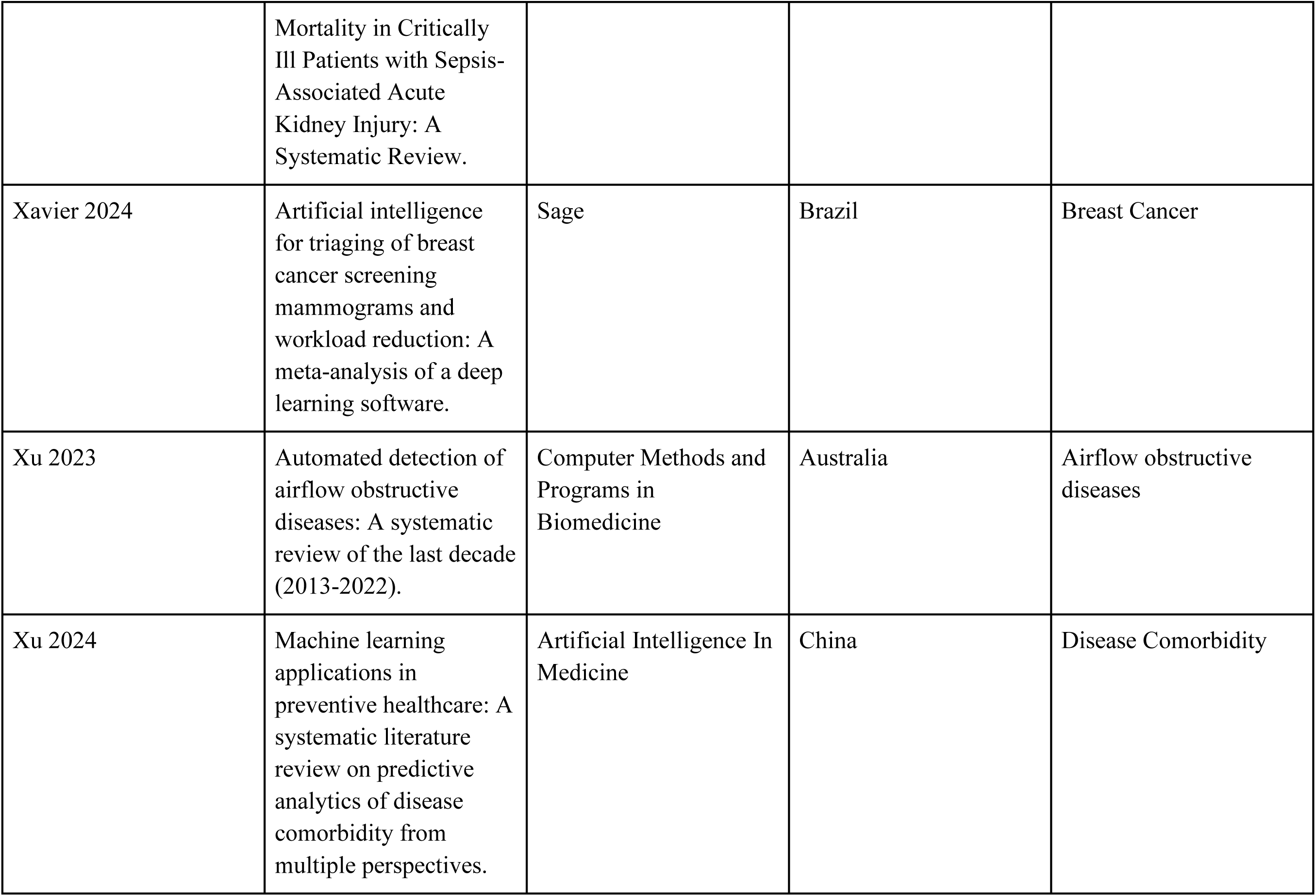
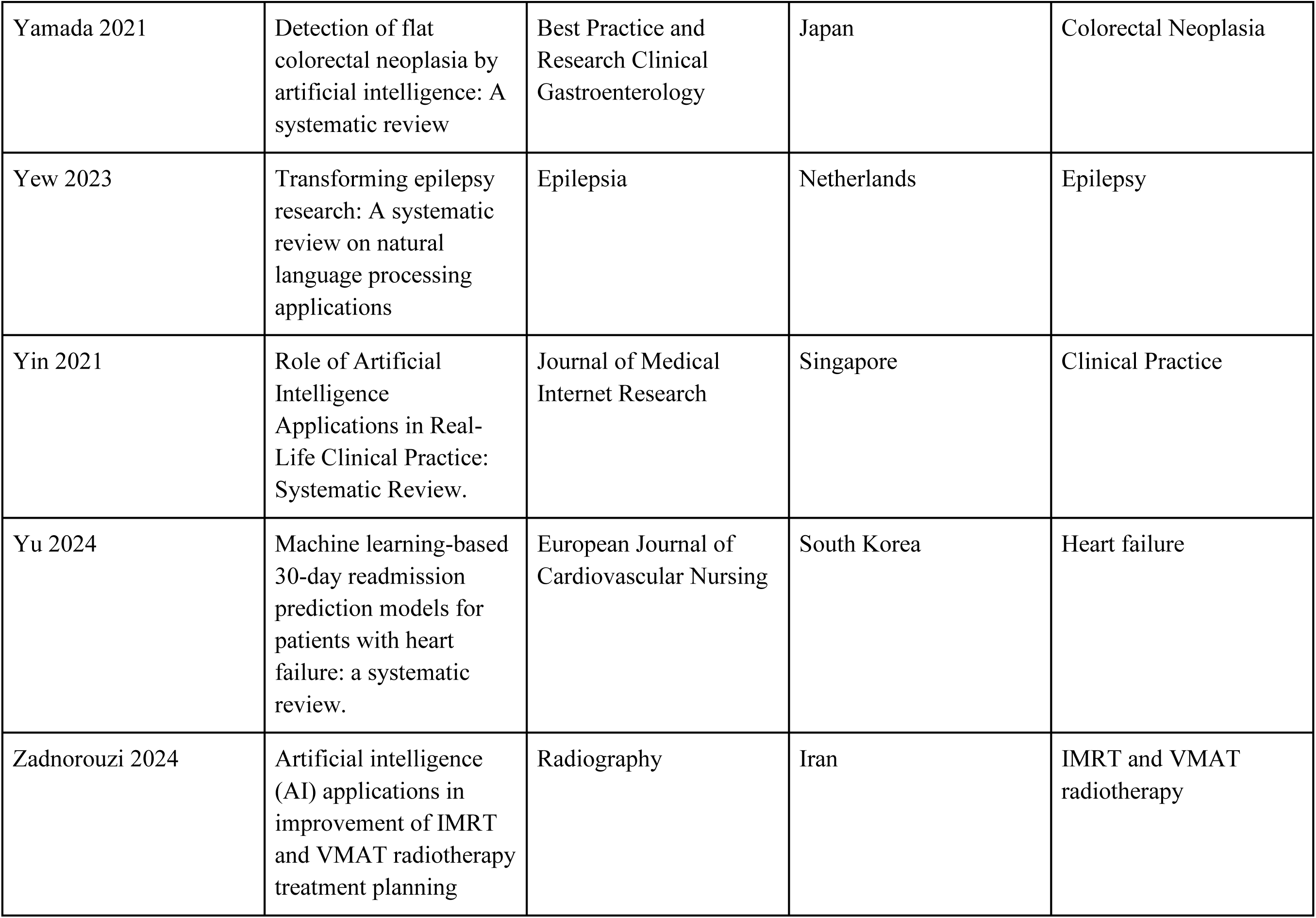
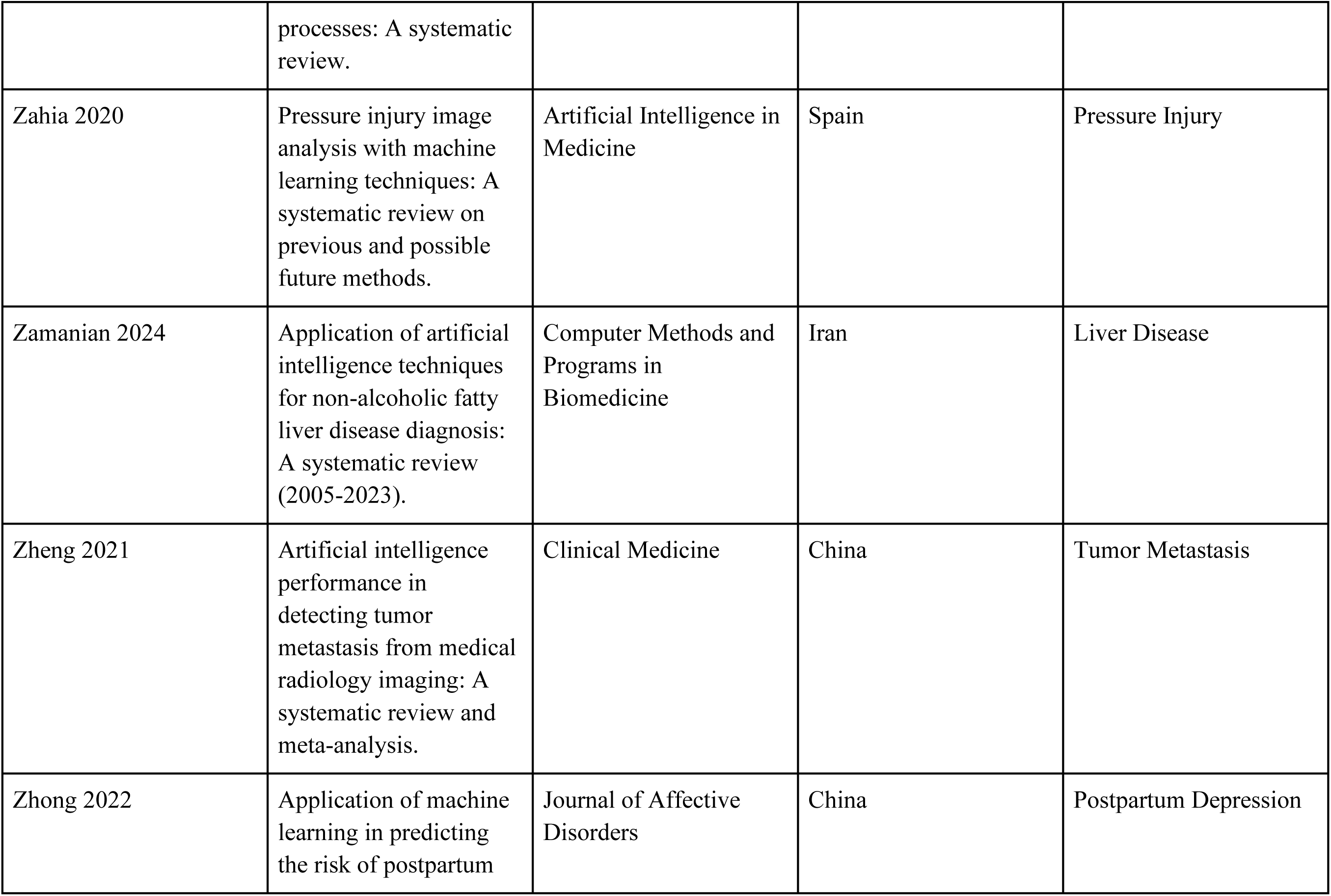
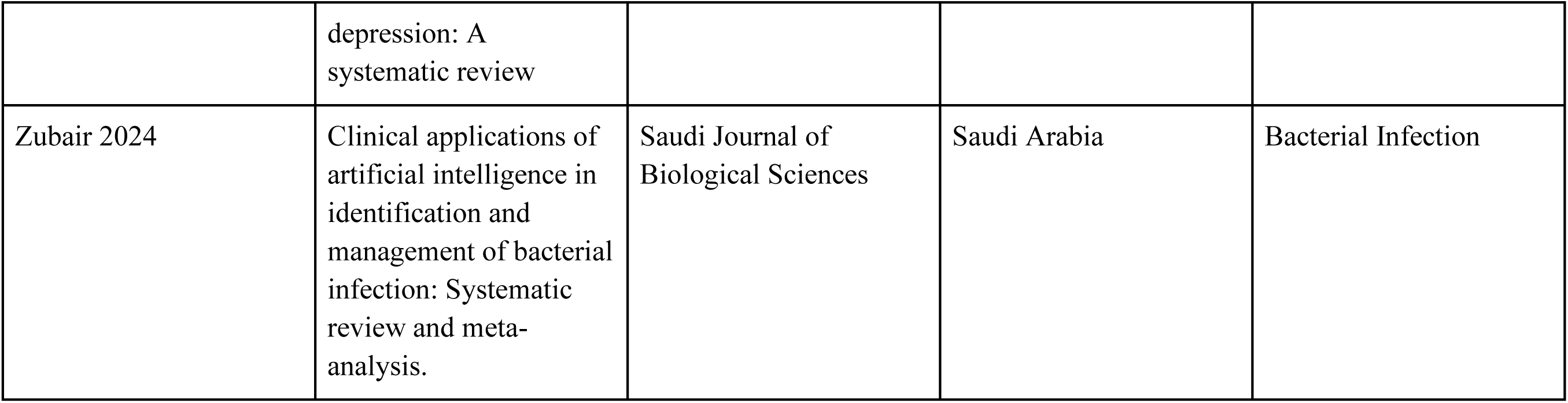

